# Longitudinal evidence for a mutually reinforcing relationship between white matter hyperintensities and cortical thickness in cognitively unimpaired older adults

**DOI:** 10.1101/2024.07.08.24309994

**Authors:** Jose Bernal, Inga Menze, Renat Yakupov, Oliver Peters, Julian Hellmann-Regen, Silka Dawn Freiesleben, Josef Priller, Eike Jakob Spruth, Slawek Altenstein, Anja Schneider, Klaus Fliessbach, Jens Wiltfang, Björn H. Schott, Frank Jessen, Ayda Rostamzadeh, Wenzel Glanz, Enise I. Incesoy, Katharina Buerger, Daniel Janowitz, Michael Ewers, Robert Perneczky, Boris-Stephan Rauchmann, Stefan Teipel, Ingo Kilimann, Christoph Laske, Sebastian Sodenkamp, Annika Spottke, Anna Esser, Falk Lüsebrink, Peter Dechent, Stefan Hetzer, Klaus Scheffler, Stefanie Schreiber, Emrah Düzel, Gabriel Ziegler

## Abstract

**Background:** For over three decades, the concomitance of cortical neurodegeneration and white matter hyperintensities (WMH) has sparked discussions about their coupled temporal dynamics. Longitudinal studies supporting this hypothesis remain nonetheless scarce.

**Methods:** In this study, we applied regional and global bivariate latent growth curve modelling (BLGCM) to longitudinal data from 436 cognitively unimpaired participants (DELCODE cohort; median age 69.70 [IQR 65.44, 74.49] years; 52.98% female) to examine the extent to which WMH and cortical thickness were interrelated over a four-year period.

**Results:** Our findings were three-fold. First, at baseline, individuals with larger WMH volumes had lower mean cortical thicknesses over the entire brain. Second, individuals who experienced a steeper thinning of their cingulate and temporal cortices over time had larger baseline WMH volumes in the frontal, parietal, and occipital lobes. Third, individuals with thinner cortices at baseline tended to undergo faster WMH progression over four years, particularly in the occipital and parietal lobes.

**Conclusions:** Our study suggests that cortical thinning and WMH progression could be mutually reinforcing rather than parallel, unrelated processes, which become entangled before cognitive deficits are detectable.

**Trial Registration:** German Clinical Trials Register (DRKS00007966, 04/05/2015)

## Introduction

Cortical thinning and white matter hyperintensities (WMH) progression are well-known ageing processes that take place throughout middle and late adulthood [1–9]. Both processes appear to be influenced by genetic and lifestyle factors [2,10–15] as well as by the onset and progression of neurodegenerative and cerebrovascular diseases [1,2,9,16–20]. Although overlapping risk factors may offer an initial explanation for their concomitance [3,6,11,21,22], their persistent association after controlling for demographics and traditional cardiovascular risk factors [3,6,10,23– 25] has sparked more than three decades of research into coupled temporal dynamics [3,26].

Coupled temporal dynamics between WMH and cortical atrophy are currently discussed from two non-exclusive perspectives: the cerebrovascular and the neurodegenerative hypotheses [17,26]. The cerebrovascular hypothesis posits that ischaemic and hypoxic damages—operationalised as WMH [15,27–29]—may initially result in the depletion of oxygen, nutrients, and trophic support in perilesional regions [16,28]. Subsequently, these damages may also disrupt the function and metabolic demands of compromised white matter tracts and associated cortical regions, leading to cortical atrophy [6,9,17,27,30]. On the other hand, the neurodegenerative hypothesis proposes that cortical neurodegeneration could contribute to WMH formation [17,26,29,31–34], especially in conjunction with tau pathologies [26,29,34]. Excessive tau phosphorylation could promote microtubule destabilisation, thereby causing axonal transport dysfunction, energy depletion, and calcium imbalance—a hallmark of Wallerian degeneration [34]. In the light of the posterior dominance of WMH in Alzheimer’s disease (AD) [26,35–38], both hypotheses would require effects of cortical neurodegeneration and WMH to be particularly pronounced in parietal and occipital brain regions. Longitudinal evidence and multivariate modelling substantiating these hypotheses remain nonetheless scarce, especially in cognitively unimpaired older adults [1].

In this study, we leveraged bivariate latent growth curve modelling (BLGCM) to examine the bidirectional relationship between lobar WMH and regional cortical thickness over four years in older individuals without objective cognitive impairment.

## Methods

### Study participants

We used baseline and annual follow-up data for up to 48 months from participants of the observational longitudinal multicentre DELCODE (DZNE Longitudinal Cognitive Impairment and Dementia) Study [39]—a memory-clinic-based observational multicentre study from the German Centre for Neurodegenerative Diseases (DZNE) that uses multimodal assessment of preclinical, prodromal, and clinical stages of AD, with a particular focus on subjective cognitive decline. In the present work, we focused on cognitively unimpaired participants who underwent at least three MRI scanning sessions and whose follow-up MRI sessions took place within four months prior or after their yearly comprehensive examination.

At baseline, participants underwent a thorough evaluation at their local study site, which included medical history checks, a psychiatric and neurological examination, neuropsychological testing, blood and cerebrospinal fluid (CSF) collection, and MRI in accordance with local standards. All DELCODE sites used the Consortium to Establish a Registry for AD (CERAD-plus) neuropsychological test battery to assess cognitive function. To be classified as cognitively unimpaired, participants were defined by performing within at least required to performed better than -1.5 standard deviations of the age-, sex-, and education-adjusted normal performance on all subtests of the test battery [39].

Additional inclusion criteria were age□≥□60 years, fluent German language skills, capacity to provide informed consent, and the the availability of a study partner. The main exclusion criteria for all groups were conditions clearly interfering with participation in the study or the study procedures, including significant sensory impairment. The following medical conditions were considered exclusion criteria: current or history of major depressive episode and major psychiatric disorders either at baseline (e.g., psychotic disorder, bipolar disorder, substance abuse), neurodegenerative diseases other than AD, vascular dementia, history of stroke with residual clinical symptoms, history of disseminated malignant disease, severe or unstable medical conditions, and clinically significant vitamin B12 deficiency at baseline. Prohibited drugs included chronic use of psychoactive compounds with sedative or anticholinergic effects, use of anti-dementia agents, and investigational drugs for the treatment of dementia or cognitive impairment one month before study entry and throughout the duration of the study.

All participants provided their written informed consent in accordance with the Declaration of Helsinki at baseline. DELCODE has been registered within the German Clinical Trials Register (DRKS00007966, 04/05/2015). Ethics committees of the medical faculties of all participating sites (i.e., Berlin (Charité - Universitätsmedizin Berlin), Bonn, Cologne, Göttingen, Magdeburg, Munich (Ludwig-Maximilians-University), Rostock, and Tübingen) approved the DELCODE study protocol before inclusion of the first participants. The ethics committee of the medical faculty of the University of Bonn led and coordinated the process.

### Total cardiovascular risk score

We established a total cardiovascular risk score for each participant by tallying their dichotomised (y/n) history of smoking, presence of obesity, hyperlipidemia, arterial hypertension, and diabetes, as reported in their medical records. We corrected the sum of present risk factors by the amount of available information. For example, if an individual had a history of arterial hypertension and diabetes but we did not have data on smoking, obesity, or hyperlipidemia, the final score would be 1.00. The corrected total cardiovascular risk scores ranged from 0.0 to 1.0, where the lowest and highest values denoted the absence or presence of all available risk factors, respectively.

### MRI

MRI acquisition took place at nine DZNE sites or associated university medical centers equipped with 3T Siemens MR scanners. In the present study, we leveraged the following structural sequences: T1w MPRAGE (full head coverage, 3D acquisition, GRAPPA factor 2, 1 mm^3^ isotropic, 256 × 256 px, 192 sagittal slices, TR/TE/TI 2500/4.33/1100 ms, FA 7°) and T2w FLAIR (full head coverage, 3D acquisition, 1 mm^3^ isotropic, 256 × 256 px, 192 sagittal slices, TR/TE/TI 5000/394/1800 ms). The DZNE imaging network oversaw operating procedures, as well as quality assurance and assessment (iNET, Magdeburg) [39].

### MRI-based measurements

#### Cortical thickness

We used the CAT12 longitudinal pipeline [40] (neuro-jena.github.io) to reconstruct cortical thickness surfaces for each subject and for each time point (ageing workflow; default parameters, except for final resolution, which we set to 1 mm^3^). We smoothed all surfaces with a 12-mm Gaussian filter and resampled them to the 32k HCP surface template. We also used CAT12 to estimate mean thickness throughout the whole brain cortex and within cortical regions as per the Desikan-Killiany cortical parcellation atlas [41].

#### WMH segmentation

We manually segmented WMH using the AI-augmented version of the Lesion Segmentation Toolbox (LST-AI) [42–44] and based the segmentation on both T1w MPRAGE and T2w FLAIR imaging data. We then tallied WMH volumes across the frontal, temporal, parietal and occipital lobes using the UCSLobes Atlas [45].

### Statistical analyses

We conducted all data analyses in RStudio (v1.3.1073; R v4.0.2) using lavaan (v0.6-16). We created figures using ggplot2 (v3.4.3) and the ENIGMA toolbox [46].

We carried out univariate and bivariate LGCM analyses. (B)LGCMs [47] are a powerful class of structural equation models (SEM) to describe sample average trajectories of one or two constructs over time through the specification of latent intercepts and latent slopes (i.e, initial levels and rates of change). First, we used univariate analyses for contextualisation to examine what covariates were associated with the baseline measurements, as well as with potential changes over repeated measures. We then focused on bivariate models to examine interrelationships between WMH and cortical thickness over time (Figure 1). Intra-domain and cross-domain relationships can be assessed via the covariance between these four latent growth parameters [48]. We specifically tested the evidence of four possible major cross-domain relationships:

**Figure 1.**
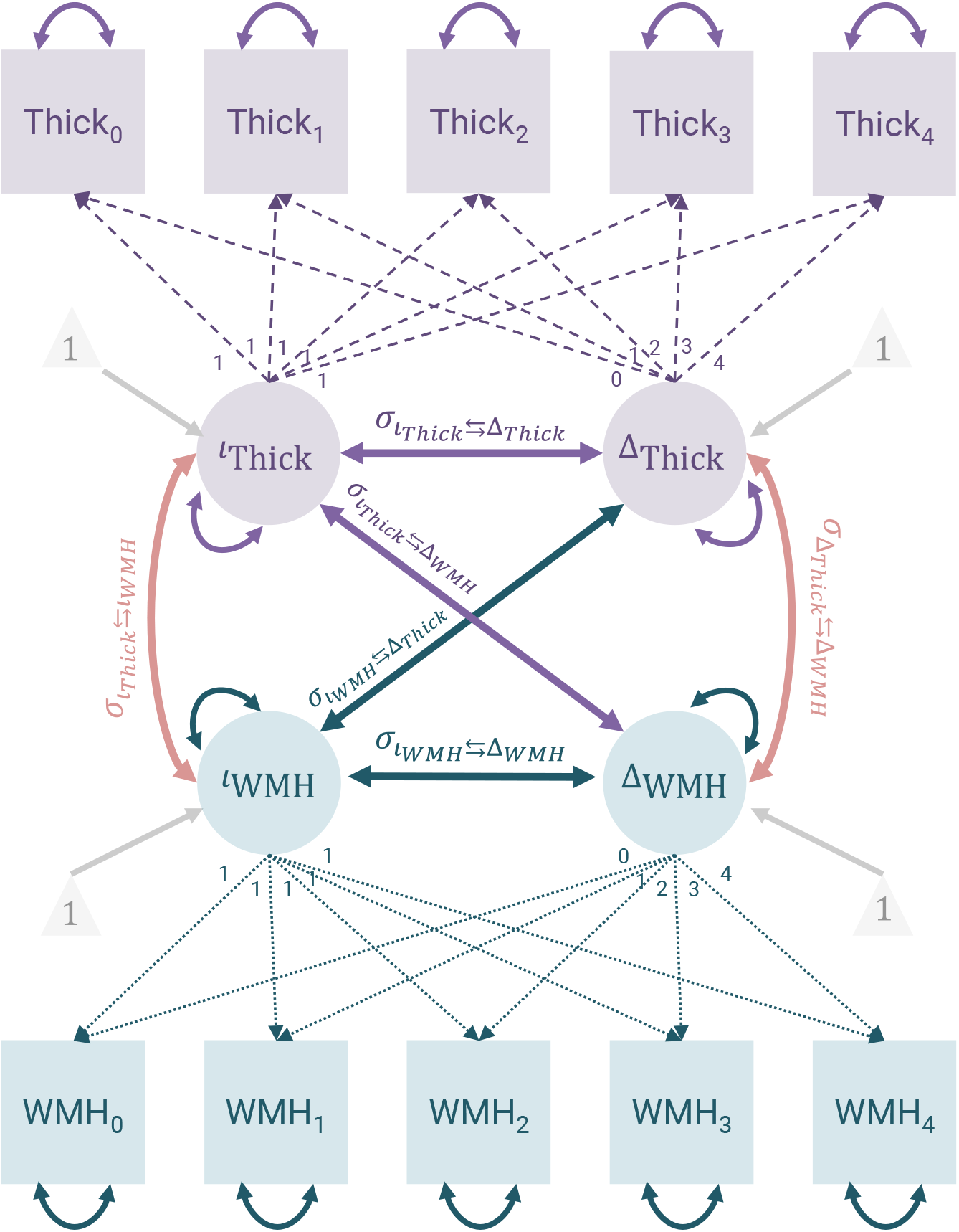
BLGCM to probe the coupling of cortical thickness and WMH over repeated measures. Illustration of the longitudinal structural equation modeling (SEM) model used on global regional/lobar level. We employed the conventional notation with squared variables indicating observed and measured variables (manifest variables) and circular ones referring to latent (unobserved) variables. Single-headed solid arrows illustrate a modelled relationship between two variables, with the arrow pointing towards the dependent variable. Single-headed dashed arrows signify a relationship between two variables, where the weight is fixed. Double-headed arrows represent the covariance (hyperparameter) between two variables. Grey triangles represent latent intercept estimates. We further adjusted latent intercepts and slopes for age, sex, years of education, total cardiovascular risk factors, and TICV. We omitted these paths for visualisation purposes.

1. *intercept-intercept covariance* 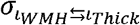: upon study entry, do individuals with larger WMH volumes have lower cortical thickness?,
2. *intercept-slope covariance* 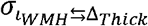 : do individuals with larger WMH volumes at study entry experience faster cortical thinning [cerebrovascular hypothesis]?,
3. *intercept-slope covariance* 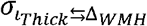: do individuals with thinner cortices at study entry exhibit a faster increase in WMH volumes [neurodegenerative hypothesis]?, and
4. *slope-slope covariance* 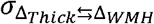: do individuals exhibiting faster WMH volume increases also undergo faster cortical thinning over time?

We also studied within-domain *intercept-slope covariances* (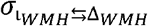 and 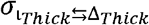) to determine whether baseline levels were associated with ongoing changes over time.

We conducted global and regional analyses to identify associations at two levels of granularity. In the global analysis—with no spatial specificity—we focused on the interrelationship between mean cortical thickness and total WMH volume. In order to elucidate potential region-specific and cross-domain relationships, we additionally examined all possible pairs of lobar WMH volumes and regional cortical thicknesses within Desikan-Killiany atlas regions. Note that our approach is similar to a mass-univariate analysis scheme, with the difference being that we investigate region-specific effects through LGCM rather than through GLM. To reduce the dimensionality and thereby improve the feasibility of our multivariate SEM analysis, we considered (corresponding) bilateral regions jointly. We used the superscripts ^Global^ and ^Regional^ to indicate the type of analysis utilised to generate the reported values. We report the completely standardised solutions and provide unstandardised solutions as Supplementary Material.

### Adjusting for covariates and confounders

We adjusted latent intercepts and slopes for effects of age, sex, years of education, total cardiovascular risk factor score, and total intracranial volume (TICV) in all models.

### Data transformation

We applied a Box-Cox transform to WMH volumes to account for potential skewness and z-scored all variables (pooled across timepoints) prior to model fitting. For the purpose of contextualisation and plotting, we back-transformed the fitted growth curve parameters afterwards.

### Model fitting

We employed the maximum likelihood robust estimator to fit the model. We used the full information maximum likelihood estimation to handle missing values. To check for compliance with the assumption of missingness at random, we tested whether missingness in one column (1: missing; 0: not missing) could be predicted from the remaining ones. In all instances, the resulting *p*-values exceeded 0.05.

Prior to model fitting and solely to ensure model fit, we used Tukey’s fences to identify and remove outliers in all data points (threshold of 1.5) [49]. We evaluated the fit of all global and regional models by analysing their root mean square error of approximation (RMSEA; values ≤ 0.05 indicate good fit), comparative fit index (CFI; values exceeding 0.95 indicate good fit), and standardised root mean residual (SRMR; values < 0.08 suggest good fit) [50]. For the sake of transparency, when discussing the models, we disclosed their convergence and compliance with the aforementioned thresholds.

### Correction for multiple comparisons

We employed the False Discovery Rate (FDR) correction [51] method to account for the issue of multiple comparisons on all region-wise analyses (Figure S1 contains the uncorrected version).

## Results

### Study participants

We included 436 cognitively unimpaired DELCODE participants with imaging data available for at least three visits (1834 MRI sessions; median age 69.70 [IQR 65.44, 74.49] years; 52.98% females; median years of education 14 [IQR 13, 17]).

### Univariate findings

#### WMH volumes increased over the course of four years

##### Model fit

All univariate LGCM on total and lobar WMH volumes converged and provided good model fit (*RMSEA* ≤ 0.05, *CFI* ≥ 0.095, *SRMR* ≤ 0.05).

##### Demographic effects on latent intercept (*t*_*WMH*_)

Baseline total WMH volumes varied significantly among individuals (variance of 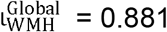, *SE* = 0.033, *Z* = 25.117, *p*-*value* < 0.001).

Total WMH volumes were larger in older individuals (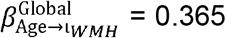, *SE* = 0.043, *Z* = 8.462, *p*-*value* < 0.001) and in those with higher total cardiovascular risk factor scores (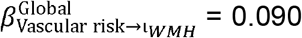, *SE* = 0.045, *Z* = 1.978, *p*-*value* = 0.048). Moreover, WMH across the temporal lobes tended to be higher in those with fewer years of education (Figure 2; 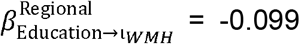, *SE* = 0.058, *Z* = -1.715, *p value* = 0.086).

**Figure 2.**
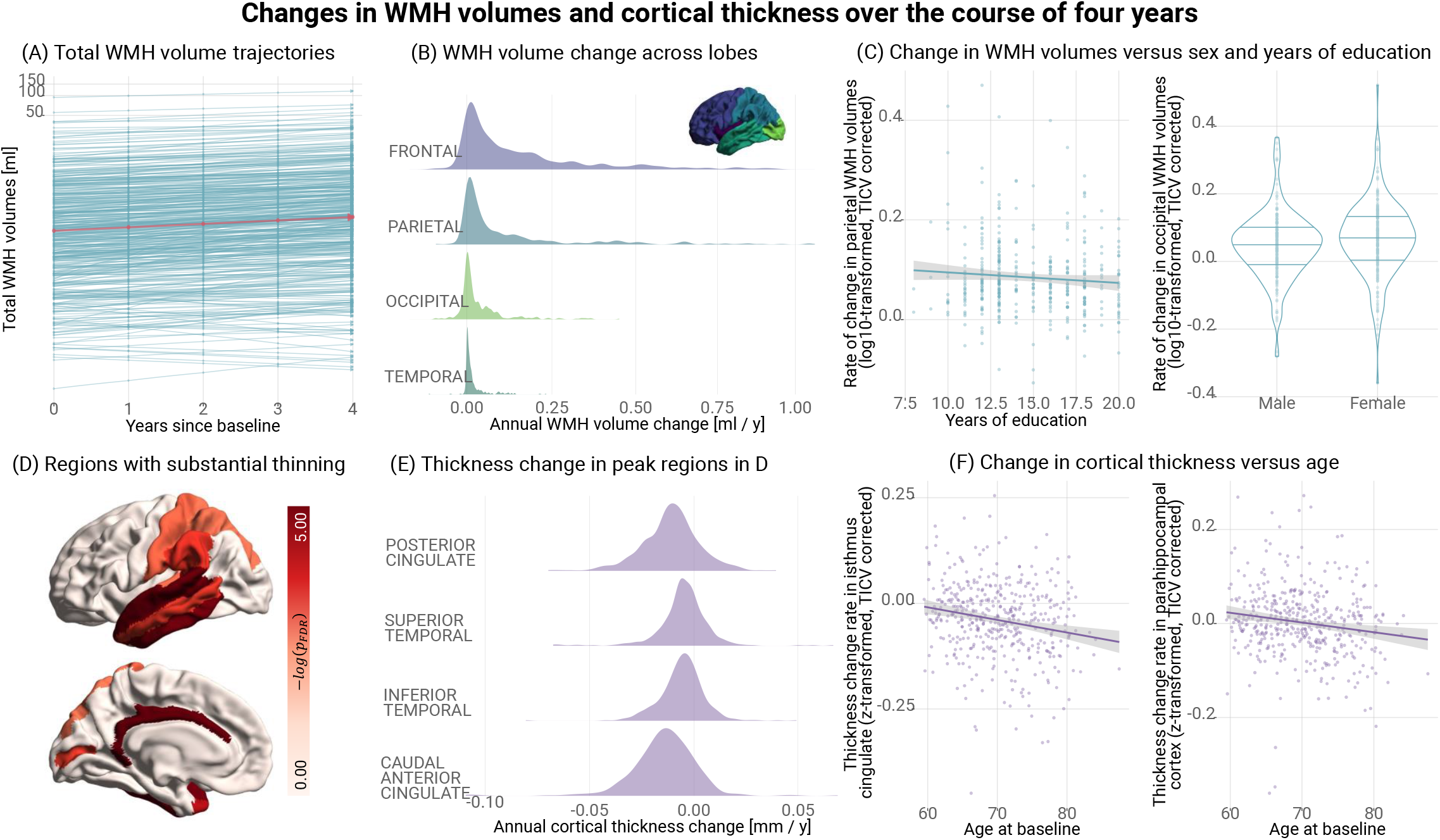
Changes in WMH volumes and cortical thickness over four years. We obtained latent intercepts and slopes for each individual through the application of univariate LGCM to WMH volumes and cortical thickness (separate models for each neuroimaging feature). We used them to compute latent growth curve parameters and predict individual trajectories, corrected for age, sex, years of education, total cardiovascular risk scores, and TICV. Prior to plotting and to enhance interpretability, we back-transformed all predicted measurements. (A) Total WMH volume trajectories, as predicted by the model. Blue lines represent the predicted trajectories and the pink one the average one. (B) Back-transformed individual factor scores of latent slopes for WMH 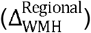, summarised in the density plots, indicate that WMH volumes generally increased over time. We adjusted density plots such that the modes attain the highest value, irrespective of the actual frequency. The rate of change varied substantially across individuals in both cases. WMH progression, from fastest to slowest, occurred in the frontal, parietal, occipital, and temporal lobes (approximated average rates of change: 0.180 [95%-CI 0.153, 0.207], 0.175 [95%-CI 0.137, 0.212], 0.034 [95%-CI 0.027, 0.042], 0.018 [IQR 0.015, 0.022] ml / year, respectively). Interestingly, while progression was predominant, a few subjects showed a clear and consistent decrease in global WMH volumes over the course of the study, especially across the occipital lobes. Visual assessment revealed that enlargement of lateral ventricles contributed to such volumetric loss. (C) Annual change in parietal and occipital WMH in relation to education and sex, respectively. (D) Regions exhibiting substantial cortical thinning over the course of the study. Both the cingulate and lateral temporal cortex underwent the most thinning (intercept of 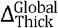 peaked in the caudal anterior cingulate cortex = -0.875, SE = 0.107, Z = -8.196, p-value < 0.001). (E) Back-transformed individual factor scores of cortical thicknesses 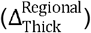 across regions experiencing the most pronounced thinning over time. Caudal anterior and posterior cingulate cortices underwent the most decline over the course of four years, with an average decrease in thickness of -0.014 [IQR -0.016, -0.013] and -0.011 [95%-CI - 0.012, -0.009] mm/year, respectively. The variability in change rates indicated significant inter-individual differences in regional cortical thinning. (F) Annual change in thickness across the isthmus cingulate and parahippocampal cortex in relation to age.

Females had larger total WMH volumes than males (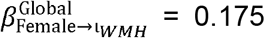, *SE* = 0.062, *Z* = 2.830, *p*-*value* = 0.005), especially across frontal brain regions (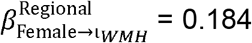, *SE* = 0.064, *Z* = 2.955, *p value* = 0.003), despite females in our sample being on average younger (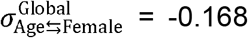, *SE* = 0.047, *Z* = -3.598, *p*-*value* < 0.001) and having lower total cardiovascular risk scores (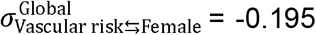, *SE* = 0.045, *Z* = -4.305, *p*-*value* < 0.001) than males. In addition, females had on average fewer years of education (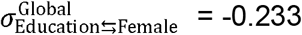, *SE* = 0.044, *Z* = - 5.346, *p*-*value* < 0.001) than males.

Demographic effects on latent slope (Δ_*WMH*_)

Across participants, total WMH volumes increased over the follow-up period of four years (Figure 2A,B; intercept of 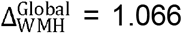, *SE* = 0.101, *Z* = 10.573, *p*-*value* < 0.001). The frontal and parietal lobes underwent the most substantial progression in WMH volumes, with an average increase of 0.180 [95%-CI 0.153, 0.207] and 0.175 [95%-CI 0.137, 0.212] ml per year, respectively. Significant WMH progression occured in occipital and temporal lobes as well (approximated average rates of change: 0.034 [95%-CI 0.027, 0.042], 0.018 [IQR 0.015, 0.022] ml per year, respectively).

A strength of the approach is the quantification of WMH progression rate variability across individuals (Figure 2B; variance of 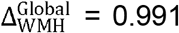, *SE* = 0.012, *Z* = 80.306, *p*-*value* < 0.001), which we found to be tied to baseline demographics and pre-existing WMH volumes. WMH progression rates in parietal regions were lower in individuals with advanced age (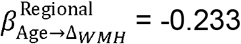, *SE* = 0.068, *Z* = -3.431, *p value* = 0.001) and with more years of education (Figure 2C; 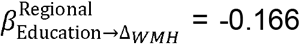, *SE* = 0.066, *Z* = -2.506, *p*-*value* = 0.036). Similarly, those with the highest initial regional WMH volumes tended to exhibit the least progression of WMH in the same region over time (parietal: 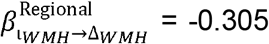, *SE* = 0.066, *Z* = -4.654, *p value* < 0.001; occipital: (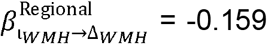, *SE* = 0.078, *Z* = -2.032, *p value* = 0.042; temporal: 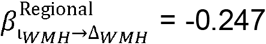, *SE* = 0.125, *Z* = -1.982, *p value* = 0.048), except in the frontal lobe (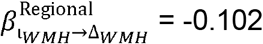, *SE* = 0.080, *Z* = -1.275, *p value* = 0.202). Sex differences in WMH progression rates were evident across the occipital lobe, with females showing faster occipital WMH progression than males (Figure 2C; 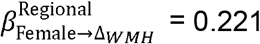, *SE* = 0.088, *Z* = 2.510, *p*-*value* = 0.012).

### Cortical thickness decreased over the course of four years

#### Model fit

All univariate LGCM fitted to cortical thickness converged and had good fit indices (*RMSEA* ≤ 0.05, *CF!* ≥ 0.095, *SRMR* ≤ 0.05).

### Demographic effects on latent intercept (*l*_*Thick*_)

Cortical thickness values were, on average, lower in individuals with advanced age (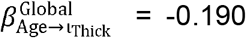, *SE* = 0.024, *Z* = -8.077, *p*-*value* < 0.001), especially within cortical regions associated with processing sensory information and orchestrating motor functions (peaks - postcentral: 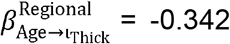, *SE* = 0.045, *Z* = -7.666, *p*-*value* < 0.001; superiortemporal: 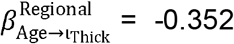, *SE* = 0.044, *Z* = -7.937, *p*-*value* < 0.001; superiorfrontal: 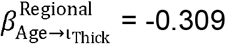, *SE* = 0.044, *Z* = -7.035, *p value* < 0.001). We did not observe sufficient evidence relating sex or cardiovascular risk factors to initial cortical measurements in the cognitively unimpaired cohort investigated here.

### Demographic effects on latent slope (Δ_*Thick*_)

The thickness of cerebral cortex generally decreased over the course of four years (Figure 2D,E; intercept of 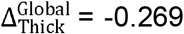, *SE* = 0.082, *Z* = -3.263, *p*-value = 0.001). Global thinning rates varied substantially among individuals (Figure 2E; variance 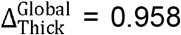, *SE* = 0.035, *Z* = 27.413, *p*-*value* < 0.001). Thinning was particularly pronounced across the cingulate and temporal cortex (Figure 2D, F; intercept of 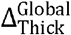 peaked in the caudal anterior cingulate cortex = -0.875, *SE* = 0.107, *Z* = - 8.196, *p*-value < 0.001). Caudal anterior and posterior cingulate cortices, for instance, underwent the most cortical thinning over the course of four years, with an average decrease in thickness of -0.014 [IQR -0.016, -0.013] and -0.011 [95%-CI - 0.012, -0.009] mm/year, respectively. Annual cortical thinning rates tended to decelerate with advance age (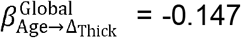, *SE* = 0.089, *Z* = -1.652, *p*-*value* = 0.098). Neither global or regional analyses hinted at an association between sex, years of education, cardiovascular risk score, and changes in cortical thickness in our cohort.

### Bivariate findings

#### Model fit

All BLGCMs also converged and had a satisfactory model fit (*RMSEA* ≤ 0.05, *CFI* ≥ 0.095, *SRMR* ≤ 0.05).

#### Individuals with larger WMH volumes have lower cortical thickness 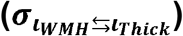

At baseline, individuals with larger WMH volumes showed lower mean cortical thickness (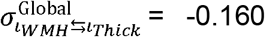, *SE* = 0.049, *Z* = -3.285, *p*-value = 0.001). Subsequent analysis of regional measurements revealed that this association was more pronounced within the same lobe (Figure 3A): frontal WMH related more strongly to the frontal cortex (peak at parstriangularis: 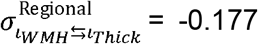, *SE* = 0.050, *Z* = -3.564, *p*-value < 0.001), occipital WMH to the occipital cortex (peak at cuneal cortex: 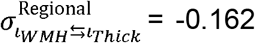, *SE* = 0.055, *Z* = -2.957, *p*-value = 0.003), and parietal WMH to the parietal cortex (peak at postcentral cortex: 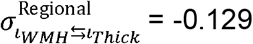, *SE* = 0.057, *Z* = -2.274, *p*-value = 0.023). However, we also observed associations across distal regions (Figure 3A), for instance, baseline frontal WMH volumes and cortical thickness in temporal brain regions, occipital WMH volumes and frontal cortical thickness.

**Figure 3.**
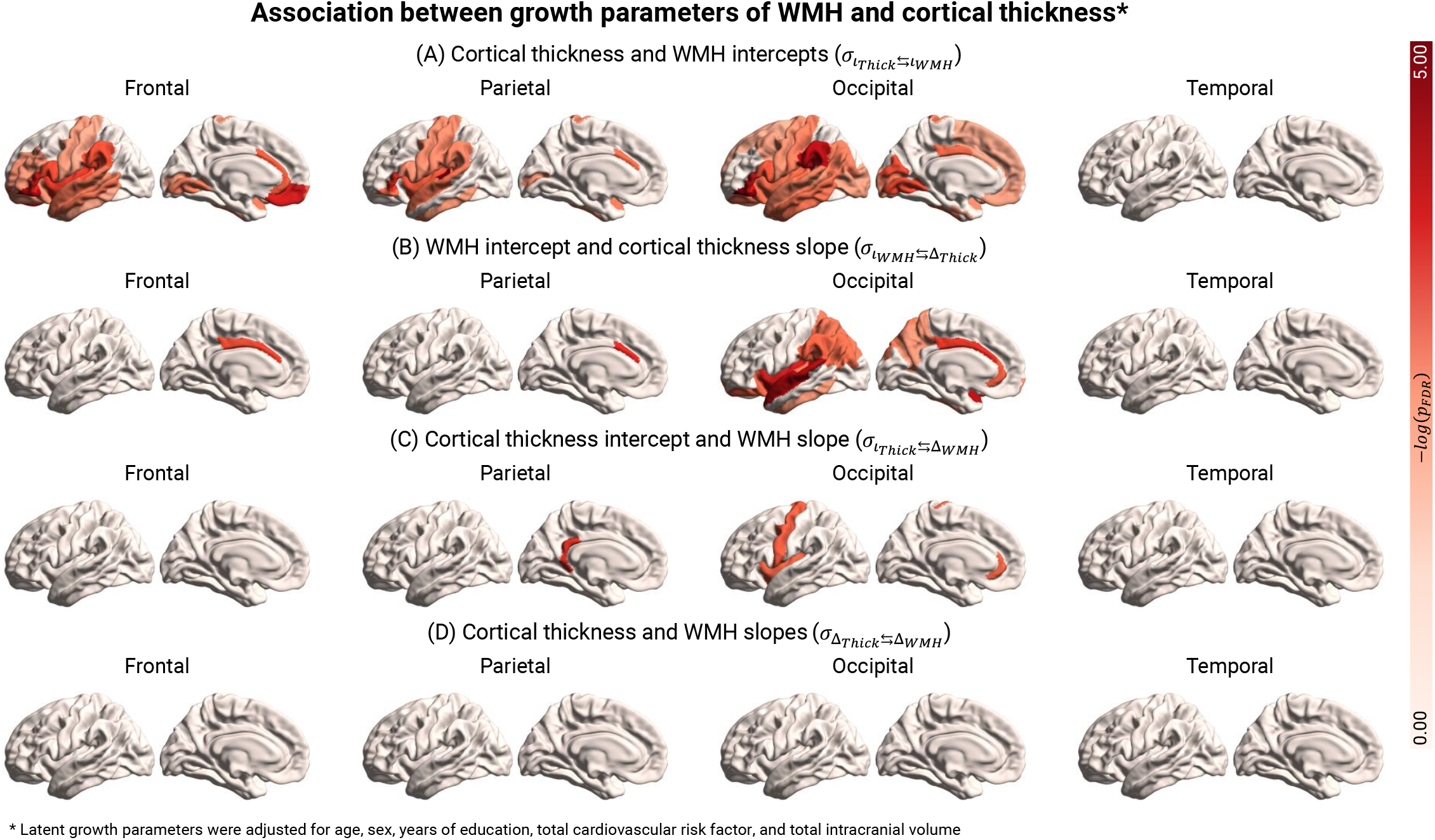
Spatiotemporal coupling between cortical thickness and WMH. We employed longitudinal BLGCMs to characterise the spatiotemporal interrelation between lobar WMH and regional cortical thickness over the span of four years—one model for each pair. We adjusted latent intercepts and slopes for age, sex, years of education, total cardiovascular risk scores, and TICV. We applied FDR correction to account for multiple comparisons. In regions highlighted in red, we found a statistically significant covariance between latent growth curve parameters after FDR correction (p_*FDR*_ < 0.05). (A; *intercept – intercept covariance*). Individuals with larger baseline WMH volumes had lower mean cortical thicknesses over the entire brain. (B; *WMH intercept – thickness slope covariance*) Individuals who underwent a steeper decrease in the thickness of their cingulate cortices had larger WMH volumes across the frontal, parietal, and occipital lobes. (C; *thickness intercept – WMH slope covariance*). Individuals with thicker cingulate cortices at baseline, particularly at the level of the isthmus, experienced a slower progression in parietal WMH volumes compared to those with thinner cortices. Also, those with initially thicker precentral, insular, and rostral anterior cingulate cortices had a slower progression in occipital WMH volumes. (D; s*lope – slope covariance*) Slope-slope covariances did not survive FDR correction. However, a rapid increase in frontal, parietal, and temporal WMH volumes was, in general, associated with accelerated cortical thinning across multiple brain regions (Figure S1). Associations with temporal WMH volumes did not survive FDR correction and, thus, we excluded them from the plot. The uncorrected version of the figure is available in Supplementary Material.

### Individuals with larger WMH volumes show faster cortical thinning 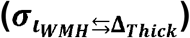

Although the association between WMH volumes at baseline and cortical thinning rates was not observed at a global scale (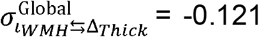, *SE* = 0.076, *Z* = - 1.595, *p*-value = 0.111), it was at a regional scale (Figure 3B). Individuals who exhibited a steeper decrease in thickness of the cingulate cortices showed larger WMH volumes across the frontal (Figure 4A.1; peak at caudal anterior: 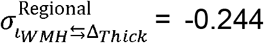, *SE* = 0.079, *Z* = -3.091, *p*-value = 0.002), parietal (peak at caudal anterior: 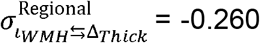, *SE* = 0.079, *Z* = -3.302, *p*-value = 0.001), and occipital lobes (peak at caudal anterior: 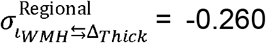, *SE* = 0.079, *Z* = - 3.302, *p*-value = 0.001). Individuals with larger occipital WMH at baseline also showed a steeper decline in thickness in adjacent cortical regions, such as the inferior parietal cortex (Figure 3B; 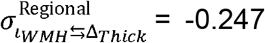, *SE* = 0.094, *Z* = -2.611, *p*-value = 0.009), but also in more remote cortical areas like the superior temporal cortex (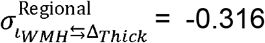, *SE* = 0.084, *Z* = -3.749, *p*-value < 0.001) and the insular cortex (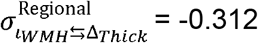, *SE* = 0.091, *Z* = -3.425, *p*-value = 0.001).

**Figure 4.**
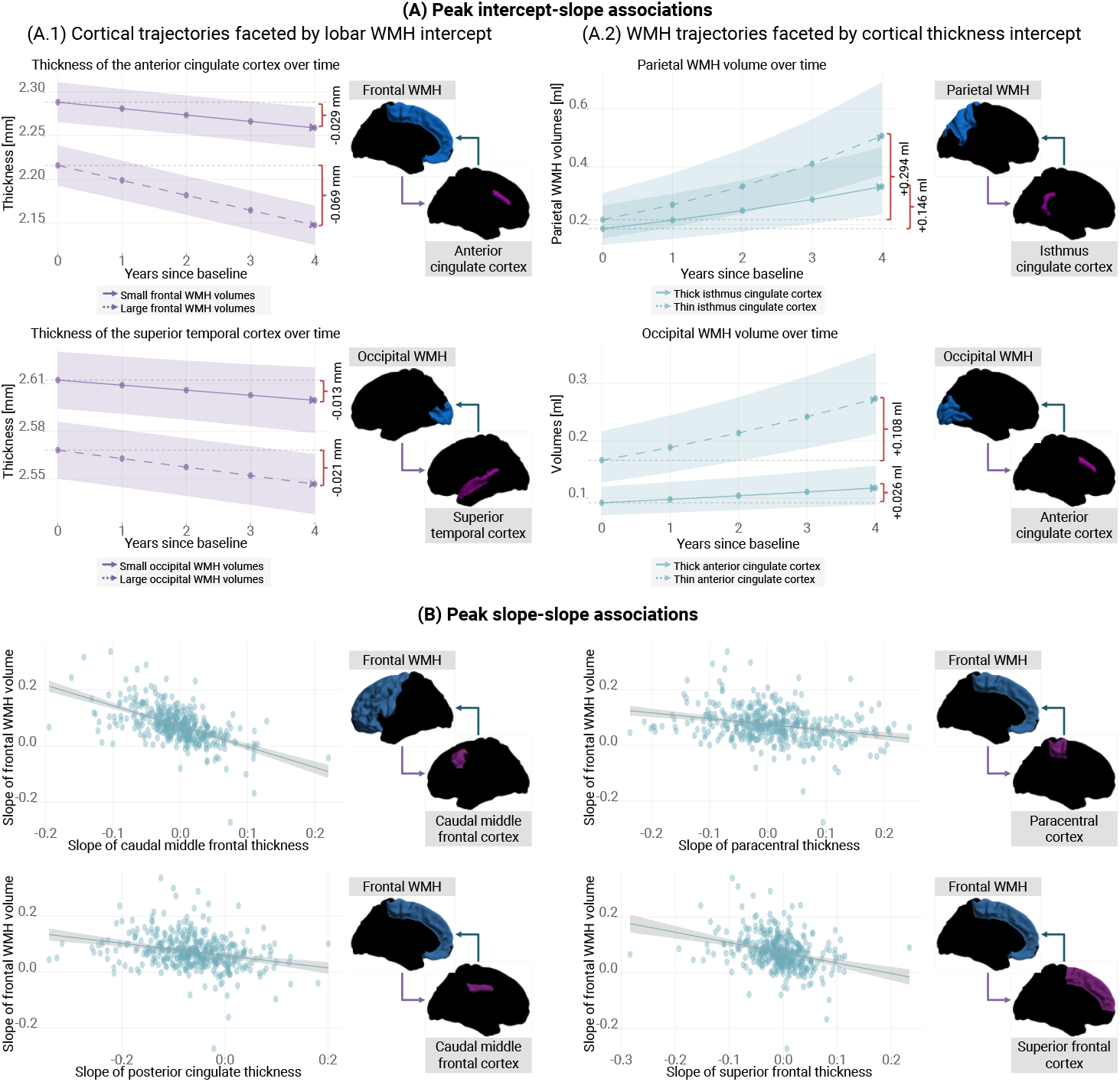
Peak intercept-slope and slope-slope asociations. (A) Average trajectories of cortical thickness and WMH trajectories over the course of four years, stratified by latent WMH and cortical thickness intercepts, respectively. We plotted here the four strongest intercept-slope associations, according to Figure 3B and C. We categorised individuals based on whether their latent intercepts were below the 33rd or above the 66th percentile, respectively. We then estimated the average trajectories for each of these groups. We back-transformed all predicted measurements to plotting for interpretability purposes. Solid and dashed lines depict the predicted average trajectories for these groups. Shadowed areas represent confidence intervals. (A.1) On average, the reduction in anterior cingulate cortex thickness among individuals with high frontal WMH volumes would be twice as large as among those with low WMH volumes (0.069 mm vs 0.029 mm in four years). Likewise, the decrease in thickness across the superior temporal cortices among individuals with high frontal WMH volumes would be approximately 1.5 times greater than that observed in those with low WMH volumes (0.021 mm vs 0.013 mm over four years). (A.2) On average, parietal WMH volume increases among individuals with thick isthmus cingulate cortices would be half as large as that observed in those with thin ones (0.146 ml vs 0.294 ml over four years). Similarly, occipital WMH volume increases across the among individuals with thin anterior cingulate cortex would be approximately four times greater than those observed in those with thick ones (0.108 ml vs 0.026 ml in four years). (B) Individuals with faster frontal WMH progression tended to have faster cortical thinning over time, especially in the caudal middle frontal, paracentral, posterior cingulate, and superior frontal cortex.

### Individuals with thinner cortices exhibit faster increases in WMH volumes 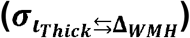

Individuals with thinner cortices at baseline tended to undergo faster WMH progression over the span of four years (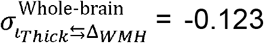, *SE* = 0.063, *Z* = - 1.964, *p*-value = 0.049). Upon closer examination of these associations, we found evidence for regional specificities (Figure 3C). Individuals with thicker cingulate cortices at baseline, particularly at the isthmus, experienced a slower progression in parietal WMH volumes compared to those with thinner cortices (Figure 3C; 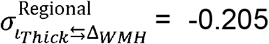, *SE* = 0.064, *Z* = -3.185, *p*-value = 0.001). Furthermore, participants with initially thicker precentral, insular, and rostral anterior cingulate cortices showed a slower progression in occipital WMH volumes (Figure 3C, 4A.2; precentral: 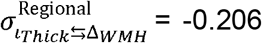, *SE* = 0.074, *Z* = -2.791, *p*-value = 0.005; insular: 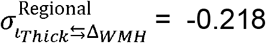, *SE* = 0.079, *Z* = -2.743, *p*-value = 0.006; rostral anterior: 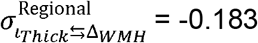, *SE* = 0.068, *Z* = -2.679, *p*-value = 0.007).

### Individuals exhibiting faster WMH volume increases also undergo faster cortical thinning over time 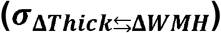

While we found no clear indication for global slope-slope associations (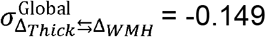, *SE* = 0.145, *Z* = -1.026, *p*-value = 0.305), we did observe that individuals with faster progression of WMH in the frontal lobe tended to undergo faster global cortical thinning over time (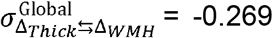, *SE* = 0.145, *Z* = - 1.861, *p*-value = 0.063). Further exploration revealed that this relationship was evident in the caudal middle frontal, paracentral, posterior cingulate, and superior frontal cortex (Figure 4B, S1; peaks – caudal middle frontal: 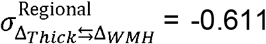, *SE* = 0.243, *Z* = -2.516, *p*-value = 0.012; paracentral: :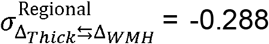, *SE* = 0.122, *Z* = -2.361, *p*-value = 0.018; posterior cingulate: : 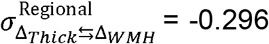, *SE* = 0.125, *Z* = -2.359, *p*-value = 0.018; superior frontal: 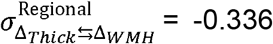, *SE* = 0.152, *Z* = -2.216, *p*-value = 0.027). Slope-slope associations were generally characterised by sparsity, with only a few regions showing statistical significance. Consequently, they collectively did not remain significant after FDR correction and should thus be interpreted with caution (FDR-corrected version: Figure 3D; uncorrected version: Figure S1).

## Discussion

We studied the interrelationships between WMH and cortical thickness over a four-year period in 436 older adults without objective cognitive impairment (1834 MRI sessions in total) using a longitudinal modelling approach. We made both methodological and clinical contributions to the ongoing efforts to understand the relationship between cerebrovascular dysfunction and neurodegeneration. First, our study demonstrates the potential of integrating surface-based morphometry and BLGCM to investigate interrelationships between neuroimaging markers over time. Second, our findings support the notion that cortical thinning and WMH progression might be mutually reinforcing processes, entangled over a four-year period in a complex and region-specific manner. Our results suggest that this coupling takes place even among individuals with a low vascular risk, given DELCODE’s inclusion and exclusion criteria.

### WMH progression

WMH generally progressed over the course of four years, reiterating that ageing is associated with WMH increase and constitutes a major risk factor for white matter pathology [2,14,15,28,52]. Progression rates were nonetheless highly subject-specific and dependent on an individual’s pre-existing white matter pathology. Parietal, occipital, and temporal WMH progression rates over four years were lower in subjects who had larger WMH volumes at the time of the baseline visit. This suggests that WMH progression may follow a non-linear trajectory, with changes occurring swiftly in its early stages, but gradually reaching a plateau. While non-linear trajectories would be conceivable at late stages of cerebral small vessel disease (CSVD) spectrum, our finding suggest that this phenomenon also applies to individuals with low-to-average WMH volumes (compared to CSVD cohorts, see [52,53]), was somewhat unexpected, albeit not a completely novel finding. The apparent non-Markovian nature of WMH progression has recently been discussed in the context of the RUN DMC study [54]. In the RUN DMC study, the total WMH burden around the age of 64 years predicted their progression over the following 11 years, with severe WMH cases progressing the most, and mild cases progressing the least [54]. Mild cases rarely progressing to severe stages [54]. Here, we found that such non-linear trajectories were observable at a study level. However, significant individual differences in WMH volume changes suggest that there are numerous other factors that were not accounted for in our study that might contribute to subject-specific progression of WMH in ageing. For example, heterogeneity of WMH volumes and progression rates could be reflective of the brain’s ability to respond to and heal from white matter injuries. By extension, heterogeneity of WMH volumes and progression rates could be reflective of past and current socioeconomic status and cardiovascular risk factors, as well as the adoption of an unhealthy lifestyle [2,54,55]. This might explain why lower years of education and greater cardiovascular risk scores were associated with higher baseline WMH volumes in our sample.

Interestingly, even though, in our study sample, males were generally older than females and had higher cardiovascular risk factor scores than females, females showed significantly greater WMH volumes at baseline compared to males even after accounting for TICV. Total WMH progression rates over the course of four years between sexes were nonetheless comparable, except in the occipital lobe, where females exhibited faster progression. For these two scenarios to be compatible, WMH would clearly need to evolve faster in females than in males in other cerebral lobes before the age of 70 years (i.e., the median age in this study). Menopause may constitute a potential explanation for this sex-specific susceptibility to WMH. A relatively recent work in the Rhineland study, a large population-based German cohort, found that while pre-menopausal women and men of similar age did not differ in WMH volumes, post-menopausal women did have significantly larger WMH volumes compared to men of similar age [56]. This finding suggests that indeed menopause and accompanying hormonal and physiological changes might be behind this sex-difference[56]. Another explanation could be that elderly women in this ageing cohort had, on average, lower educational attainment, which could also contribute to their vulnerability to CSVD. The likely multifactorial nature of this finding requires careful consideration during modelling and reporting as well as dedicated analysis shedding light on the mechanisms potentially mediating such a vulnerability.

Albeit less commonly, a small number of participants exhibited clear and consistent WMH volume regression throughout the study period, as reported in previous literature [14,57]. The most pronounced case of WMH volume regression was observed in a female participant in her 60s, with a total cardiovascular risk score of 0.0, and 15 years of education (higher education). Regression in this participant was most noticeable in occipital brain regions and could be attributed to a loss of periventricular tissue caused by a substantial enlargement of the occipital horns of the lateral ventricles over time (Figure S2). While frequently discussed in the context of a radiological or technical issue [57], our finding adds a new dimension to the current explanations for WMH regression, wherein genuine changes in one neuroimaging marker can directly influence another. This finding strongly highlights the need for multimodal longitudinal strategies to gain a more comprehensive understanding of the synergistic role of cerebrovascular and neurodegenerative processes.

### Cortical thinning

The thickness of the cerebral cortex decreased over the course of four years, corroborating that ageing also drives cortical thinning [7,58]. The rate at which thinning occurred was nonetheless subject- and region-specific. Cortical regions associated with sensory and sensorimotor functions appeared to be the earliest affected and most decreased by ageing. A decline in cortical thickness across these regions might contribute to decreased visual acuity, hearing sensitivity, and motor abilities [58–61]—all of which are expected during normal ageing, but were not examined as per the objectives of the current work. We also found substantial evidence of thinning across the cingulate cortex. This apparent ageing-related vulnerability is consistent with previous research indicating that both the caudal anterior and posterior cingulate cortex shrink during normal ageing [62]. The behavioural consequences of the rate of thinning in terms of decline in cognitive control and integrating behavioural, affective, and cognitive processes [63], remain to be elucidated.

Cortical thinning shows considerable heterogeneity across subjects. Somewhat surprisingly such inter-subject variability could not be fully explained by age, sex, years of education or cardiovascular risk factors, suggesting that other factors, e.g., genetics and lifestyle factors beyond cardiovascular risk factors [10–13], might influence cortical thinning during late life, possibly to a larger extent than demographics and established cardiovascular risk factors. Given that the rate of thinning might affect cognitive performance and activities of daily living, future research should determine the contribution of brain resilience and (modifiable) lifestyle factors to abnormal cortical thinning, as such findings could advance the development of novel interventions [64].

### Co-occurrence beyond common risk factors

Even after adjusting for shared risk factors, we found evidence for a negative correlation between the initial thickness of the cerebral cortex and the initial volume of WMH, in line with previous work [3,6,10,23–25]. While other factors may contribute to this relationship, which we did not include in our analysis (e.g., genetics and lifestyle), this observation, found in a relatively healthy sample, suggests shared underlying pathological mechanisms.

### WMH and cortical thinning

The initial volume of WMH in the brain partly explained the rate of thinning observed across multiple cortical regions over four years. This observation is consistent with the cerebrovascular hypothesis [1,65–67] and supports the notion that WMH are the visible tip of the iceberg [1], a sign of widespread rather than focal cerebrovascular and metabolic impairment [68,69].The apparent region-specific nature of the coupling between lobar WMH and regional cortical thickness raises the possibility that white matter fibres could be involved in the downstream effects of WMH. Potential secondary effects of WMH along the superior and inferior longitudinal fasciculus may, for instance, explain why those with occipital WMH would experience rapid thinning simulaneously across the temporal cortex and the cingulate cortex, including its rostral anterior regions. Mounting data indeed suggests that abnormal tissue characteristics can be found in intra- and perilesional white matter regions, but also in white matter fibres traversing WMH [1,27,69,70]. Also, cross-sectional investigations conducted in CSVD cohorts have demonstrated that cortical regions connected to incident lacunes, subcortical lacunar infarcts, and WMH through white matter fibers exhibit significantly reduced thickness than those that are not [30,65– 67]. Despite the overall compelling evidence for a contribution of WMH to cortical thinning, additional research leveraging imaging techniques like white matter tractography as well as animal models is needed to elucidate the role of white matter fibres in the long-term and remote effects of WMH in the brain.

### Cortical thickness and WMH progression

The rate of WMH volumes increase over four years was partly explained by the thickness of the cerebral cortex at study enrolment, with slower WMH progression occuring in participants with higher initial cortical thicknesses. This may indicate potentially higher brain maintanance as a mechanism of healthy ageing [71]. This relationship was particularly evident when examining WMH progression across occipital regions in relation to insular and precentral cortex thickness at baseline. The simultaneous association of insular and precentral cortex thickness with WMH development may be multifaceted. Neuronal loss in both cortical regions may be linked to lifestyle adaptations stemming from ageing that contribute to a decline in sensorimotor functions—a primary risk factor for cardiovascular disease [72]. Considering the involvement of the insular cortex in the regulation of autonomic functions, a decline in this region could also result in blood pressure dysregulation [73,74], a condition which has been extensively shown to be associated with increased progression of WMH, and with more severe manifestations of CSVD [28,54,75].

The association between baseline cortical thickness and posterior WMH progression has a fundamental ramification: it supports the spatial heterogeneity of WMH, with neurodegeneration relating more to the progression of WMH in parietal and occipital regions than in frontal ones. Since cortical neurodegeneration accelerates with the pathophysiology of Alzheimer’s disease (AD), this would explain why posterior WMH appear in subjects with minimal vascular pathology across the AD spectrum and why WMH in deep and periventricular posterior regions appear characteristics of AD [26,36,38,76]. It is also possible that an early (preclinical) increase in biomarkers indicative for AD may cause changes in the insular cortex, which then affects the cardiovascular system [73,74,77] and ultimately speeds up the progression of WMH in the brain—a possible explanation for Figure 3C. While promising, further research in other cohorts—especially with available amyloid- or tau-positron emission tomography [78]—are needed to determine how age- and AD-driven cortical neurodegeneration influences posterior WMH [76].

### WMH progression and cortical thinning

WMH progression and cortical thinning rates were associated with one another, suggesting a rather consistent and predictable relationship between the two processes, wherein changes in one marker are accompanied by corresponding changes in the other and *vice versa*. In our group of cognitively unimpaired participants, this slope-slope association was particularly evident across frontal brain regions. This pattern seems to be less confined and more widespread with advanced stages of AD, as highlighted in a recent work with autosomal dominant AD and late-onset AD [33]. Further application of our methodology to cohorts at various stages of AD could, for example, provide further information on the mechanisms underlying the simultaneous progression of both processes.

### Strengths and contextualisation

Longitudinal studies with cognitively unimpaired elderly participants exploring cross-domain associations between WMH and cortical thickness are scarce [1,4,79]. Whenever this kind of research has been done, the evidence supporting any kind of coupling has generally been lacking. In septuagenarian community-dwelling participants, Dickie et al. [4] could not find enough evidence supporting the relationship between total WMH volumes and cortical thickness of cortical grey matter structures neighbouring the Sylvian fissures over a three-year period. In a cohort of cognitively unimpaired participants, Hotz et al. [79] investigated cross-domain associations between total WMH volume and thinning of the entorhinal cortex over a duration of seven years using BLGCM. The authors found no evidence for cross-domain coupling and this absence of association was evident both at the study’s baseline and throughout its duration. Our findings are also compatible with the observation that patients with multiple sclerosis (MS), a neuroinflammatory condition primarily affecting white matter fiber tracts through demyelination, exhibit focal thinning of cortical areas [80,81].

Evidence supporting cross-domain has been, nonetheless, growing in participants symptomatic or more severe presentations of cerebrovascular [65–67,78,82] and neurodegenerative pathologies [33,78]. One potential explanation thus far for contradictory results could be the stage of dysfunction at which each participant is situated, i.e., coupling only becomes evident at advanced, symptomatic stages of cerebrovascular and neurodegenerative disease. On the other hand, as emphasised by our study, there are regional nuances to these cross-domain relationships that analyses with a lower level of granularity might fail to capture. Had we solely relied on global analyses for our study, we would have essentially been limited to observing nothing beyond the evident intercept-intercept association. This underscores the significance of employing multimodal and regional approaches to gain a more comprehensive understanding of the local and distant effects of one process on the other.

### Limitations

Our research has three main limitations. First, even though our BLGCM aligns with the data, causality remains elusive due to model equivariance. Latent change score models might be promising for further study of specific interactions over discrete time intervals [83]. The mass-univariate application of the BLGCM could be streamlined by using extended measurement models [84]. We can state, however, that our data supports a specific and partial spatiotemporal coupling between cortical neurodegeneration and cerebrovascular dysfunction. The specific circumstances that might lead to such coupling often remain undetermined and likely require the inclusion of more extensive biological parameters including complementary imaging modalities, such as diffusion tensor imaging [27,78,80]. If a Wallerian-like degeneration is responsible for the observed coupling—as also discussed in the literature [3,5,9,17,26,34,85]—there should be evidence within the white matter fibres themselves that mediate the interrelationships between cortical thickness and WMH. Second, we included a relatively healthy sample and a short time span (48 months, i.e., 4 years), which may have prevented a few cross-domain associations to become evident. The dynamics over longer time periods, as well as in other cohorts thus remain elusive, but will be a matter of future investigation. Third, we have, thus far, not assessed potential cognitive sequelae of WMH progression, cortical thinning, or their coupling in this study. Because these two processes appear to be coupled prior to any observable objective cognitive deficiencies, it could be that cognitive consequences are not detectable at this asymptomatic stage or that cognitive reserve is still able to compensate for the ongoing pathology or, as a recent study suggests, that cortical measurements predict well chronological age but not memory performance [86]. A trivariate latent change score model with WMH, cortical thickness, and cognitive performance could be used in the future to address this limitation.

## Conclusion

Our work provides longitudinal evidence that cortical thinning and WMH progression could be mutually reinforcing as opposed to parallel, disassociated processes. The coupling between these two neuroradiological features appears to be entangled prior to the onset of any detectable cognitive deficits. Our findings support the ongoing discussion on perilesional and remote impacts of WMH, but, at the same time, provide evidence for the effects of cortical neurodegeneration on white matter integrity. Comprehensive, multimodal approaches, such as the one applied in this study, have the potential to facilitate the detection of downstream damage associated with the synergistic interaction among ageing, CSVD, and neurodegeneration in the brain.

## Data Availability

The datasets used and analysed during the current study are available from the corresponding author on reasonable request.

## Abbreviations

AD: Alzheimer’s Disease
BLGCM: Bivariate Latent Growth Curve Model
CERAD: Consortium to Establish a Registry for AD
CFI: Comparative Fit Index
CI: Confidence Interval
CSF: Cerebrospinal Fluid
CSVD: Cerebral Small Vessel Disease
DELCODE: DZNE Longitudinal Cognitive Impairment and Dementia Study
DZNE: German Centre for Neurodegenerative Diseases
FDR: False Discovery Rate
HCP: Human Connectome Project
LGCM: Latent Growth Curve Model
MRI: Magnetic Resonance Imaging
MS: Multiple Sclerosis
RMSEA: Root Mean Square Error of Approximation
SE: Standard Error
SEM: Structural Equation Model
SRMR: Standardised Root Mean Residual
TICV: Total Intracranial Volume
WMH: White Matter Hyperintensities

## Declarations

### Ethics approval and consent to participate

The study protocol was approved by the ethical committees of the medical faculties of all participating sites: the ethical committees of Berlin (Charite, University Medicine), Bonn, Cologne, Goettingen, Magdeburg, Munich (Ludwig-Maximilians-University), Rostock, and Tuebingen. The process was led and coordinated by the ethical committee of the medical faculty of the University of Bonn. All commitees gave ethical approval for this work. All participants provided their written informed consent before inclusion in the study. DELCODE is retrospectively registered at the German Clinical Trials Register (DRKS00007966, 04/05/2015). The DELCODE study was conducted in accordance with the Declaration of Helsinki.

### Consent for publication

Not applicable.

### Author’s contributions

Conceptualisation: JB, IM, GZ. Methodology: JB, IM, GZ. Software: JB, IM, GZ. Formal analysis: JB. Data Acquisition: OP, JHR, SDF, JP, EJS, SA, ASc, KF, JW, BHS, FJ, AR, WG, EII, KB, DJ, ME, RP, BSR, ST, IK, CL, SSo, ASp, AE, FL, PD, SH, KS, ED. Image processing: JB, RY. Visualisation: JB. Image analysis and modelling: JB, GZ. Investigation: JB, GZ. Supervision: GZ, ED. Project administration: GZ. Funding acquisition: ED. Resources: GZ, ED. Writing original draft preparation: JB, GZ. Writing – review and editing: All authors. All authors read and approved the final manuscript.

## Acknowledgements

We would like to express our gratitude to all DELCODE study participants. We also thank the Max-Delbrück-centrum für Molekulare Medizin in der Helmholtz-Gemeinschaft (MDC), Freie Universität Berlin Center for Cognitive Neuroscience Berlin (CCNB), Bernstein Center für Computional Neuroscience Berlin, Universitätsmedizin Göttingen Core Facility MR-Research in Neurosciences, Institut für Klinische Radiologie Klinikum der Universität München, and Universitätsklinikum Tübingen MR-Forschungszentrum.

## References

1. Ter Telgte A, Van Leijsen EMC, Wiegertjes K, Klijn CJM, Tuladhar AM, De Leeuw FE. Cerebral small vessel disease: From a focal to a global perspective. Nat Rev Neurol. 2018;14:387–98.

2. Duering M, Biessels GJ, Brodtmann A, Chen C, Cordonnier C, de Leeuw F-E, et al. Neuroimaging standards for research into small vessel disease-advances since 2013. Lancet Neurol. 2023;4422:2–4.

3. Appelman APA, Exalto LG, Van Der Graaf Y, Biessels GJ, Mali Wptm, Geerlings MI. White matter lesions and brain atrophy: More than shared risk factors? A systematic review. Cerebrovascular Diseases. 2009;28:227–42.

4. Dickie DA, Karama S, Ritchie SJ, Cox SR, Sakka E, Royle NA, et al. Progression of White Matter Disease and Cortical Thinning Are Not Related in Older Community-Dwelling Subjects. Stroke. 2016;47:410–6.

5. Fiford CM, Manning EN, Bartlett JW, Cash DM, Malone IB, Ridgway GR, et al. White matter hyperintensities are associated with disproportionate progressive hippocampal atrophy. Hippocampus. 2017;27:249–62.

6. Lambert C, Benjamin P, Zeestraten E, Lawrence AJ, Barrick TR, Markus HS. Longitudinal patterns of leukoaraiosis and brain atrophy in symptomatic small vessel disease. Brain. 2016;139:1136–51.

7. Bethlehem RAI, Seidlitz J, White SR, Vogel JW, Anderson KM, Adamson C, et al. Brain charts for the human lifespan. Nature. 2022;604:525–33.

8. Narvacan K, Treit S, Camicioli R, Martin W, Beaulieu C. Evolution of deep gray matter volume across the human lifespan. Hum Brain Mapp. 2017;38:3771–90.

9. Jouvent E, Viswanathan A, Chabriat H. Views and Reviews Cerebral Atrophy in Cerebrovascular Disorders. 2009;213–8.

10. Enzinger C, Fazekas F, Matthews PM, Ropele S, Schmidt H, Smith S, et al. Risk factors for progression of brain atrophy in aging: Six-year follow-up of normal subjects. Neurology. 2005;64:1704–11.

11. Carmelli D, Swan GE, Reed T, Wolf PA, Miller BL, DeCarli C. Midlife cardiovascular risk factors and brain morphology in identical older male twins. Neurology. 1999;52:1119–24.

12. Ong M, Foo H, Chander RJ, Wen MC, Au WL, Sitoh YY, et al. Influence of diabetes mellitus on longitudinal atrophy and cognition in Parkinson’s disease. J Neurol Sci [Internet]. 2017;377:122–6. Available from: 10.1016/j.jns.2017.04.010

13. Xu J, Li Y, Lin H, Sinha R, Potenza MN. Body mass index correlates negatively with white matter integrity in the fornix and corpus callosum: A diffusion tensor imaging study. Hum Brain Mapp. 2013;34:1044–52.

14. Jochems ACC, Arteaga C, Chappell F, Ritakari T, Hooley M, Doubal F, et al. Longitudinal Changes of White Matter Hyperintensities in Sporadic Small Vessel Disease: A Systematic Review and Meta-analysis. Neurology. 2022;99:E2454–63.

15. Wardlaw JM, Valdés Hernández MC, Muñoz-Maniega S. What are white matter hyperintensities made of? Relevance to vascular cognitive impairment. J Am Heart Assoc. 2015;4:001140.

16. Behl C. Apoptosis and Alzheimer ‘ s disease Review. Journal of Neural Transmissio. 2000;1325–44.

17. Nasrabady SE, Rizvi B, Goldman JE, Brickman AM. White matter changes in Alzheimer’s disease: a focus on myelin and oligodendrocytes. Acta Neuropathol Commun. 2018;6:22.

18. Obulesu M, Lakshmi MJ. Apoptosis in Alzheimer’s Disease: An Understanding of the Physiology, Pathology and Therapeutic Avenues. Neurochem Res. 2014;39:2301–12.

19. Jouvent E, Mangin JF, Duchesnay E, Porcher R, Düring M, Mewald Y, et al. Longitudinal changes of cortical morphology in CADASIL. Neurobiol Aging [Internet]. 2012;33:1002.e29-1002.e36. Available from: 10.1016/j.neurobiolaging.2011.09.013

20. Brown WR, Moody DM, Thore CR, Challa VR. Apoptosis in leukoaraiosis. American Journal of Neuroradiology. 2000;21:79–82.

21. Wen W, Sachdev PS, Chen X, Anstey K. Gray matter reduction is correlated with white matter hyperintensity volume: A voxel-based morphometric study in a large epidemiological sample. Neuroimage. 2006;29:1031–9.

22. Kim SE, Kim HJ, Jang H, Weiner MW, DeCarli C, Na DL, et al. Interaction between Alzheimer’s Disease and Cerebral Small Vessel Disease: A Review Focused on Neuroimaging Markers. Int J Mol Sci. MDPI; 2022.

23. Dadar M, Manera AL, Ducharme S, Collins DL. White matter hyperintensities are associated with grey matter atrophy and cognitive decline in Alzheimer’s disease and frontotemporal dementia. Neurobiol Aging [Internet]. 2022;111:54–63. Available from: 10.1016/j.neurobiolaging.2021.11.007

24. Rizvi B, Lao PJ, Chesebro AG, Dworkin JD, Amarante E, Beato JM, et al. Association of Regional White Matter Hyperintensities with Longitudinal Alzheimer-Like Pattern of Neurodegeneration in Older Adults. JAMA Netw Open. 2021;1–13.

25. Rizvi B, Sathishkumar M, Kim S, Márquez F, Granger SJ, Larson MS, et al. Posterior white matter hyperintensities are associated with reduced medial temporal lobe subregional integrity and long-term memory in older adults. Neuroimage Clin. 2023;37.

26. Garnier-crussard A, Krolak-salmon P, Garnier-crussard A, Cotton F, Krolak-salmon P. White matter hyperintensities in Alzheimer ‘ s disease?: Beyond vascular contribution. Alzheimers Dement. 2023;

27. Dalby RB, Eskildsen SF, Videbech P, Frandsen J, Mouridsen K, Sørensen L, et al. Oxygenation differs among white matter hyperintensities, intersected fiber tracts and unaffected white matter. Brain Commun. 2019;1.

28. Ungvari Z, Toth P, Tarantini S, Prodan CI, Sorond F, Merkely B, et al. Hypertension-induced cognitive impairment: from pathophysiology to public health. Nat Rev Nephrol. 2021;17:639–54.

29. van Veluw SJ, Arfanakis K, Schneider JA. Neuropathology of Vascular Brain Health: Insights from Ex Vivo Magnetic Resonance Imaging-Histopathology Studies in Cerebral Small Vessel Disease. Stroke. 2022;53:404–15.

30. Mayer C, Frey BM, Schlemm E, Petersen M, Engelke K, Hanning U, et al. Linking cortical atrophy to white matter hyperintensities of presumed vascular origin. Journal of Cerebral Blood Flow and Metabolism. 2021;41:1682–91.

31. McAleese KE, Firbank M, Dey M, Colloby SJ, Walker L, Johnson M, et al. Cortical tau load is associated with white matter hyperintensities. Acta Neuropathol Commun. 2015;3:60.

32. McAleese KE, Walker L, Graham S, Moya ELJ, Johnson M, Erskine D, et al. Parietal white matter lesions in Alzheimer’s disease are associated with cortical neurodegenerative pathology, but not with small vessel disease. Acta Neuropathol. 2017;134:459–73.

33. Shirzadi Z, Schultz SA, Yau W-YW, Joseph-Mathurin N, Fitzpatrick CD, Levin R, et al. Etiology of White Matter Hyperintensities in Autosomal Dominant and Sporadic Alzheimer Disease. JAMA Neurol [Internet]. 2023; Available from: https://jamanetwork.com/journals/jamaneurology/fullarticle/2810315

34. Salvadores N, Gerónimo-Olvera C, Court FA. Axonal Degeneration in AD: The Contribution of Aβ and Tau. Front Aging Neurosci. Frontiers Media S.A.; 2020.

35. Bernal J, Schreiber S, Menze I, Ostendorf A, Pfister M, Geisendörfer J, et al. Arterial hypertension and β-amyloid accumulation have spatially overlapping effects on posterior white matter hyperintensity volume: a cross-sectional study. Alzheimers Res Ther. 2023;15.

36. Alber J, Alladi S, Bae HJ, Barton DA, Beckett LA, Bell JM, et al. White matter hyperintensities in vascular contributions to cognitive impairment and dementia (VCID): Knowledge gaps and opportunities. Alzheimer’s and Dementia: Translational Research and Clinical Interventions. 2019;5:107–17.

37. Garnier-Crussard A, Bougacha S, Wirth M, Dautricourt S, Sherif S, Landeau B, et al. White matter hyperintensity topography in Alzheimer’s disease and links to cognition. Alzheimer’s and Dementia. 2022;18:422–33.

38. Pålhaugen L, Sudre CH, Tecelao S, Nakling A, Almdahl IS, Kalheim LF, et al. Brain amyloid and vascular risk are related to distinct white matter hyperintensity patterns. Journal of Cerebral Blood Flow and Metabolism. 2021;41:1162–74.

39. Jessen F, Spottke A, Boecker H, Brosseron F, Buerger K, Catak C, et al. Design and first baseline data of the DZNE multicenter observational study on predementia Alzheimer’s disease (DELCODE). Alzheimers Res Ther. 2018;10:1–10.

40. Gaser C, Dahnke R, Kurth K, Luders E, Alzheimer’s Disease Neuroimaging Initiative. A computational Anatomy Toolbox for the Analysis of Structural MRI Data. bioRxiv. 2022[1–37.

41. Desikan RS, Ségonne F, Fischl B, Quinn BT, Dickerson BC, Blacker D, et al. An automated labeling system for subdividing the human cerebral cortex on MRI scans into gyral based regions of interest. Neuroimage. 2006;31:968–80.

42. Yushkevich PA, Pluta J, Wang H, Wisse LEM, Das S, Wolk D. IC-P-174: Fast Automatic Segmentation of Hippocampal Subfields and Medial Temporal Lobe Subregions In 3 Tesla and 7 Tesla T2-Weighted MRI. Alzheimer’s & Dementia. 2016;12.

43. Isensee F, Schell M, Pflueger I, Brugnara G, Bonekamp D, Neuberger U, et al. Automated brain extraction of multisequence MRI using artificial neural networks. Hum Brain Mapp. 2019;40:4952–64.

44. Wiltgen T, McGinnis J, Schlaeger S, Kofler F, Voon CC, Berthele A, et al. LST-AI: A deep learning ensemble for accurate MS lesion segmentation. Neuroimage Clin. 2024;42.

45. Joshi AA, Choi S, Liu Y, Chong M, Sonkar G, Gonzalez-Martinez J, et al. A hybrid high-resolution anatomical MRI atlas with sub-parcellation of cortical gyri using resting fMRI. J Neurosci Methods. 2022;374.

46. Larivière S, Paquola C, Park B yong, Royer J, Wang Y, Benkarim O, et al. The ENIGMA Toolbox: multiscale neural contextualization of multisite neuroimaging datasets. Nat Methods. Nature Research; 2021. p. 698–700.

47. McArdle JJ, Nesselroade John R. Using multivariate data to structure developmental change.” Life-span developmental psychology: Methodological contributions. In: Cohen SH, Reese HW, editors. Life-Span Developmental Psychology: Methodological Contributions. 1st ed. New York: Psychology Press; 1994. p. 223–67.

48. Muniz-Terrera G, Robitaille A, Kelly A, Johansson B, Hofer S, Piccinin A. Latent growth models matched to research questions to answer questions about dynamics of change in multiple processes. J Clin Epidemiol. 2017;82:158–66.

49. Tukey JW. Exploratory data analysis. Addison-Wesley Pub. Co.; 1977.

50. Hu LT, Bentler PM. Cutoff criteria for fit indexes in covariance structure analysis: Conventional criteria versus new alternatives. Structural Equation Modeling. 1999;6:1–55.

51. Benjamini Y, Hochberg Y. Controlling the false discovery rate: a practical and powerful approach to multiple testing. Journal of the royal statistical society Series B (Methodological). 1995;289–300.

52. Van Leijsen EMC, Van Uden IWM, Ghafoorian M, Bergkamp MI, Lohner V, Kooijmans ECM, et al. Nonlinear temporal dynamics of cerebral small vessel disease. Neurology. 2017;89:1569–77.

53. Keuss SE, Coath W, Nicholas JM, Poole T, Barnes J, Cash DM, et al. Associations of β-Amyloid and Vascular Burden with Rates of Neurodegeneration in Cognitively Normal Members of the 1946 British Birth Cohort. Neurology. 2022;99:E129–41.

54. Cai M, Jacob MA, Van Loenen MR, Bergkamp M, Marques J, Norris DG, et al. Determinants and Temporal Dynamics of Cerebral Small Vessel Disease: 14-Year Follow-Up. Stroke. 2022;53:2789–98.

55. Wardlaw JM, Smith C, Dichgans M. Small vessel disease: mechanisms and clinical implications. Lancet Neurol. 2019;18:684–96.

56. Lohner V, Pehlivan G, Sanroma G, Miloschewski A, Schirmer MD, Stöcker T, et al. Relation Between Sex, Menopause, and White Matter Hyperintensities: The Rhineland Study. Neurology. 2022;99:E935–43.

57. Brown RB, Tozer DJ, Egle M, Tuladhar AM, de Leeuw FE, Markus HS. How often does white matter hyperintensity volume regress in cerebral small vessel disease? International Journal of Stroke. 2023;00.

58. Salat DH, Buckner RL, Snyder AZ, Greve DN, Desikan RSR, Busa E, et al. Thinning of the Cerebral Cortex in Aging. Cerebral Cortex [Internet]. 2004;14:721–30. Available from: 10.1093/cercor/bhh032

59. Cardin V. Effects of aging and adult-onset hearing loss on cortical auditory regions. Front Neurosci. Frontiers Media S.A.; 2016.

60. Li KZH, Lindenberger U. Relations between aging sensory/sensorimotor and cognitive functions. Neurosci Biobehav Rev [Internet]. 2002;26:777–83. Available from: https://www.elsevier.com/locate/neubiorev

61. Schneider BA, Pichora-Fuller MK. Implications of perceptual deterioration for cognitive aging research. The handbook of aging and cognition, 2nd ed. Mahwah, NJ, US: Lawrence Erlbaum Associates Publishers; 2000. p. 155–219.

62. Mann SL, Hazlett EA, Byne W, Hof PR, Buchsbaum MS, Cohen BH, et al. Anterior and posterior cingulate cortex volume in healthy adults: Effects of aging and gender differences. Brain Res. 2011;1401:18–29.

63. Shackman AJ, Salomons T V., Slagter HA, Fox AS, Winter JJ, Davidson RJ. The integration of negative affect, pain and cognitive control in the cingulate cortex. Nat Rev Neurosci. 2011. p. 154–67.

64. Schwarck S, Voelkle MC, Becke A, Busse N, Glanz W, Ziegler G. Interplay of physical and cognitive performance using hierarchical continuous-time dynamic modelling and a 2 dual-task training regime in Alzheimer’s patients. Available from: 10.1101/2022.12.14.22283428

65. Duering M, Righart R, Wollenweber FA, Zietemann V, Gesierich B, Dichgans M. Acute infarcts cause focal thinning in remote cortex via degeneration of connecting fiber tracts. Neurology. 2015;84:1685–92.

66. Duering M, Righart R, Csanadi E, Jouvent E, Herve D, Chabriat H, et al. Incident subcortical infarcts induce focal thinning in connected cortical regions. Neurology. 2012;79:2025–8.

67. Li H, Jacob MA, Cai M, Duering M, Chamberland M, Norris DG, et al. Regional cortical thinning, demyelination, and iron loss in cerebral small vessel disease. Brain. 2023;

68. Jiaerken Y, Luo X, Yu X, Huang P, Xu X, Zhang M. Microstructural and metabolic changes in the longitudinal progression of white matter hyperintensities. Journal of Cerebral Blood Flow and Metabolism. 2019;39:1613–22.

69. Reginold W, Sam K, Poublanc J, Fisher J, Crawley A, Mikulis DJ. Impact of white matter hyperintensities on surrounding white matter tracts. Neuroradiology. 2018;60:933–44.

70. Wardlaw JM, Makin SJ, Valdés Hernández MC, Armitage PA, Heye AK, Chappell FM, et al. Blood-brain barrier failure as a core mechanism in cerebral small vessel disease and dementia: evidence from a cohort study. Alzheimer’s and Dementia. 2017;13:634–43.

71. Cabeza R, Albert M, Belleville S, Craik FIM, Duarte A, Grady CL, et al. Maintenance, reserve and compensation: the cognitive neuroscience of healthy ageing. Nat Rev Neurosci. Nature Publishing Group; 2018. p. 701–10.

72. Li J, Siegrist J. Physical activity and risk of cardiovascular disease-a meta-analysis of prospective cohort studies. Int J Environ Res Public Health. MDPI; 2012. p. 391–407.

73. Benarroch EE. Insular cortex: Functional complexity and clinical correlations. Neurology. 2019;93:932–8.

74. Gogolla N. The insular cortex. Current Biology. Cell Press; 2017. p. R580–6.

75. Duering M, Biessels GJ, Brodtmann A, Chen C, Cordonnier C, de Leeuw F-E, et al. Neuroimaging standards for research into small vessel disease—advances since 2013. Lancet Neurol [Internet]. 2023;22:602–18. Available from: https://linkinghub.elsevier.com/retrieve/pii/S147444222300131X

76. Desmarais P, Gao AF, Lanctôt K, Rogaeva E, Ramirez J, Herrmann N, et al. White matter hyperintensities in autopsy-confirmed frontotemporal lobar degeneration and Alzheimer’s disease. Alzheimers Res Ther. 2021;13:1–16.

77. Kitamura J, Nagai M, Ueno H, Ohshita T, Kikumoto M, Toko M, et al. The Insular Cortex, Alzheimer Disease Pathology, and Their Effects on Blood Pressure Variability [Internet]. 2020. Available from: https://www.alzheimerjournal.com

78. Zhang J, Chen H, Wang J, Huang Q, Xu X, Wang W, et al. Linking white matter hyperintensities to regional cortical thinning, amyloid deposition, and synaptic density loss in Alzheimer’s disease. Alzheimer’s & Dementia [Internet]. 2024; Available from: https://alz-journals.onlinelibrary.wiley.com/doi/10.1002/alz.13845

79. Hotz I, Deschwanden PF, Mérillat S, Jäncke L. Associations between white matter hyperintensities, lacunes, entorhinal cortex thickness, declarative memory and leisure activity in cognitively healthy older adults: A 7-year study. Neuroimage. 2023;284.

80. Bussas M, Grahl S, Pongratz V, Berthele A, Gasperi C, Andlauer T, et al. Gray matter atrophy in relapsing-remitting multiple sclerosis is associated with white matter lesions in connecting fibers. Multiple Sclerosis Journal. 2022;28:900–9.

81. Sailer M, Fischl B, Salat D, Tempelmann C, Schönfeld MA, Busa E, et al. Focal thinning of the cerebral cortex in multiple sclerosis. Brain. 2003;126:1734–44.

82. Kim SJ, Lee DK, Jang YK, Jang H, Kim SE, Cho SH, et al. The effects of longitudinal white matter hyperintensity change on cognitive decline and cortical thinning over three years. J Clin Med. 2020;9:1–13.

83. Kievit RA, Brandmaier AM, Ziegler G, van Harmelen AL, de Mooij SMM, Moutoussis M, et al. Developmental cognitive neuroscience using latent change score models: A tutorial and applications. Dev Cogn Neurosci [Internet]. 2018;33:99–117. Available from: 10.1016/j.dcn.2017.11.007

84. Wang L, Lyu X, Zhang Z, Li L. High-dimensional Response Growth Curve Modeling for Longitudinal Neuroimaging Analysis. ArXiv [Internet]. 2023;1–30. Available from: http://arxiv.org/abs/2305.15751

85. Reginold W, Itorralba J, Luedke AC, Fernandez-Ruiz J, Reginold J, Islam O, et al. Tractography at 3T MRI of corpus callosum tracts crossing white matter hyperintensities. American Journal of Neuroradiology. 2016;37:1617–22.

86. Soch J, Richter A, Kizilirmak JM, Schütze H, Feldhoff H, Fischer L, et al. Structural and Functional MRI Data Differentially Predict Chronological Age and Behavioral Memory Performance. eNeuro. 2022;9.

