## Supplementary Material for "Longitudinal evidence for a mutually reinforcing relationship between white matter hyperintensities and cortical thickness in cognitively unimpaired older adults"

Supplementary Figures

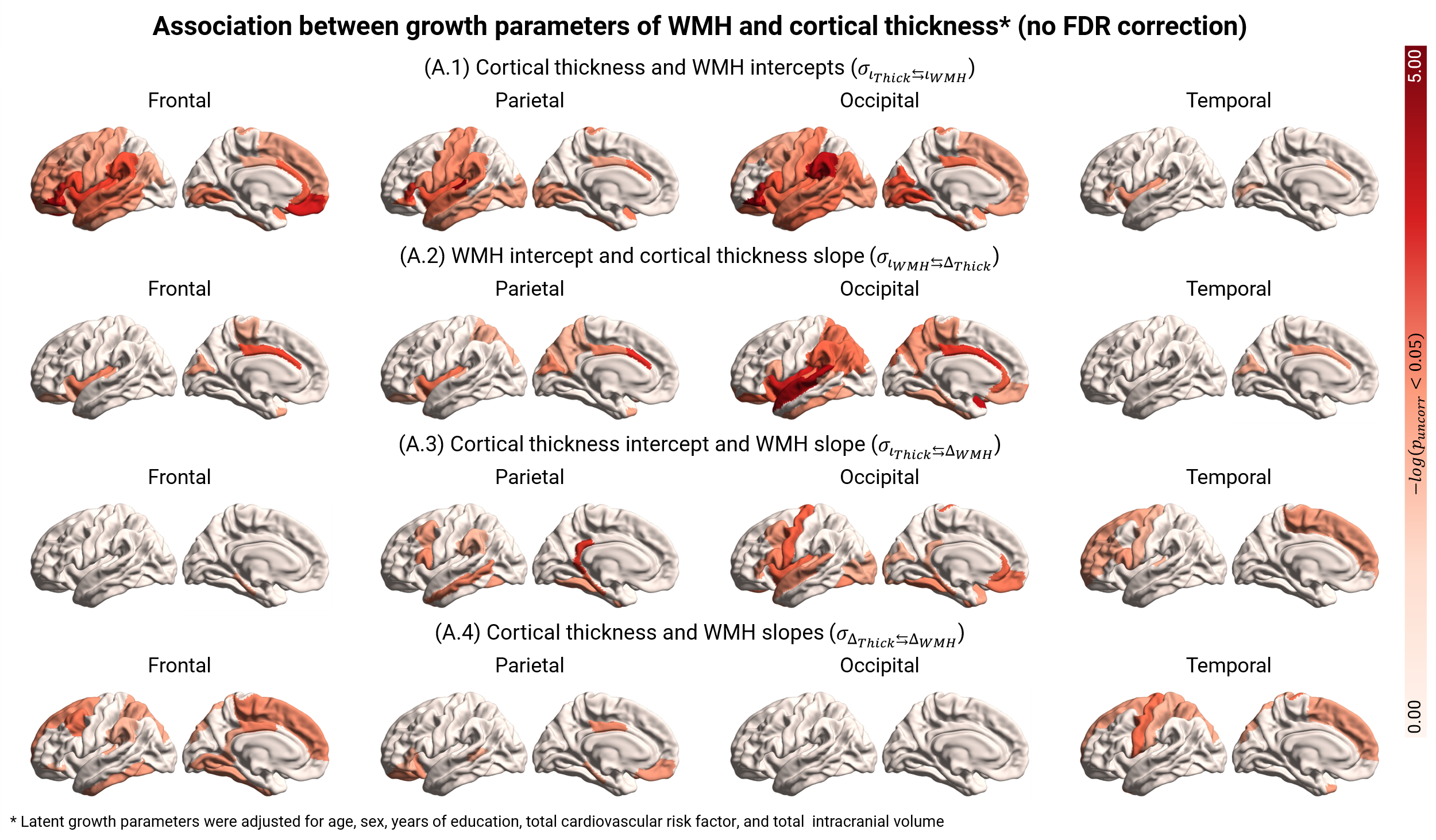

**Figure S1. Spatiotemporal coupling between cortical thickness and WMH.** We employed BLGCMs to characterise the spatiotemporal interrelation between WMH and cortical thickness over the span of four years. Latent intercepts and slopes were adjusted for age, sex, years of education, total cardiovascular risk scores, and TICV. Unlike in Figure 3, this image contains uncorrected results. Regions highlighted in red denote those where we found a statistically significant covariance between latent growth curve parameters.

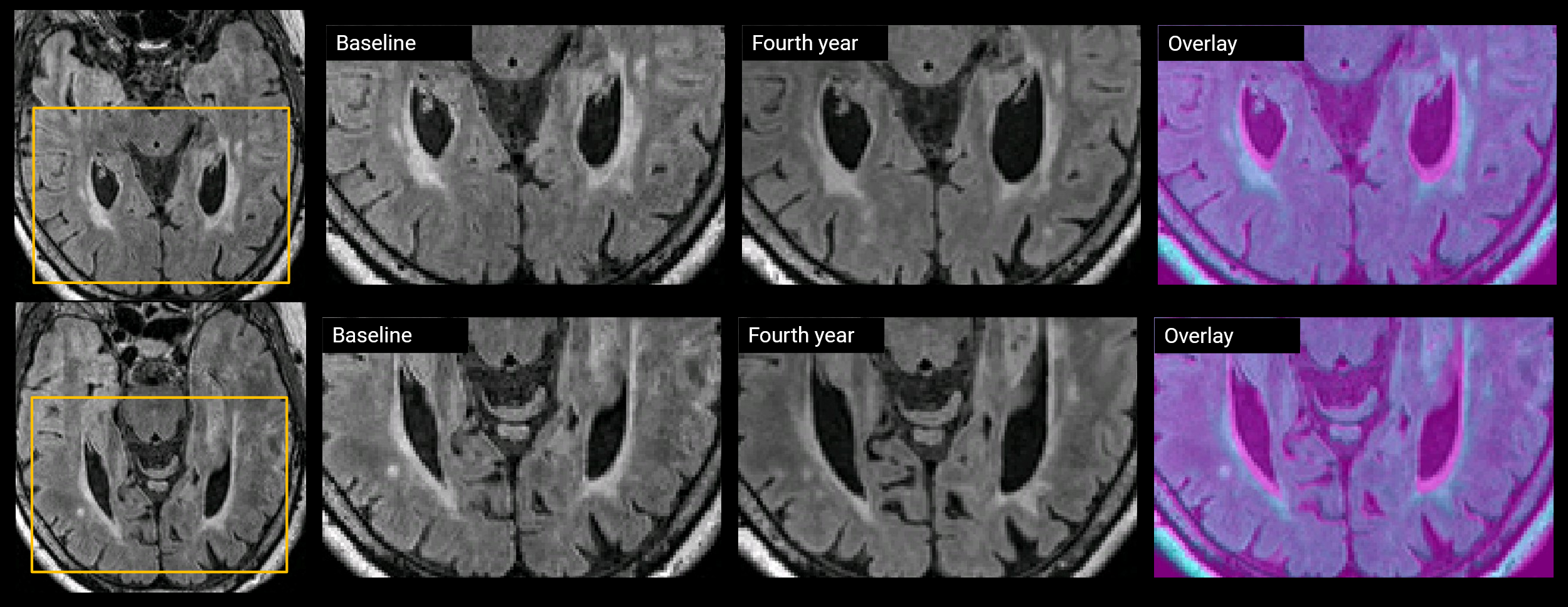

**Figure S2. Subject with regression of occipital WMH volumes over the course of four years.** The expansion of the lateral ventricles (highlighted in purple on the overlay image) causes tissue loss, which ultimately leads to a reduction in occipital WMH volumes.

Supplementary Models

### ULGCM – Global - Mean cortical thickness

```
lavaan 0.6.17 ended normally after 72 iterations

  Estimator                                         ML
  Optimization method                           NLMINB
  Number of model parameters                        30
  Number of equality constraints                     4

  Number of observations                           436
  Number of missing patterns                        21

Model Test User Model:
                                              Standard      Scaled
  Test Statistic                                31.800      21.650
  Degrees of freedom                                39          39
  P-value (Chi-square)                           0.787       0.989
  Scaling correction factor                                  1.469
    Yuan-Bentler correction (Mplus variant)

Model Test Baseline Model:

  Test statistic                              2515.603    1533.806
  Degrees of freedom                                45          45
  P-value                                        0.000       0.000
  Scaling correction factor                                  1.640

User Model versus Baseline Model:

  Comparative Fit Index (CFI)                    1.000       1.000
  Tucker-Lewis Index (TLI)                       1.003       1.013

  Robust Comparative Fit Index (CFI)                         1.000
  Robust Tucker-Lewis Index (TLI)                            1.014

Loglikelihood and Information Criteria:

  Loglikelihood user model (H0)              -3312.281   -3312.281
  Scaling correction factor                                  1.153
      for the MLR correction
  Loglikelihood unrestricted model (H1)      -3296.381   -3296.381
  Scaling correction factor                                  1.413
      for the MLR correction

  Akaike (AIC)                                6676.561    6676.561
  Bayesian (BIC)                              6782.580    6782.580
  Sample-size adjusted Bayesian (SABIC)       6700.070    6700.070

Root Mean Square Error of Approximation:

  RMSEA                                          0.000       0.000
  90 Percent confidence interval - lower         0.000       0.000
  90 Percent confidence interval - upper         0.023       0.000
  P-value H_0: RMSEA <= 0.050                    1.000       1.000
  P-value H_0: RMSEA >= 0.080                    0.000       0.000

  Robust RMSEA                                               0.000
  90 Percent confidence interval - lower                     0.000
  90 Percent confidence interval - upper                     0.000
  P-value H_0: Robust RMSEA <= 0.050                         1.000
  P-value H_0: Robust RMSEA >= 0.080                         0.000

Standardized Root Mean Square Residual:

  SRMR                                           0.017       0.017

Parameter Estimates:

  Standard errors                             Sandwich
  Information bread                           Observed
  Observed information based on                Hessian

Latent Variables:
                   Estimate  Std.Err  z-value  P(>|z|)   Std.lv  Std.all
  ix =~
    x1                1.000                               0.509    0.933
    x2                1.000                               0.509    0.925
    x3                1.000                               0.509    0.909
    x4                1.000                               0.509    0.886
    x5                1.000                               0.509    0.857
  sx =~
    x1                0.000                               0.000    0.000
    x2                1.000                               0.054    0.099
    x3                2.000                               0.109    0.194
    x4                3.000                               0.163    0.283
    x5                4.000                               0.217    0.365

Regressions:
                   Estimate  Std.Err  z-value  P(>|z|)   Std.lv  Std.all
  ix ~
    age_M00          -0.190    0.024   -8.077    0.000   -0.374   -0.374
    sex              -0.007    0.030   -0.247    0.805   -0.015   -0.015
    edyears          -0.020    0.025   -0.781    0.435   -0.039   -0.039
    vsclr_rsk_smcr   -0.038    0.025   -1.561    0.118   -0.075   -0.075
    eTIV             -0.104    0.030   -3.453    0.001   -0.203   -0.203
  sx ~
    age_M00          -0.008    0.005   -1.750    0.080   -0.147   -0.147
    sex               0.005    0.006    0.979    0.328    0.101    0.101
    edyears           0.004    0.004    0.896    0.370    0.071    0.071
    vsclr_rsk_smcr    0.000    0.005    0.026    0.980    0.002    0.002
    eTIV              0.007    0.005    1.404    0.160    0.133    0.133

Covariances:
                           Estimate  Std.Err  z-value  P(>|z|)   Std.lv  Std.all
 .ix ~~
   .sx                       -0.000    0.003   -0.091    0.928   -0.010   -0.010
  age_M00 ~~
    sex                      -0.168    0.047   -3.598    0.000   -0.168   -0.168
    edyears                  -0.128    0.050   -2.558    0.011   -0.128   -0.128
    vsclr_rsk_smcr            0.140    0.048    2.931    0.003    0.140    0.140
    eTIV                      0.072    0.047    1.539    0.124    0.072    0.072
  sex ~~
    edyears                  -0.233    0.044   -5.346    0.000   -0.233   -0.233
    vsclr_rsk_smcr           -0.195    0.045   -4.305    0.000   -0.195   -0.195
    eTIV                     -0.670    0.022  -30.135    0.000   -0.670   -0.670
  edyears ~~
    vsclr_rsk_smcr           -0.154    0.044   -3.502    0.000   -0.154   -0.154
    eTIV                      0.242    0.043    5.627    0.000    0.242    0.242
  vascular_risk_sumcorr ~~
    eTIV                      0.111    0.050    2.220    0.026    0.111    0.111

Intercepts:
                   Estimate  Std.Err  z-value  P(>|z|)   Std.lv  Std.all
   .ix                0.109    0.023    4.719    0.000    0.215    0.215
   .sx               -0.015    0.004   -3.270    0.001   -0.269   -0.269
   .x1                0.000                               0.000    0.000
   .x2                0.000                               0.000    0.000
   .x3                0.000                               0.000    0.000
   .x4                0.000                               0.000    0.000
   .x5                0.000                               0.000    0.000
    age_M00           0.000                               0.000    0.000
    edyears           0.000                               0.000    0.000
    sex               0.000                               0.000    0.000
    vsclr_rsk_smcr    0.000                               0.000    0.000
    eTIV              0.000                               0.000    0.000

Variances:
                   Estimate  Std.Err  z-value  P(>|z|)   Std.lv  Std.all
   .ix                0.207    0.017   12.325    0.000    0.797    0.797
   .sx                0.003    0.001    2.507    0.012    0.958    0.958
   .x1      (tht_)    0.039    0.004    9.301    0.000    0.039    0.129
   .x2      (tht_)    0.039    0.004    9.301    0.000    0.039    0.127
   .x3      (tht_)    0.039    0.004    9.301    0.000    0.039    0.123
   .x4      (tht_)    0.039    0.004    9.301    0.000    0.039    0.117
   .x5      (tht_)    0.039    0.004    9.301    0.000    0.039    0.109
    age_M00           1.000                               1.000    1.000
    sex               1.000                               1.000    1.000
    edyears           1.000                               1.000    1.000
    vsclr__           1.000                               1.000    1.000
    eTIV              1.000                               1.000    1.000

R-Square:
                   Estimate
    ix                0.203
    sx                0.042
    x1                0.871
    x2                0.873
    x3                0.877
    x4                0.883
    x5                0.891

                     lhs op                   rhs   label est.std    se       z pvalue ci.lower ci.upper
1                     ix =~                    x1           0.933 0.008 113.546  0.000    0.917    0.949
2                     ix =~                    x2           0.925 0.013  73.753  0.000    0.901    0.950
3                     ix =~                    x3           0.909 0.018  51.479  0.000    0.875    0.944
4                     ix =~                    x4           0.886 0.022  40.139  0.000    0.843    0.929
5                     ix =~                    x5           0.857 0.026  32.743  0.000    0.806    0.909
6                     sx =~                    x1           0.000 0.000      NA     NA    0.000    0.000
7                     sx =~                    x2           0.099 0.019   5.194  0.000    0.061    0.136
8                     sx =~                    x3           0.194 0.037   5.184  0.000    0.120    0.267
9                     sx =~                    x4           0.283 0.054   5.262  0.000    0.178    0.389
10                    sx =~                    x5           0.365 0.067   5.419  0.000    0.233    0.497
11                    ix ~~                    ix           0.797 0.036  22.027  0.000    0.726    0.868
12                    sx ~~                    sx           0.958 0.035  27.413  0.000    0.889    1.026
13                    ix ~~                    sx          -0.010 0.114  -0.092  0.927   -0.234    0.213
14                    ix ~1                                 0.215 0.047   4.619  0.000    0.124    0.306
15                    sx ~1                                -0.269 0.082  -3.263  0.001   -0.430   -0.107
16                    x1 ~~                    x1 theta_y   0.129 0.015   8.433  0.000    0.099    0.159
17                    x2 ~~                    x2 theta_y   0.127 0.015   8.617  0.000    0.098    0.156
18                    x3 ~~                    x3 theta_y   0.123 0.014   8.685  0.000    0.095    0.151
19                    x4 ~~                    x4 theta_y   0.117 0.013   8.664  0.000    0.090    0.143
20                    x5 ~~                    x5 theta_y   0.109 0.013   8.520  0.000    0.084    0.134
21                    x1 ~1                                 0.000 0.000      NA     NA    0.000    0.000
22                    x2 ~1                                 0.000 0.000      NA     NA    0.000    0.000
23                    x3 ~1                                 0.000 0.000      NA     NA    0.000    0.000
24                    x4 ~1                                 0.000 0.000      NA     NA    0.000    0.000
25                    x5 ~1                                 0.000 0.000      NA     NA    0.000    0.000
26                    ix  ~               age_M00          -0.374 0.042  -8.801  0.000   -0.457   -0.290
27                    ix  ~                   sex          -0.015 0.059  -0.247  0.805   -0.130    0.101
28                    ix  ~               edyears          -0.039 0.050  -0.783  0.434   -0.137    0.059
29                    ix  ~ vascular_risk_sumcorr          -0.075 0.048  -1.573  0.116   -0.169    0.019
30                    ix  ~                  eTIV          -0.203 0.058  -3.526  0.000   -0.316   -0.090
31                    sx  ~               age_M00          -0.147 0.089  -1.653  0.098   -0.322    0.027
32                    sx  ~                   sex           0.101 0.107   0.945  0.345   -0.109    0.311
33                    sx  ~               edyears           0.071 0.083   0.859  0.390   -0.091    0.233
34                    sx  ~ vascular_risk_sumcorr           0.002 0.084   0.026  0.980   -0.162    0.167
35                    sx  ~                  eTIV           0.133 0.094   1.406  0.160   -0.052    0.317
36               age_M00 ~1                                 0.000 0.000      NA     NA    0.000    0.000
37               edyears ~1                                 0.000 0.000      NA     NA    0.000    0.000
38                   sex ~1                                 0.000 0.000      NA     NA    0.000    0.000
39 vascular_risk_sumcorr ~1                                 0.000 0.000      NA     NA    0.000    0.000
40                  eTIV ~1                                 0.000 0.000      NA     NA    0.000    0.000
41               age_M00 ~~               age_M00           1.000 0.000      NA     NA    1.000    1.000
42                   sex ~~                   sex           1.000 0.000      NA     NA    1.000    1.000
43               edyears ~~               edyears           1.000 0.000      NA     NA    1.000    1.000
44 vascular_risk_sumcorr ~~ vascular_risk_sumcorr           1.000 0.000      NA     NA    1.000    1.000
45                  eTIV ~~                  eTIV           1.000 0.000      NA     NA    1.000    1.000
46               age_M00 ~~                   sex          -0.168 0.047  -3.598  0.000   -0.259   -0.076
47               age_M00 ~~               edyears          -0.128 0.050  -2.558  0.011   -0.226   -0.030
48               age_M00 ~~ vascular_risk_sumcorr           0.140 0.048   2.931  0.003    0.046    0.233
49               age_M00 ~~                  eTIV           0.072 0.047   1.539  0.124   -0.020    0.165
50                   sex ~~               edyears          -0.233 0.044  -5.346  0.000   -0.318   -0.147
51                   sex ~~ vascular_risk_sumcorr          -0.195 0.045  -4.305  0.000   -0.283   -0.106
52                   sex ~~                  eTIV          -0.670 0.022 -30.135  0.000   -0.713   -0.626
53               edyears ~~ vascular_risk_sumcorr          -0.154 0.044  -3.502  0.000   -0.240   -0.068
54               edyears ~~                  eTIV           0.242 0.043   5.627  0.000    0.158    0.326
55 vascular_risk_sumcorr ~~                  eTIV           0.111 0.050   2.220  0.026    0.013    0.209
```

### ULGCM – Global - Total WMH volumes

```
lavaan 0.6.17 ended normally after 76 iterations

  Estimator                                         ML
  Optimization method                           NLMINB
  Number of model parameters                        30
  Number of equality constraints                     4

  Number of observations                           436
  Number of missing patterns                        14

Model Test User Model:
                                              Standard      Scaled
  Test Statistic                                36.915      31.120
  Degrees of freedom                                39          39
  P-value (Chi-square)                           0.565       0.812
  Scaling correction factor                                  1.186
    Yuan-Bentler correction (Mplus variant)

Model Test Baseline Model:

  Test statistic                              4921.515    3706.964
  Degrees of freedom                                45          45
  P-value                                        0.000       0.000
  Scaling correction factor                                  1.328

User Model versus Baseline Model:

  Comparative Fit Index (CFI)                    1.000       1.000
  Tucker-Lewis Index (TLI)                       1.000       1.002

  Robust Comparative Fit Index (CFI)                         1.000
  Robust Tucker-Lewis Index (TLI)                            1.003

Loglikelihood and Information Criteria:

  Loglikelihood user model (H0)              -3121.296   -3121.296
  Scaling correction factor                                  1.029
      for the MLR correction
  Loglikelihood unrestricted model (H1)      -3102.839   -3102.839
  Scaling correction factor                                  1.187
      for the MLR correction

  Akaike (AIC)                                6294.593    6294.593
  Bayesian (BIC)                              6400.612    6400.612
  Sample-size adjusted Bayesian (SABIC)       6318.101    6318.101

Root Mean Square Error of Approximation:

  RMSEA                                          0.000       0.000
  90 Percent confidence interval - lower         0.000       0.000
  90 Percent confidence interval - upper         0.031       0.019
  P-value H_0: RMSEA <= 0.050                    1.000       1.000
  P-value H_0: RMSEA >= 0.080                    0.000       0.000

  Robust RMSEA                                               0.000
  90 Percent confidence interval - lower                     0.000
  90 Percent confidence interval - upper                     0.021
  P-value H_0: Robust RMSEA <= 0.050                         0.999
  P-value H_0: Robust RMSEA >= 0.080                         0.000

Standardized Root Mean Square Residual:

  SRMR                                           0.007       0.007

Parameter Estimates:

  Standard errors                             Sandwich
  Information bread                           Observed
  Observed information based on                Hessian

Latent Variables:
                   Estimate  Std.Err  z-value  P(>|z|)   Std.lv  Std.all
  iy =~
    y1                1.000                               0.962    0.992
    y2                1.000                               0.962    0.986
    y3                1.000                               0.962    0.976
    y4                1.000                               0.962    0.963
    y5                1.000                               0.962    0.946
  sy =~
    y1                0.000                               0.000    0.000
    y2                1.000                               0.064    0.066
    y3                2.000                               0.128    0.130
    y4                3.000                               0.192    0.192
    y5                4.000                               0.256    0.251

Regressions:
                   Estimate  Std.Err  z-value  P(>|z|)   Std.lv  Std.all
  iy ~
    age_M00           0.351    0.044    8.019    0.000    0.365    0.365
    sex               0.168    0.060    2.805    0.005    0.175    0.175
    edyears          -0.035    0.043   -0.814    0.416   -0.037   -0.037
    vsclr_rsk_smcr    0.086    0.044    1.954    0.051    0.090    0.090
    eTIV              0.247    0.056    4.390    0.000    0.256    0.256
  sy ~
    age_M00          -0.004    0.004   -0.860    0.390   -0.057   -0.057
    sex               0.003    0.006    0.459    0.646    0.040    0.040
    edyears          -0.004    0.004   -1.007    0.314   -0.066   -0.066
    vsclr_rsk_smcr    0.001    0.004    0.156    0.876    0.010    0.010
    eTIV              0.001    0.005    0.267    0.790    0.020    0.020

Covariances:
                           Estimate  Std.Err  z-value  P(>|z|)   Std.lv  Std.all
 .iy ~~
   .sy                        0.004    0.004    1.129    0.259    0.077    0.077
  age_M00 ~~
    sex                      -0.168    0.047   -3.598    0.000   -0.168   -0.168
    edyears                  -0.128    0.050   -2.558    0.011   -0.128   -0.128
    vsclr_rsk_smcr            0.140    0.048    2.931    0.003    0.140    0.140
    eTIV                      0.072    0.047    1.539    0.124    0.072    0.072
  sex ~~
    edyears                  -0.233    0.044   -5.346    0.000   -0.233   -0.233
    vsclr_rsk_smcr           -0.195    0.045   -4.305    0.000   -0.195   -0.195
    eTIV                     -0.670    0.022  -30.135    0.000   -0.670   -0.670
  edyears ~~
    vsclr_rsk_smcr           -0.154    0.044   -3.502    0.000   -0.154   -0.154
    eTIV                      0.242    0.043    5.627    0.000    0.242    0.242
  vascular_risk_sumcorr ~~
    eTIV                      0.111    0.050    2.220    0.026    0.111    0.111

Intercepts:
                   Estimate  Std.Err  z-value  P(>|z|)   Std.lv  Std.all
   .iy               -0.107    0.042   -2.548    0.011   -0.111   -0.111
   .sy                0.068    0.004   17.374    0.000    1.066    1.066
   .y1                0.000                               0.000    0.000
   .y2                0.000                               0.000    0.000
   .y3                0.000                               0.000    0.000
   .y4                0.000                               0.000    0.000
   .y5                0.000                               0.000    0.000
    age_M00           0.000                               0.000    0.000
    edyears           0.000                               0.000    0.000
    sex               0.000                               0.000    0.000
    vsclr_rsk_smcr    0.000                               0.000    0.000
    eTIV              0.000                               0.000    0.000

Variances:
                   Estimate  Std.Err  z-value  P(>|z|)   Std.lv  Std.all
   .iy                0.757    0.048   15.852    0.000    0.818    0.818
   .sy                0.004    0.001    5.482    0.000    0.991    0.991
   .y1      (tht_)    0.016    0.001   11.056    0.000    0.016    0.017
   .y2      (tht_)    0.016    0.001   11.056    0.000    0.016    0.017
   .y3      (tht_)    0.016    0.001   11.056    0.000    0.016    0.016
   .y4      (tht_)    0.016    0.001   11.056    0.000    0.016    0.016
   .y5      (tht_)    0.016    0.001   11.056    0.000    0.016    0.015
    age_M00           1.000                               1.000    1.000
    sex               1.000                               1.000    1.000
    edyears           1.000                               1.000    1.000
    vsclr__           1.000                               1.000    1.000
    eTIV              1.000                               1.000    1.000

R-Square:
                   Estimate
    iy                0.182
    sy                0.009
    y1                0.983
    y2                0.983
    y3                0.984
    y4                0.984
    y5                0.985
```

#### Standardised solution

```
                     lhs op                   rhs   label est.std    se        z pvalue ci.lower ci.upper
1                     iy =~                    y1           0.992 0.001 1135.796  0.000    0.990    0.993
2                     iy =~                    y2           0.986 0.005  210.736  0.000    0.977    0.995
3                     iy =~                    y3           0.976 0.009  111.058  0.000    0.959    0.993
4                     iy =~                    y4           0.963 0.013   76.508  0.000    0.938    0.987
5                     iy =~                    y5           0.946 0.016   59.027  0.000    0.915    0.978
6                     sy =~                    y1           0.000 0.000       NA     NA    0.000    0.000
7                     sy =~                    y2           0.066 0.006   10.750  0.000    0.054    0.077
8                     sy =~                    y3           0.130 0.012   10.752  0.000    0.106    0.153
9                     sy =~                    y4           0.192 0.018   10.821  0.000    0.157    0.227
10                    sy =~                    y5           0.251 0.023   10.953  0.000    0.206    0.296
11                    iy ~~                    iy           0.818 0.033   25.117  0.000    0.754    0.882
12                    sy ~~                    sy           0.991 0.012   80.306  0.000    0.967    1.015
```

```
13                    iy ~~                    sy           0.077 0.070    1.098  0.272   -0.061    0.215
14                    iy ~1                                -0.111 0.044   -2.516  0.012   -0.198   -0.025
15                    sy ~1                                 1.066 0.101   10.573  0.000    0.868    1.263
16                    y1 ~~                    y1 theta_y   0.017 0.002    9.675  0.000    0.013    0.020
17                    y2 ~~                    y2 theta_y   0.017 0.002    9.781  0.000    0.013    0.020
18                    y3 ~~                    y3 theta_y   0.016 0.002    9.818  0.000    0.013    0.019
19                    y4 ~~                    y4 theta_y   0.016 0.002    9.791  0.000    0.013    0.019
20                    y5 ~~                    y5 theta_y   0.015 0.002    9.708  0.000    0.012    0.018
21                    y1 ~1                                 0.000 0.000       NA     NA    0.000    0.000
22                    y2 ~1                                 0.000 0.000       NA     NA    0.000    0.000
23                    y3 ~1                                 0.000 0.000       NA     NA    0.000    0.000
24                    y4 ~1                                 0.000 0.000       NA     NA    0.000    0.000
25                    y5 ~1                                 0.000 0.000       NA     NA    0.000    0.000
26                    iy  ~               age_M00           0.365 0.043    8.462  0.000    0.281    0.450
27                    iy  ~                   sex           0.175 0.062    2.830  0.005    0.054    0.296
28                    iy  ~               edyears          -0.037 0.045   -0.815  0.415   -0.125    0.052
29                    iy  ~ vascular_risk_sumcorr           0.090 0.045    1.978  0.048    0.001    0.179
30                    iy  ~                  eTIV           0.256 0.057    4.535  0.000    0.146    0.367
31                    sy  ~               age_M00          -0.057 0.067   -0.862  0.389   -0.188    0.073
32                    sy  ~                   sex           0.040 0.087    0.455  0.649   -0.131    0.210
33                    sy  ~               edyears          -0.066 0.066   -0.998  0.318   -0.196    0.064
34                    sy  ~ vascular_risk_sumcorr           0.010 0.066    0.156  0.876   -0.120    0.141
35                    sy  ~                  eTIV           0.020 0.075    0.265  0.791   -0.127    0.166
36               age_M00 ~1                                 0.000 0.000       NA     NA    0.000    0.000
37               edyears ~1                                 0.000 0.000       NA     NA    0.000    0.000
38                   sex ~1                                 0.000 0.000       NA     NA    0.000    0.000
39 vascular_risk_sumcorr ~1                                 0.000 0.000       NA     NA    0.000    0.000
40                  eTIV ~1                                 0.000 0.000       NA     NA    0.000    0.000
41               age_M00 ~~               age_M00           1.000 0.000       NA     NA    1.000    1.000
42                   sex ~~                   sex           1.000 0.000       NA     NA    1.000    1.000
43               edyears ~~               edyears           1.000 0.000       NA     NA    1.000    1.000
44 vascular_risk_sumcorr ~~ vascular_risk_sumcorr           1.000 0.000       NA     NA    1.000    1.000
45                  eTIV ~~                  eTIV           1.000 0.000       NA     NA    1.000    1.000
46               age_M00 ~~                   sex          -0.168 0.047   -3.598  0.000   -0.259   -0.076
47               age_M00 ~~               edyears          -0.128 0.050   -2.558  0.011   -0.226   -0.030
48               age_M00 ~~ vascular_risk_sumcorr           0.140 0.048    2.931  0.003    0.046    0.233
49               age_M00 ~~                  eTIV           0.072 0.047    1.539  0.124   -0.020    0.165
50                   sex ~~               edyears          -0.233 0.044   -5.346  0.000   -0.318   -0.147
51                   sex ~~ vascular_risk_sumcorr          -0.195 0.045   -4.305  0.000   -0.283   -0.106
52                   sex ~~                  eTIV          -0.670 0.022  -30.135  0.000   -0.713   -0.626
53               edyears ~~ vascular_risk_sumcorr          -0.154 0.044   -3.502  0.000   -0.240   -0.068
54               edyears ~~                  eTIV           0.242 0.043    5.627  0.000    0.158    0.326
55 vascular_risk_sumcorr ~~                  eTIV           0.111 0.050    2.220  0.026    0.013    0.209
```

### ULGCM – Regional - Frontal WMH volumes

```
lavaan 0.6.17 ended normally after 63 iterations

  Estimator                                         ML
  Optimization method                           NLMINB
  Number of model parameters                        30
  Number of equality constraints                     4

  Number of observations                           416
  Number of missing patterns                        12

Model Test User Model:
                                              Standard      Scaled
  Test Statistic                                38.856      28.535
  Degrees of freedom                                39          39
  P-value (Chi-square)                           0.476       0.891
  Scaling correction factor                                  1.362
    Yuan-Bentler correction (Mplus variant)

Model Test Baseline Model:

  Test statistic                              3916.021    2581.130
  Degrees of freedom                                45          45
  P-value                                        0.000       0.000
  Scaling correction factor                                  1.517

User Model versus Baseline Model:

  Comparative Fit Index (CFI)                    1.000       1.000
  Tucker-Lewis Index (TLI)                       1.000       1.005

  Robust Comparative Fit Index (CFI)                         1.000
  Robust Tucker-Lewis Index (TLI)                            1.006

Loglikelihood and Information Criteria:

  Loglikelihood user model (H0)              -3404.377   -3404.377
  Scaling correction factor                                  1.068
      for the MLR correction
  Loglikelihood unrestricted model (H1)      -3384.950   -3384.950
  Scaling correction factor                                  1.310
      for the MLR correction

  Akaike (AIC)                                6860.755    6860.755
  Bayesian (BIC)                              6965.553    6965.553
  Sample-size adjusted Bayesian (SABIC)       6883.048    6883.048

Root Mean Square Error of Approximation:

  RMSEA                                          0.000       0.000
  90 Percent confidence interval - lower         0.000       0.000
  90 Percent confidence interval - upper         0.034       0.010
  P-value H_0: RMSEA <= 0.050                    0.999       1.000
  P-value H_0: RMSEA >= 0.080                    0.000       0.000

  Robust RMSEA                                               0.000
  90 Percent confidence interval - lower                     0.000
  90 Percent confidence interval - upper                     0.010
  P-value H_0: Robust RMSEA <= 0.050                         0.999
  P-value H_0: Robust RMSEA >= 0.080                         0.000

Standardized Root Mean Square Residual:

  SRMR                                           0.009       0.009

Parameter Estimates:

  Standard errors                             Sandwich
  Information bread                           Observed
  Observed information based on                Hessian

Latent Variables:
                   Estimate  Std.Err  z-value  P(>|z|)   Std.lv  Std.all
  iy =~
    y1                1.000                               0.980    0.982
    y2                1.000                               0.980    0.987
    y3                1.000                               0.980    0.988
    y4                1.000                               0.980    0.986
    y5                1.000                               0.980    0.980
  sy =~
    y1                0.000                               0.000    0.000
    y2                1.000                               0.060    0.060
    y3                2.000                               0.120    0.121
    y4                3.000                               0.180    0.181
    y5                4.000                               0.240    0.240

Regressions:
                   Estimate  Std.Err  z-value  P(>|z|)   Std.lv  Std.all
  iy ~
    age_M00           0.295    0.048    6.116    0.000    0.301    0.301
    sex               0.184    0.063    2.924    0.003    0.188    0.188
```

```
    edyears          -0.023    0.045   -0.503    0.615   -0.023   -0.023
    vsclr_rsk_smcr    0.075    0.048    1.579    0.114    0.077    0.077
    eTIV              0.253    0.060    4.194    0.000    0.258    0.258
  sy ~
    age_M00          -0.005    0.005   -0.972    0.331   -0.081   -0.081
    sex              -0.001    0.007   -0.101    0.920   -0.011   -0.011
    edyears          -0.005    0.005   -1.072    0.284   -0.087   -0.087
    vsclr_rsk_smcr    0.004    0.005    0.798    0.425    0.071    0.071
    eTIV             -0.003    0.006   -0.606    0.545   -0.058   -0.058

Covariances:
                           Estimate  Std.Err  z-value  P(>|z|)   Std.lv  Std.all
 .iy ~~
   .sy                       -0.006    0.005   -1.200    0.230   -0.102   -0.102
  age_M00 ~~
    sex                      -0.165    0.048   -3.439    0.001   -0.165   -0.165
    edyears                  -0.134    0.051   -2.609    0.009   -0.134   -0.134
    vsclr_rsk_smcr            0.135    0.048    2.791    0.005    0.135    0.135
    eTIV                      0.058    0.048    1.195    0.232    0.058    0.058
  sex ~~
    edyears                  -0.228    0.045   -5.111    0.000   -0.228   -0.228
    vsclr_rsk_smcr           -0.206    0.046   -4.491    0.000   -0.206   -0.206
    eTIV                     -0.672    0.023  -29.509    0.000   -0.672   -0.672
  edyears ~~
    vsclr_rsk_smcr           -0.148    0.045   -3.292    0.001   -0.148   -0.148
    eTIV                      0.248    0.044    5.690    0.000    0.248    0.248
  vascular_risk_sumcorr ~~
    eTIV                      0.119    0.051    2.330    0.020    0.119    0.119

Intercepts:
                   Estimate  Std.Err  z-value  P(>|z|)   Std.lv  Std.all
   .iy               -0.122    0.045   -2.677    0.007   -0.124   -0.124
   .sy                0.073    0.005   16.035    0.000    1.226    1.226
   .y1                0.000                               0.000    0.000
   .y2                0.000                               0.000    0.000
   .y3                0.000                               0.000    0.000
   .y4                0.000                               0.000    0.000
   .y5                0.000                               0.000    0.000
    age_M00           0.000                               0.000    0.000
    edyears           0.000                               0.000    0.000
    sex               0.000                               0.000    0.000
    vsclr_rsk_smcr    0.000                               0.000    0.000
    eTIV              0.000                               0.000    0.000

Variances:
                   Estimate  Std.Err  z-value  P(>|z|)   Std.lv  Std.all
   .iy                0.835    0.051   16.209    0.000    0.869    0.869
   .sy                0.004    0.001    3.977    0.000    0.978    0.978
   .y1      (tht_)    0.035    0.004    9.666    0.000    0.035    0.035
   .y2      (tht_)    0.035    0.004    9.666    0.000    0.035    0.036
   .y3      (tht_)    0.035    0.004    9.666    0.000    0.035    0.036
   .y4      (tht_)    0.035    0.004    9.666    0.000    0.035    0.036
   .y5      (tht_)    0.035    0.004    9.666    0.000    0.035    0.035
    age_M00           1.000                               1.000    1.000
    sex               1.000                               1.000    1.000
    edyears           1.000                               1.000    1.000
    vsclr__           1.000                               1.000    1.000
    eTIV              1.000                               1.000    1.000

R-Square:
                   Estimate
    iy                0.131
    sy                0.022
    y1                0.965
    y2                0.964
    y3                0.964
    y4                0.964
    y5                0.965
```

#### Standardised solution

```
                     lhs op                   rhs   label est.std    se       z pvalue ci.lower ci.upper
1                     iy =~                    y1           0.982 0.002 489.972  0.000    0.978    0.986
2                     iy =~                    y2           0.987 0.006 175.490  0.000    0.976    0.998
3                     iy =~                    y3           0.988 0.010  98.530  0.000    0.969    1.008
4                     iy =~                    y4           0.986 0.014  68.578  0.000    0.958    1.014
5                     iy =~                    y5           0.980 0.019  52.699  0.000    0.944    1.017
6                     sy =~                    y1           0.000 0.000      NA     NA    0.000    0.000
7                     sy =~                    y2           0.060 0.008   7.638  0.000    0.045    0.076
8                     sy =~                    y3           0.121 0.016   7.615  0.000    0.090    0.152
9                     sy =~                    y4           0.181 0.024   7.638  0.000    0.134    0.227
10                    sy =~                    y5           0.240 0.031   7.707  0.000    0.179    0.301
11                    iy ~~                    iy           0.869 0.031  28.249  0.000    0.809    0.930
12                    sy ~~                    sy           0.978 0.023  42.146  0.000    0.933    1.024
13                    iy ~~                    sy          -0.102 0.080  -1.275  0.202   -0.259    0.055
14                    iy ~1                                -0.124 0.046  -2.670  0.008   -0.215   -0.033
15                    sy ~1                                 1.226 0.166   7.407  0.000    0.902    1.551
16                    y1 ~~                    y1 theta_y   0.035 0.004   8.990  0.000    0.028    0.043
17                    y2 ~~                    y2 theta_y   0.036 0.004   9.203  0.000    0.028    0.043
18                    y3 ~~                    y3 theta_y   0.036 0.004   9.385  0.000    0.028    0.043
19                    y4 ~~                    y4 theta_y   0.036 0.004   9.525  0.000    0.028    0.043
20                    y5 ~~                    y5 theta_y   0.035 0.004   9.614  0.000    0.028    0.042
21                    y1 ~1                                 0.000 0.000      NA     NA    0.000    0.000
22                    y2 ~1                                 0.000 0.000      NA     NA    0.000    0.000
23                    y3 ~1                                 0.000 0.000      NA     NA    0.000    0.000
24                    y4 ~1                                 0.000 0.000      NA     NA    0.000    0.000
25                    y5 ~1                                 0.000 0.000      NA     NA    0.000    0.000
26                    iy  ~               age_M00           0.301 0.047   6.351  0.000    0.208    0.394
27                    iy  ~                   sex           0.188 0.064   2.955  0.003    0.063    0.313
```

```
28                    iy  ~               edyears          -0.023 0.046  -0.503  0.615   -0.114    0.067
29                    iy  ~ vascular_risk_sumcorr           0.077 0.048   1.592  0.111   -0.018    0.171
30                    iy  ~                  eTIV           0.258 0.060   4.327  0.000    0.141    0.375
31                    sy  ~               age_M00          -0.081 0.081  -0.992  0.321   -0.240    0.079
32                    sy  ~                   sex          -0.011 0.108  -0.101  0.920   -0.223    0.202
33                    sy  ~               edyears          -0.087 0.079  -1.103  0.270   -0.242    0.068
34                    sy  ~ vascular_risk_sumcorr           0.071 0.087   0.818  0.413   -0.100    0.242
35                    sy  ~                  eTIV          -0.058 0.096  -0.604  0.546   -0.247    0.131
36               age_M00 ~1                                 0.000 0.000      NA     NA    0.000    0.000
37               edyears ~1                                 0.000 0.000      NA     NA    0.000    0.000
38                   sex ~1                                 0.000 0.000      NA     NA    0.000    0.000
39 vascular_risk_sumcorr ~1                                 0.000 0.000      NA     NA    0.000    0.000
40                  eTIV ~1                                 0.000 0.000      NA     NA    0.000    0.000
41               age_M00 ~~               age_M00           1.000 0.000      NA     NA    1.000    1.000
42                   sex ~~                   sex           1.000 0.000      NA     NA    1.000    1.000
43               edyears ~~               edyears           1.000 0.000      NA     NA    1.000    1.000
44 vascular_risk_sumcorr ~~ vascular_risk_sumcorr           1.000 0.000      NA     NA    1.000    1.000
45                  eTIV ~~                  eTIV           1.000 0.000      NA     NA    1.000    1.000
46               age_M00 ~~                   sex          -0.165 0.048  -3.439  0.001   -0.258   -0.071
47               age_M00 ~~               edyears          -0.134 0.051  -2.609  0.009   -0.235   -0.033
48               age_M00 ~~ vascular_risk_sumcorr           0.135 0.048   2.791  0.005    0.040    0.230
49               age_M00 ~~                  eTIV           0.058 0.048   1.195  0.232   -0.037    0.152
50                   sex ~~               edyears          -0.228 0.045  -5.111  0.000   -0.316   -0.141
51                   sex ~~ vascular_risk_sumcorr          -0.206 0.046  -4.491  0.000   -0.295   -0.116
52                   sex ~~                  eTIV          -0.672 0.023 -29.509  0.000   -0.717   -0.627
53               edyears ~~ vascular_risk_sumcorr          -0.148 0.045  -3.292  0.001   -0.236   -0.060
54               edyears ~~                  eTIV           0.248 0.044   5.690  0.000    0.162    0.333
55 vascular_risk_sumcorr ~~                  eTIV           0.119 0.051   2.330  0.020    0.019    0.219
```

### ULGCM – Regional - Occipital WMH

```
lavaan 0.6.17 ended normally after 43 iterations

  Estimator                                         ML
  Optimization method                           NLMINB
  Number of model parameters                        30
  Number of equality constraints                     4

  Number of observations                           390
  Number of missing patterns                        19

Model Test User Model:
                                              Standard      Scaled
  Test Statistic                                51.992      42.664
  Degrees of freedom                                39          39
  P-value (Chi-square)                           0.080       0.317
  Scaling correction factor                                  1.219
    Yuan-Bentler correction (Mplus variant)

Model Test Baseline Model:

  Test statistic                              2531.059    1876.076
  Degrees of freedom                                45          45
  P-value                                        0.000       0.000
  Scaling correction factor                                  1.349

User Model versus Baseline Model:

  Comparative Fit Index (CFI)                    0.995       0.998
  Tucker-Lewis Index (TLI)                       0.994       0.998

  Robust Comparative Fit Index (CFI)                         0.995
  Robust Tucker-Lewis Index (TLI)                            0.994

Loglikelihood and Information Criteria:

  Loglikelihood user model (H0)              -3667.144   -3667.144
  Scaling correction factor                                  1.024
      for the MLR correction
  Loglikelihood unrestricted model (H1)      -3641.148   -3641.148
  Scaling correction factor                                  1.204
      for the MLR correction

  Akaike (AIC)                                7386.288    7386.288
  Bayesian (BIC)                              7489.408    7489.408
  Sample-size adjusted Bayesian (SABIC)       7406.912    7406.912

Root Mean Square Error of Approximation:

  RMSEA                                          0.029       0.016
  90 Percent confidence interval - lower         0.000       0.000
  90 Percent confidence interval - upper         0.049       0.038
  P-value H_0: RMSEA <= 0.050                    0.963       0.998
  P-value H_0: RMSEA >= 0.080                    0.000       0.000

  Robust RMSEA                                               0.034
  90 Percent confidence interval - lower                     0.000
  90 Percent confidence interval - upper                     0.062
  P-value H_0: Robust RMSEA <= 0.050                         0.799
  P-value H_0: Robust RMSEA >= 0.080                         0.002

Standardized Root Mean Square Residual:

  SRMR                                           0.014       0.014

Parameter Estimates:

  Standard errors                             Sandwich
  Information bread                           Observed
  Observed information based on                Hessian

Latent Variables:
                   Estimate  Std.Err  z-value  P(>|z|)   Std.lv  Std.all
  iy =~
    y1                1.000                               0.929    0.957
    y2                1.000                               0.929    0.964
    y3                1.000                               0.929    0.960
    y4                1.000                               0.929    0.944
    y5                1.000                               0.929    0.920
  sy =~
    y1                0.000                               0.000    0.000
    y2                1.000                               0.105    0.109
    y3                2.000                               0.210    0.217
    y4                3.000                               0.315    0.321
    y5                4.000                               0.421    0.416

Regressions:
                   Estimate  Std.Err  z-value  P(>|z|)   Std.lv  Std.all
  iy ~
    age_M00           0.297    0.043    6.984    0.000    0.320    0.320
    sex              -0.015    0.065   -0.234    0.815   -0.016   -0.016
    edyears          -0.047    0.049   -0.958    0.338   -0.051   -0.051
    vsclr_rsk_smcr    0.053    0.050    1.065    0.287    0.057    0.057
    eTIV              0.132    0.068    1.954    0.051    0.142    0.142
  sy ~
    age_M00          -0.000    0.008   -0.007    0.994   -0.001   -0.001
    sex               0.023    0.010    2.427    0.015    0.221    0.221
    edyears          -0.004    0.009   -0.447    0.655   -0.039   -0.039
    vsclr_rsk_smcr   -0.011    0.008   -1.451    0.147   -0.109   -0.109
    eTIV              0.037    0.010    3.683    0.000    0.354    0.354

Covariances:
                           Estimate  Std.Err  z-value  P(>|z|)   Std.lv  Std.all
 .iy ~~
   .sy                       -0.014    0.008   -1.806    0.071   -0.159   -0.159
  age_M00 ~~
    sex                      -0.186    0.049   -3.804    0.000   -0.186   -0.186
    edyears                  -0.097    0.053   -1.842    0.066   -0.097   -0.097
    vsclr_rsk_smcr            0.123    0.051    2.417    0.016    0.123    0.123
    eTIV                      0.097    0.049    1.966    0.049    0.097    0.097
  sex ~~
    edyears                  -0.219    0.047   -4.712    0.000   -0.219   -0.219
    vsclr_rsk_smcr           -0.186    0.048   -3.854    0.000   -0.186   -0.186
    eTIV                     -0.675    0.023  -28.872    0.000   -0.675   -0.675
  edyears ~~
    vsclr_rsk_smcr           -0.143    0.047   -3.054    0.002   -0.143   -0.143
    eTIV                      0.238    0.046    5.224    0.000    0.238    0.238
  vascular_risk_sumcorr ~~
    eTIV                      0.102    0.053    1.926    0.054    0.102    0.102

Intercepts:
                   Estimate  Std.Err  z-value  P(>|z|)   Std.lv  Std.all
   .iy               -0.102    0.045   -2.263    0.024   -0.110   -0.110
   .sy                0.058    0.008    7.478    0.000    0.551    0.551
   .y1                0.000                               0.000    0.000
   .y2                0.000                               0.000    0.000
   .y3                0.000                               0.000    0.000
   .y4                0.000                               0.000    0.000
   .y5                0.000                               0.000    0.000
    age_M00           0.000                               0.000    0.000
    edyears           0.000                               0.000    0.000
    sex               0.000                               0.000    0.000
    vsclr_rsk_smcr    0.000                               0.000    0.000
    eTIV              0.000                               0.000    0.000

Variances:
                   Estimate  Std.Err  z-value  P(>|z|)   Std.lv  Std.all
   .iy                0.734    0.053   13.936    0.000    0.851    0.851
   .sy                0.010    0.002    5.070    0.000    0.921    0.921
   .y1      (tht_)    0.080    0.009    9.165    0.000    0.080    0.085
   .y2      (tht_)    0.080    0.009    9.165    0.000    0.080    0.086
   .y3      (tht_)    0.080    0.009    9.165    0.000    0.080    0.085
   .y4      (tht_)    0.080    0.009    9.165    0.000    0.080    0.082
   .y5      (tht_)    0.080    0.009    9.165    0.000    0.080    0.078
    age_M00           1.000                               1.000    1.000
    sex               1.000                               1.000    1.000
    edyears           1.000                               1.000    1.000
    vsclr__           1.000                               1.000    1.000
    eTIV              1.000                               1.000    1.000

R-Square:
                   Estimate
    iy                0.149
    sy                0.079
    y1                0.915
    y2                0.914
    y3                0.915
    y4                0.918
    y5                0.922
```

#### Standardised solution

```
                     lhs op                   rhs   label est.std    se       z pvalue ci.lower ci.upper
1                     iy =~                    y1           0.957 0.005 173.990  0.000    0.946    0.968
2                     iy =~                    y2           0.964 0.011  91.482  0.000    0.943    0.985
3                     iy =~                    y3           0.960 0.017  57.394  0.000    0.927    0.992
4                     iy =~                    y4           0.944 0.022  42.525  0.000    0.901    0.988
5                     iy =~                    y5           0.920 0.027  34.300  0.000    0.867    0.972
6                     sy =~                    y1           0.000 0.000      NA     NA    0.000    0.000
7                     sy =~                    y2           0.109 0.011   9.499  0.000    0.087    0.132
8                     sy =~                    y3           0.217 0.023   9.408  0.000    0.172    0.262
9                     sy =~                    y4           0.321 0.034   9.497  0.000    0.254    0.387
10                    sy =~                    y5           0.416 0.043   9.754  0.000    0.333    0.500
11                    iy ~~                    iy           0.851 0.032  26.285  0.000    0.787    0.914
12                    sy ~~                    sy           0.921 0.040  23.230  0.000    0.843    0.999
13                    iy ~~                    sy          -0.159 0.078  -2.032  0.042   -0.313   -0.006
```

```
14                    iy ~1                                -0.110 0.049  -2.241  0.025   -0.205   -0.014
15                    sy ~1                                 0.551 0.086   6.425  0.000    0.383    0.720
16                    y1 ~~                    y1 theta_y   0.085 0.011   8.033  0.000    0.064    0.105
17                    y2 ~~                    y2 theta_y   0.086 0.010   8.242  0.000    0.065    0.106
18                    y3 ~~                    y3 theta_y   0.085 0.010   8.308  0.000    0.065    0.105
19                    y4 ~~                    y4 theta_y   0.082 0.010   8.238  0.000    0.063    0.102
20                    y5 ~~                    y5 theta_y   0.078 0.010   8.053  0.000    0.059    0.097
21                    y1 ~1                                 0.000 0.000      NA     NA    0.000    0.000
22                    y2 ~1                                 0.000 0.000      NA     NA    0.000    0.000
23                    y3 ~1                                 0.000 0.000      NA     NA    0.000    0.000
24                    y4 ~1                                 0.000 0.000      NA     NA    0.000    0.000
25                    y5 ~1                                 0.000 0.000      NA     NA    0.000    0.000
26                    iy  ~               age_M00           0.320 0.043   7.475  0.000    0.236    0.404
27                    iy  ~                   sex          -0.016 0.070  -0.233  0.816   -0.153    0.121
28                    iy  ~               edyears          -0.051 0.053  -0.959  0.338   -0.154    0.053
29                    iy  ~ vascular_risk_sumcorr           0.057 0.054   1.071  0.284   -0.048    0.162
30                    iy  ~                  eTIV           0.142 0.071   1.988  0.047    0.002    0.282
31                    sy  ~               age_M00          -0.001 0.075  -0.007  0.994   -0.147    0.146
32                    sy  ~                   sex           0.221 0.088   2.510  0.012    0.048    0.393
```

```
33                    sy  ~               edyears          -0.039 0.086  -0.451  0.652   -0.208    0.130
34                    sy  ~ vascular_risk_sumcorr          -0.109 0.072  -1.502  0.133   -0.250    0.033
35                    sy  ~                  eTIV           0.354 0.088   4.034  0.000    0.182    0.525
36               age_M00 ~1                                 0.000 0.000      NA     NA    0.000    0.000
37               edyears ~1                                 0.000 0.000      NA     NA    0.000    0.000
38                   sex ~1                                 0.000 0.000      NA     NA    0.000    0.000
39 vascular_risk_sumcorr ~1                                 0.000 0.000      NA     NA    0.000    0.000
40                  eTIV ~1                                 0.000 0.000      NA     NA    0.000    0.000
41               age_M00 ~~               age_M00           1.000 0.000      NA     NA    1.000    1.000
42                   sex ~~                   sex           1.000 0.000      NA     NA    1.000    1.000
43               edyears ~~               edyears           1.000 0.000      NA     NA    1.000    1.000
44 vascular_risk_sumcorr ~~ vascular_risk_sumcorr           1.000 0.000      NA     NA    1.000    1.000
45                  eTIV ~~                  eTIV           1.000 0.000      NA     NA    1.000    1.000
46               age_M00 ~~                   sex          -0.186 0.049  -3.804  0.000   -0.282   -0.090
47               age_M00 ~~               edyears          -0.097 0.053  -1.842  0.066   -0.200    0.006
48               age_M00 ~~ vascular_risk_sumcorr           0.123 0.051   2.417  0.016    0.023    0.223
49               age_M00 ~~                  eTIV           0.097 0.049   1.966  0.049    0.000    0.194
50                   sex ~~               edyears          -0.219 0.047  -4.712  0.000   -0.310   -0.128
51                   sex ~~ vascular_risk_sumcorr          -0.186 0.048  -3.854  0.000   -0.281   -0.092
52                   sex ~~                  eTIV          -0.675 0.023 -28.872  0.000   -0.721   -0.629
53               edyears ~~ vascular_risk_sumcorr          -0.143 0.047  -3.054  0.002   -0.235   -0.051
54               edyears ~~                  eTIV           0.238 0.046   5.224  0.000    0.149    0.328
55 vascular_risk_sumcorr ~~                  eTIV           0.102 0.053   1.926  0.054   -0.002    0.206
```

### ULGCM – Regional - Parietal WMH

```
lavaan 0.6.17 ended normally after 52 iterations

  Estimator                                         ML
  Optimization method                           NLMINB
  Number of model parameters                        30
  Number of equality constraints                     4

  Number of observations                           387
  Number of missing patterns                        15

Model Test User Model:
                                              Standard      Scaled
  Test Statistic                                55.682      41.442
  Degrees of freedom                                39          39
  P-value (Chi-square)                           0.041       0.365
  Scaling correction factor                                  1.344
    Yuan-Bentler correction (Mplus variant)

Model Test Baseline Model:

  Test statistic                              3717.792    2447.928
  Degrees of freedom                                45          45
  P-value                                        0.000       0.000
  Scaling correction factor                                  1.519

User Model versus Baseline Model:

  Comparative Fit Index (CFI)                    0.995       0.999
  Tucker-Lewis Index (TLI)                       0.995       0.999

  Robust Comparative Fit Index (CFI)                         1.000
  Robust Tucker-Lewis Index (TLI)                            1.001

Loglikelihood and Information Criteria:

  Loglikelihood user model (H0)              -3121.092   -3121.092
  Scaling correction factor                                  1.099
      for the MLR correction
  Loglikelihood unrestricted model (H1)      -3093.251   -3093.251
  Scaling correction factor                                  1.313
      for the MLR correction

  Akaike (AIC)                                6294.183    6294.183
  Bayesian (BIC)                              6397.102    6397.102
  Sample-size adjusted Bayesian (SABIC)       6314.607    6314.607

Root Mean Square Error of Approximation:

  RMSEA                                          0.033       0.013
  90 Percent confidence interval - lower         0.007       0.000
  90 Percent confidence interval - upper         0.052       0.035
  P-value H_0: RMSEA <= 0.050                    0.928       0.999
  P-value H_0: RMSEA >= 0.080                    0.000       0.000

  Robust RMSEA                                               0.000
  90 Percent confidence interval - lower                     0.000
  90 Percent confidence interval - upper                     0.049
  P-value H_0: Robust RMSEA <= 0.050                         0.953
  P-value H_0: Robust RMSEA >= 0.080                         0.000

Standardized Root Mean Square Residual:

  SRMR                                           0.009       0.009

Parameter Estimates:

  Standard errors                             Sandwich
  Information bread                           Observed
  Observed information based on                Hessian

Latent Variables:
                   Estimate  Std.Err  z-value  P(>|z|)   Std.lv  Std.all
  iy =~
    y1                1.000                               1.023    0.986
    y2                1.000                               1.023    1.010
    y3                1.000                               1.023    1.029
    y4                1.000                               1.023    1.042
    y5                1.000                               1.023    1.050
  sy =~
    y1                0.000                               0.000    0.000
    y2                1.000                               0.077    0.076
    y3                2.000                               0.154    0.155
    y4                3.000                               0.231    0.235
    y5                4.000                               0.307    0.316

Regressions:
                   Estimate  Std.Err  z-value  P(>|z|)   Std.lv  Std.all
  iy ~
    age_M00           0.300    0.050    5.987    0.000    0.293    0.293
    sex               0.073    0.069    1.060    0.289    0.072    0.072
    edyears          -0.020    0.051   -0.396    0.692   -0.020   -0.020
    vsclr_rsk_smcr    0.077    0.054    1.444    0.149    0.076    0.076
    eTIV              0.187    0.068    2.762    0.006    0.183    0.183
  sy ~
    age_M00          -0.018    0.006   -3.139    0.002   -0.233   -0.233
    sex               0.003    0.007    0.453    0.651    0.038    0.038
    edyears          -0.013    0.005   -2.416    0.016   -0.166   -0.166
    vsclr_rsk_smcr   -0.001    0.005   -0.224    0.822   -0.016   -0.016
    eTIV             -0.004    0.007   -0.584    0.559   -0.050   -0.050

Covariances:
                           Estimate  Std.Err  z-value  P(>|z|)   Std.lv  Std.all
 .iy ~~
   .sy                       -0.021    0.006   -3.678    0.000   -0.305   -0.305
  age_M00 ~~
    sex                      -0.162    0.050   -3.269    0.001   -0.162   -0.162
    edyears                  -0.129    0.053   -2.433    0.015   -0.129   -0.129
    vsclr_rsk_smcr            0.108    0.050    2.150    0.032    0.108    0.108
    eTIV                      0.066    0.050    1.327    0.185    0.066    0.066
  sex ~~
    edyears                  -0.243    0.046   -5.333    0.000   -0.243   -0.243
    vsclr_rsk_smcr           -0.204    0.048   -4.280    0.000   -0.204   -0.204
    eTIV                     -0.661    0.024  -27.656    0.000   -0.661   -0.661
  edyears ~~
    vsclr_rsk_smcr           -0.141    0.047   -3.010    0.003   -0.141   -0.141
    eTIV                      0.254    0.046    5.575    0.000    0.254    0.254
  vascular_risk_sumcorr ~~
    eTIV                      0.122    0.053    2.273    0.023    0.122    0.122

Intercepts:
                   Estimate  Std.Err  z-value  P(>|z|)   Std.lv  Std.all
   .iy               -0.153    0.049   -3.106    0.002   -0.150   -0.150
   .sy                0.084    0.005   16.396    0.000    1.099    1.099
   .y1                0.000                               0.000    0.000
   .y2                0.000                               0.000    0.000
   .y3                0.000                               0.000    0.000
   .y4                0.000                               0.000    0.000
   .y5                0.000                               0.000    0.000
    age_M00           0.000                               0.000    0.000
    edyears           0.000                               0.000    0.000
    sex               0.000                               0.000    0.000
    vsclr_rsk_smcr    0.000                               0.000    0.000
    eTIV              0.000                               0.000    0.000

Variances:
                   Estimate  Std.Err  z-value  P(>|z|)   Std.lv  Std.all
   .iy                0.920    0.058   15.743    0.000    0.880    0.880
   .sy                0.005    0.001    4.254    0.000    0.909    0.909
   .y1      (tht_)    0.029    0.004    8.119    0.000    0.029    0.027
   .y2      (tht_)    0.029    0.004    8.119    0.000    0.029    0.028
   .y3      (tht_)    0.029    0.004    8.119    0.000    0.029    0.029
   .y4      (tht_)    0.029    0.004    8.119    0.000    0.029    0.030
   .y5      (tht_)    0.029    0.004    8.119    0.000    0.029    0.031
    age_M00           1.000                               1.000    1.000
    sex               1.000                               1.000    1.000
    edyears           1.000                               1.000    1.000
    vsclr__           1.000                               1.000    1.000
    eTIV              1.000                               1.000    1.000

R-Square:
                   Estimate
    iy                0.120
    sy                0.091
    y1                0.973
    y2                0.972
    y3                0.971
    y4                0.970
    y5                0.969
```

#### Standardised solution

```
                     lhs op                   rhs   label est.std    se       z pvalue ci.lower ci.upper
1                     iy =~                    y1           0.986 0.002 584.587  0.000    0.983    0.990
2                     iy =~                    y2           1.010 0.006 158.816  0.000    0.997    1.022
3                     iy =~                    y3           1.029 0.012  84.600  0.000    1.005    1.053
4                     iy =~                    y4           1.042 0.018  58.407  0.000    1.007    1.077
5                     iy =~                    y5           1.050 0.023  45.105  0.000    1.004    1.096
6                     sy =~                    y1           0.000 0.000      NA     NA    0.000    0.000
7                     sy =~                    y2           0.076 0.009   8.364  0.000    0.058    0.094
8                     sy =~                    y3           0.155 0.019   8.197  0.000    0.118    0.192
9                     sy =~                    y4           0.235 0.029   8.108  0.000    0.178    0.292
10                    sy =~                    y5           0.316 0.039   8.101  0.000    0.239    0.392
11                    iy ~~                    iy           0.880 0.032  27.580  0.000    0.817    0.942
12                    sy ~~                    sy           0.909 0.040  22.702  0.000    0.831    0.988
13                    iy ~~                    sy          -0.305 0.066  -4.654  0.000   -0.434   -0.177
```

```
14                    iy ~1                                -0.150 0.049  -3.081  0.002   -0.245   -0.054
15                    sy ~1                                 1.099 0.115   9.572  0.000    0.874    1.324
16                    y1 ~~                    y1 theta_y   0.027 0.003   8.138  0.000    0.021    0.034
17                    y2 ~~                    y2 theta_y   0.028 0.003   8.165  0.000    0.022    0.035
18                    y3 ~~                    y3 theta_y   0.029 0.004   8.092  0.000    0.022    0.037
19                    y4 ~~                    y4 theta_y   0.030 0.004   7.932  0.000    0.023    0.038
20                    y5 ~~                    y5 theta_y   0.031 0.004   7.705  0.000    0.023    0.039
21                    y1 ~1                                 0.000 0.000      NA     NA    0.000    0.000
22                    y2 ~1                                 0.000 0.000      NA     NA    0.000    0.000
23                    y3 ~1                                 0.000 0.000      NA     NA    0.000    0.000
24                    y4 ~1                                 0.000 0.000      NA     NA    0.000    0.000
25                    y5 ~1                                 0.000 0.000      NA     NA    0.000    0.000
26                    iy  ~               age_M00           0.293 0.047   6.213  0.000    0.201    0.386
27                    iy  ~                   sex           0.072 0.068   1.060  0.289   -0.061    0.204
28                    iy  ~               edyears          -0.020 0.050  -0.397  0.692   -0.117    0.078
29                    iy  ~ vascular_risk_sumcorr           0.076 0.052   1.452  0.147   -0.026    0.178
30                    iy  ~                  eTIV           0.183 0.065   2.812  0.005    0.056    0.311
31                    sy  ~               age_M00          -0.233 0.068  -3.431  0.001   -0.366   -0.100
```

```
32                    sy  ~                   sex           0.038 0.085   0.451  0.652   -0.128    0.205
33                    sy  ~               edyears          -0.166 0.066  -2.506  0.012   -0.296   -0.036
```

```
34                    sy  ~ vascular_risk_sumcorr          -0.016 0.070  -0.225  0.822   -0.154    0.122
35                    sy  ~                  eTIV          -0.050 0.084  -0.594  0.553   -0.214    0.115
36               age_M00 ~1                                 0.000 0.000      NA     NA    0.000    0.000
37               edyears ~1                                 0.000 0.000      NA     NA    0.000    0.000
38                   sex ~1                                 0.000 0.000      NA     NA    0.000    0.000
39 vascular_risk_sumcorr ~1                                 0.000 0.000      NA     NA    0.000    0.000
40                  eTIV ~1                                 0.000 0.000      NA     NA    0.000    0.000
41               age_M00 ~~               age_M00           1.000 0.000      NA     NA    1.000    1.000
42                   sex ~~                   sex           1.000 0.000      NA     NA    1.000    1.000
43               edyears ~~               edyears           1.000 0.000      NA     NA    1.000    1.000
44 vascular_risk_sumcorr ~~ vascular_risk_sumcorr           1.000 0.000      NA     NA    1.000    1.000
45                  eTIV ~~                  eTIV           1.000 0.000      NA     NA    1.000    1.000
46               age_M00 ~~                   sex          -0.162 0.050  -3.269  0.001   -0.260   -0.065
47               age_M00 ~~               edyears          -0.129 0.053  -2.433  0.015   -0.234   -0.025
48               age_M00 ~~ vascular_risk_sumcorr           0.108 0.050   2.150  0.032    0.010    0.206
49               age_M00 ~~                  eTIV           0.066 0.050   1.327  0.185   -0.032    0.164
50                   sex ~~               edyears          -0.243 0.046  -5.333  0.000   -0.332   -0.154
51                   sex ~~ vascular_risk_sumcorr          -0.204 0.048  -4.280  0.000   -0.297   -0.111
52                   sex ~~                  eTIV          -0.661 0.024 -27.656  0.000   -0.708   -0.614
53               edyears ~~ vascular_risk_sumcorr          -0.141 0.047  -3.010  0.003   -0.233   -0.049
54               edyears ~~                  eTIV           0.254 0.046   5.575  0.000    0.165    0.343
55 vascular_risk_sumcorr ~~                  eTIV           0.122 0.053   2.273  0.023    0.017    0.226
```

### ULGCM – Regional - Temporal WMH

```
lavaan 0.6.17 ended normally after 49 iterations

  Estimator                                         ML
  Optimization method                           NLMINB
  Number of model parameters                        30
  Number of equality constraints                     4

  Number of observations                           318
  Number of missing patterns                        17

Model Test User Model:
                                              Standard      Scaled
  Test Statistic                                50.094      39.182
  Degrees of freedom                                39          39
  P-value (Chi-square)                           0.110       0.462
  Scaling correction factor                                  1.278
    Yuan-Bentler correction (Mplus variant)

Model Test Baseline Model:

  Test statistic                              1947.549    1376.019
  Degrees of freedom                                45          45
  P-value                                        0.000       0.000
  Scaling correction factor                                  1.415

User Model versus Baseline Model:

  Comparative Fit Index (CFI)                    0.994       1.000
  Tucker-Lewis Index (TLI)                       0.993       1.000

  Robust Comparative Fit Index (CFI)                         0.985
  Robust Tucker-Lewis Index (TLI)                            0.983

Loglikelihood and Information Criteria:

  Loglikelihood user model (H0)              -3065.596   -3065.596
  Scaling correction factor                                  1.058
      for the MLR correction
  Loglikelihood unrestricted model (H1)      -3040.549   -3040.549
  Scaling correction factor                                  1.255
      for the MLR correction

  Akaike (AIC)                                6183.192    6183.192
  Bayesian (BIC)                              6281.005    6281.005
  Sample-size adjusted Bayesian (SABIC)       6198.539    6198.539

Root Mean Square Error of Approximation:

  RMSEA                                          0.030       0.004
  90 Percent confidence interval - lower         0.000       0.000
  90 Percent confidence interval - upper         0.052       0.036
  P-value H_0: RMSEA <= 0.050                    0.931       0.998
  P-value H_0: RMSEA >= 0.080                    0.000       0.000

  Robust RMSEA                                               0.056
  90 Percent confidence interval - lower                     0.017
  90 Percent confidence interval - upper                     0.086
  P-value H_0: Robust RMSEA <= 0.050                         0.359
  P-value H_0: Robust RMSEA >= 0.080                         0.096

Standardized Root Mean Square Residual:

  SRMR                                           0.017       0.017

Parameter Estimates:

  Standard errors                             Sandwich
  Information bread                           Observed
  Observed information based on                Hessian

Latent Variables:
                   Estimate  Std.Err  z-value  P(>|z|)   Std.lv  Std.all
  iy =~
    y1                1.000                               0.969    0.950
    y2                1.000                               0.969    0.966
    y3                1.000                               0.969    0.978
    y4                1.000                               0.969    0.984
    y5                1.000                               0.969    0.984
  sy =~
    y1                0.000                               0.000    0.000
    y2                1.000                               0.075    0.075
    y3                2.000                               0.150    0.151
    y4                3.000                               0.224    0.228
    y5                4.000                               0.299    0.304

Regressions:
                   Estimate  Std.Err  z-value  P(>|z|)   Std.lv  Std.all
  iy ~
    age_M00           0.189    0.054    3.497    0.000    0.195    0.195
    sex               0.126    0.077    1.644    0.100    0.130    0.130
    edyears          -0.096    0.056   -1.707    0.088   -0.099   -0.099
    vsclr_rsk_smcr   -0.016    0.059   -0.268    0.789   -0.016   -0.016
    eTIV              0.229    0.074    3.079    0.002    0.237    0.237
  sy ~
    age_M00          -0.015    0.007   -2.039    0.041   -0.198   -0.198
    sex              -0.016    0.010   -1.620    0.105   -0.215   -0.215
    edyears          -0.002    0.008   -0.192    0.848   -0.021   -0.021
    vsclr_rsk_smcr    0.007    0.011    0.607    0.544    0.089    0.089
    eTIV             -0.009    0.009   -1.055    0.292   -0.127   -0.127

Covariances:
                           Estimate  Std.Err  z-value  P(>|z|)   Std.lv  Std.all
 .iy ~~
   .sy                       -0.017    0.011   -1.470    0.142   -0.247   -0.247
  age_M00 ~~
    sex                      -0.187    0.054   -3.446    0.001   -0.187   -0.187
    edyears                  -0.128    0.058   -2.188    0.029   -0.128   -0.128
    vsclr_rsk_smcr            0.093    0.057    1.643    0.100    0.093    0.093
    eTIV                      0.054    0.054    0.994    0.320    0.054    0.054
  sex ~~
    edyears                  -0.244    0.050   -4.850    0.000   -0.244   -0.244
    vsclr_rsk_smcr           -0.198    0.052   -3.778    0.000   -0.198   -0.198
    eTIV                     -0.665    0.027  -24.937    0.000   -0.665   -0.665
  edyears ~~
    vsclr_rsk_smcr           -0.133    0.052   -2.543    0.011   -0.133   -0.133
    eTIV                      0.235    0.049    4.768    0.000    0.235    0.235
  vascular_risk_sumcorr ~~
    eTIV                      0.132    0.061    2.154    0.031    0.132    0.132

Intercepts:
                   Estimate  Std.Err  z-value  P(>|z|)   Std.lv  Std.all
   .iy               -0.166    0.054   -3.049    0.002   -0.172   -0.172
   .sy                0.072    0.008    8.472    0.000    0.960    0.960
   .y1                0.000                               0.000    0.000
   .y2                0.000                               0.000    0.000
   .y3                0.000                               0.000    0.000
   .y4                0.000                               0.000    0.000
   .y5                0.000                               0.000    0.000
    age_M00           0.000                               0.000    0.000
    edyears           0.000                               0.000    0.000
    sex               0.000                               0.000    0.000
    vsclr_rsk_smcr    0.000                               0.000    0.000
    eTIV              0.000                               0.000    0.000

Variances:
                   Estimate  Std.Err  z-value  P(>|z|)   Std.lv  Std.all
   .iy                0.868    0.071   12.258    0.000    0.925    0.925
   .sy                0.005    0.003    1.530    0.126    0.940    0.940
   .y1      (tht_)    0.101    0.011    9.158    0.000    0.101    0.097
   .y2      (tht_)    0.101    0.011    9.158    0.000    0.101    0.100
   .y3      (tht_)    0.101    0.011    9.158    0.000    0.101    0.102
   .y4      (tht_)    0.101    0.011    9.158    0.000    0.101    0.104
   .y5      (tht_)    0.101    0.011    9.158    0.000    0.101    0.104
    age_M00           1.000                               1.000    1.000
    sex               1.000                               1.000    1.000
    edyears           1.000                               1.000    1.000
    vsclr__           1.000                               1.000    1.000
    eTIV              1.000                               1.000    1.000

R-Square:
                   Estimate
    iy                0.075
    sy                0.060
    y1                0.903
    y2                0.900
    y3                0.898
    y4                0.896
    y5                0.896
```

#### Standardised solution

```
                     lhs op                   rhs   label est.std    se       z pvalue ci.lower ci.upper
1                     iy =~                    y1           0.950 0.006 148.267  0.000    0.938    0.963
2                     iy =~                    y2           0.966 0.014  69.878  0.000    0.939    0.993
3                     iy =~                    y3           0.978 0.022  44.672  0.000    0.935    1.021
4                     iy =~                    y4           0.984 0.029  33.460  0.000    0.926    1.041
5                     iy =~                    y5           0.984 0.037  26.803  0.000    0.912    1.056
6                     sy =~                    y1           0.000 0.000      NA     NA    0.000    0.000
7                     sy =~                    y2           0.075 0.023   3.263  0.001    0.030    0.119
8                     sy =~                    y3           0.151 0.047   3.237  0.001    0.060    0.242
9                     sy =~                    y4           0.228 0.070   3.244  0.001    0.090    0.365
10                    sy =~                    y5           0.304 0.092   3.287  0.001    0.123    0.485
11                    iy ~~                    iy           0.925 0.031  29.931  0.000    0.865    0.986
12                    sy ~~                    sy           0.940 0.066  14.175  0.000    0.810    1.070
13                    iy ~~                    sy          -0.247 0.125  -1.982  0.048   -0.492   -0.003
```

```
14                    iy ~1                                -0.172 0.057  -3.002  0.003   -0.284   -0.060
15                    sy ~1                                 0.960 0.289   3.320  0.001    0.393    1.527
16                    y1 ~~                    y1 theta_y   0.097 0.012   7.948  0.000    0.073    0.121
17                    y2 ~~                    y2 theta_y   0.100 0.012   8.460  0.000    0.077    0.123
18                    y3 ~~                    y3 theta_y   0.102 0.012   8.657  0.000    0.079    0.126
19                    y4 ~~                    y4 theta_y   0.104 0.012   8.484  0.000    0.080    0.128
20                    y5 ~~                    y5 theta_y   0.104 0.013   7.973  0.000    0.078    0.129
21                    y1 ~1                                 0.000 0.000      NA     NA    0.000    0.000
22                    y2 ~1                                 0.000 0.000      NA     NA    0.000    0.000
23                    y3 ~1                                 0.000 0.000      NA     NA    0.000    0.000
24                    y4 ~1                                 0.000 0.000      NA     NA    0.000    0.000
25                    y5 ~1                                 0.000 0.000      NA     NA    0.000    0.000
26                    iy  ~               age_M00           0.195 0.055   3.563  0.000    0.088    0.302
27                    iy  ~                   sex           0.130 0.078   1.666  0.096   -0.023    0.283
28                    iy  ~               edyears          -0.099 0.058  -1.715  0.086   -0.213    0.014
```

```
29                    iy  ~ vascular_risk_sumcorr          -0.016 0.061  -0.268  0.789   -0.137    0.104
30                    iy  ~                  eTIV           0.237 0.075   3.173  0.002    0.090    0.383
31                    sy  ~               age_M00          -0.198 0.121  -1.637  0.102   -0.435    0.039
32                    sy  ~                   sex          -0.215 0.144  -1.493  0.135   -0.498    0.067
33                    sy  ~               edyears          -0.021 0.112  -0.191  0.849   -0.241    0.199
34                    sy  ~ vascular_risk_sumcorr           0.089 0.148   0.600  0.548   -0.202    0.380
35                    sy  ~                  eTIV          -0.127 0.127  -0.996  0.319   -0.377    0.123
36               age_M00 ~1                                 0.000 0.000      NA     NA    0.000    0.000
37               edyears ~1                                 0.000 0.000      NA     NA    0.000    0.000
38                   sex ~1                                 0.000 0.000      NA     NA    0.000    0.000
39 vascular_risk_sumcorr ~1                                 0.000 0.000      NA     NA    0.000    0.000
40                  eTIV ~1                                 0.000 0.000      NA     NA    0.000    0.000
41               age_M00 ~~               age_M00           1.000 0.000      NA     NA    1.000    1.000
42                   sex ~~                   sex           1.000 0.000      NA     NA    1.000    1.000
43               edyears ~~               edyears           1.000 0.000      NA     NA    1.000    1.000
44 vascular_risk_sumcorr ~~ vascular_risk_sumcorr           1.000 0.000      NA     NA    1.000    1.000
45                  eTIV ~~                  eTIV           1.000 0.000      NA     NA    1.000    1.000
46               age_M00 ~~                   sex          -0.187 0.054  -3.446  0.001   -0.293   -0.080
47               age_M00 ~~               edyears          -0.128 0.058  -2.188  0.029   -0.242   -0.013
48               age_M00 ~~ vascular_risk_sumcorr           0.093 0.057   1.643  0.100   -0.018    0.205
49               age_M00 ~~                  eTIV           0.054 0.054   0.994  0.320   -0.052    0.160
50                   sex ~~               edyears          -0.244 0.050  -4.850  0.000   -0.343   -0.146
51                   sex ~~ vascular_risk_sumcorr          -0.198 0.052  -3.778  0.000   -0.301   -0.095
52                   sex ~~                  eTIV          -0.665 0.027 -24.937  0.000   -0.717   -0.612
53               edyears ~~ vascular_risk_sumcorr          -0.133 0.052  -2.543  0.011   -0.236   -0.031
54               edyears ~~                  eTIV           0.235 0.049   4.768  0.000    0.139    0.332
55 vascular_risk_sumcorr ~~                  eTIV           0.132 0.061   2.154  0.031    0.012    0.251
```

### BLGCM – Global - Total WMH and Mean Cortical Thickness

```
lavaan 0.6.17 ended normally after 140 iterations

  Estimator                                         ML
  Optimization method                           NLMINB
  Number of model parameters                        59
  Number of equality constraints                    12

  Number of observations                           436
  Number of missing patterns                        51

Model Test User Model:
                                              Standard      Scaled
  Test Statistic                               105.811      82.905
  Degrees of freedom                                88          88
  P-value (Chi-square)                           0.095       0.634
  Scaling correction factor                                  1.276
    Yuan-Bentler correction (Mplus variant)

Model Test Baseline Model:

  Test statistic                              7143.130    5143.761
  Degrees of freedom                               105         105
  P-value                                        0.000       0.000
  Scaling correction factor                                  1.389

User Model versus Baseline Model:

  Comparative Fit Index (CFI)                    0.997       1.000
  Tucker-Lewis Index (TLI)                       0.997       1.001

  Robust Comparative Fit Index (CFI)                         1.000
  Robust Tucker-Lewis Index (TLI)                            1.002

Loglikelihood and Information Criteria:

  Loglikelihood user model (H0)              -3508.336   -3508.336
  Scaling correction factor                                  1.028
      for the MLR correction
  Loglikelihood unrestricted model (H1)      -3455.431   -3455.431
  Scaling correction factor                                  1.281
      for the MLR correction

  Akaike (AIC)                                7110.673    7110.673
  Bayesian (BIC)                              7302.322    7302.322
  Sample-size adjusted Bayesian (SABIC)       7153.169    7153.169

Root Mean Square Error of Approximation:

  RMSEA                                          0.022       0.000
  90 Percent confidence interval - lower         0.000       0.000
  90 Percent confidence interval - upper         0.035       0.021
  P-value H_0: RMSEA <= 0.050                    1.000       1.000
  P-value H_0: RMSEA >= 0.080                    0.000       0.000

  Robust RMSEA                                               0.000
  90 Percent confidence interval - lower                     0.000
  90 Percent confidence interval - upper                     0.031
  P-value H_0: Robust RMSEA <= 0.050                         0.999
  P-value H_0: Robust RMSEA >= 0.080                         0.000

Standardized Root Mean Square Residual:

  SRMR                                           0.017       0.017

Parameter Estimates:

  Standard errors                             Sandwich
  Information bread                           Observed
  Observed information based on                Hessian

Latent Variables:
                   Estimate  Std.Err  z-value  P(>|z|)   Std.lv  Std.all
  iy =~
    y1                1.000                               0.963    0.992
    y2                1.000                               0.963    0.986
    y3                1.000                               0.963    0.976
    y4                1.000                               0.963    0.963
    y5                1.000                               0.963    0.946
  sy =~
    y1                0.000                               0.000    0.000
    y2                1.000                               0.064    0.066
    y3                2.000                               0.129    0.130
    y4                3.000                               0.193    0.193
    y5                4.000                               0.257    0.253
  ix =~
    x1                1.000                               0.510    0.933
    x2                1.000                               0.510    0.926
    x3                1.000                               0.510    0.909
    x4                1.000                               0.510    0.886
    x5                1.000                               0.510    0.857
  sx =~
    x1                0.000                               0.000    0.000
    x2                1.000                               0.054    0.099
    x3                2.000                               0.109    0.194
    x4                3.000                               0.163    0.284
    x5                4.000                               0.218    0.366

Regressions:
                   Estimate  Std.Err  z-value  P(>|z|)   Std.lv  Std.all
  iy ~
    age_M00           0.352    0.044    8.021    0.000    0.365    0.365
    sex               0.168    0.060    2.805    0.005    0.175    0.175
    edyears          -0.035    0.043   -0.814    0.416   -0.037   -0.037
    vsclr_rsk_smcr    0.086    0.044    1.954    0.051    0.090    0.090
    eTIV              0.246    0.056    4.386    0.000    0.256    0.256
  sy ~
    age_M00          -0.004    0.004   -0.882    0.378   -0.059   -0.059
    sex               0.003    0.006    0.456    0.648    0.039    0.039
    edyears          -0.004    0.004   -1.008    0.314   -0.066   -0.066
    vsclr_rsk_smcr    0.001    0.004    0.167    0.867    0.011    0.011
    eTIV              0.001    0.005    0.301    0.763    0.022    0.022
  ix ~
    age_M00          -0.191    0.024   -8.109    0.000   -0.375   -0.375
    sex              -0.008    0.030   -0.254    0.799   -0.015   -0.015
    edyears          -0.019    0.025   -0.761    0.447   -0.038   -0.038
    vsclr_rsk_smcr   -0.039    0.025   -1.577    0.115   -0.076   -0.076
    eTIV             -0.104    0.030   -3.467    0.001   -0.204   -0.204
  sx ~
    age_M00          -0.008    0.005   -1.747    0.081   -0.147   -0.147
    sex               0.006    0.006    1.000    0.317    0.103    0.103
    edyears           0.004    0.004    0.877    0.381    0.069    0.069
    vsclr_rsk_smcr    0.000    0.005    0.043    0.966    0.004    0.004
    eTIV              0.007    0.005    1.424    0.154    0.134    0.134

Covariances:
                           Estimate  Std.Err  z-value  P(>|z|)   Std.lv  Std.all
 .iy ~~
   .sy                        0.004    0.004    1.104    0.270    0.075    0.075
 .ix ~~
   .sx                       -0.000    0.003   -0.090    0.928   -0.011   -0.011
 .y1 ~~
   .x1      (tht_)           -0.001    0.001   -0.723    0.469   -0.001   -0.022
 .y2 ~~
   .x2      (tht_)           -0.001    0.001   -0.723    0.469   -0.001   -0.022
 .y3 ~~
   .x3      (tht_)           -0.001    0.001   -0.723    0.469   -0.001   -0.022
 .y4 ~~
   .x4      (tht_)           -0.001    0.001   -0.723    0.469   -0.001   -0.022
 .y5 ~~
   .x5      (tht_)           -0.001    0.001   -0.723    0.469   -0.001   -0.022
 .iy ~~
   .ix                       -0.063    0.020   -3.202    0.001   -0.160   -0.160
   .sx                       -0.006    0.003   -1.654    0.098   -0.121   -0.121
 .sy ~~
   .ix                       -0.004    0.002   -1.870    0.061   -0.123   -0.123
   .sx                       -0.001    0.000   -1.128    0.259   -0.149   -0.149
  age_M00 ~~
    sex                      -0.168    0.047   -3.598    0.000   -0.168   -0.168
    edyears                  -0.128    0.050   -2.558    0.011   -0.128   -0.128
    vsclr__                   0.140    0.048    2.931    0.003    0.140    0.140
    eTIV                      0.072    0.047    1.539    0.124    0.072    0.072
  sex ~~
    edyears                  -0.233    0.044   -5.346    0.000   -0.233   -0.233
    vsclr__                  -0.195    0.045   -4.305    0.000   -0.195   -0.195
    eTIV                     -0.670    0.022  -30.135    0.000   -0.670   -0.670
  edyears ~~
    vsclr__                  -0.154    0.044   -3.502    0.000   -0.154   -0.154
    eTIV                      0.242    0.043    5.627    0.000    0.242    0.242
  vascular_risk_sumcorr ~~
    eTIV                      0.111    0.050    2.220    0.026    0.111    0.111

Intercepts:
                   Estimate  Std.Err  z-value  P(>|z|)   Std.lv  Std.all
   .iy               -0.107    0.042   -2.549    0.011   -0.111   -0.111
   .sy                0.068    0.004   17.325    0.000    1.061    1.061
   .y1                0.000                               0.000    0.000
   .y2                0.000                               0.000    0.000
   .y3                0.000                               0.000    0.000
   .y4                0.000                               0.000    0.000
   .y5                0.000                               0.000    0.000
   .ix                0.109    0.023    4.711    0.000    0.214    0.214
   .sx               -0.015    0.004   -3.333    0.001   -0.272   -0.272
   .x1                0.000                               0.000    0.000
   .x2                0.000                               0.000    0.000
   .x3                0.000                               0.000    0.000
   .x4                0.000                               0.000    0.000
   .x5                0.000                               0.000    0.000
    age_M00           0.000                               0.000    0.000
    edyears           0.000                               0.000    0.000
    sex               0.000                               0.000    0.000
    vsclr_rsk_smcr    0.000                               0.000    0.000
    eTIV              0.000                               0.000    0.000

Variances:
                   Estimate  Std.Err  z-value  P(>|z|)   Std.lv  Std.all
   .iy                0.758    0.048   15.840    0.000    0.818    0.818
   .sy                0.004    0.001    5.504    0.000    0.991    0.991
   .y1     (tht_y)    0.016    0.001   11.058    0.000    0.016    0.017
   .y2     (tht_y)    0.016    0.001   11.058    0.000    0.016    0.017
   .y3     (tht_y)    0.016    0.001   11.058    0.000    0.016    0.016
   .y4     (tht_y)    0.016    0.001   11.058    0.000    0.016    0.016
   .y5     (tht_y)    0.016    0.001   11.058    0.000    0.016    0.015
   .ix                0.207    0.017   12.282    0.000    0.795    0.795
   .sx                0.003    0.001    2.512    0.012    0.958    0.958
   .x1     (tht_x)    0.039    0.004    9.286    0.000    0.039    0.129
   .x2     (tht_x)    0.039    0.004    9.286    0.000    0.039    0.127
   .x3     (tht_x)    0.039    0.004    9.286    0.000    0.039    0.123
   .x4     (tht_x)    0.039    0.004    9.286    0.000    0.039    0.116
   .x5     (tht_x)    0.039    0.004    9.286    0.000    0.039    0.109
    ag_M00            1.000                               1.000    1.000
    sex               1.000                               1.000    1.000
    edyers            1.000                               1.000    1.000
    vscl__            1.000                               1.000    1.000
    eTIV              1.000                               1.000    1.000

R-Square:
                   Estimate
    iy                0.182
    sy                0.009
    y1                0.983
    y2                0.983
    y3                0.984
    y4                0.984
    y5                0.985
    ix                0.205
    sx                0.042
    x1                0.871
    x2                0.873
    x3                0.877
    x4                0.884
    x5                0.891

                     lhs op                   rhs    label est.std    se        z pvalue ci.lower ci.upper
1                     iy =~                    y1            0.992 0.001 1139.777  0.000    0.990    0.993
2                     iy =~                    y2            0.986 0.005  210.119  0.000    0.977    0.995
3                     iy =~                    y3            0.976 0.009  110.687  0.000    0.959    0.994
4                     iy =~                    y4            0.963 0.013   76.235  0.000    0.938    0.988
5                     iy =~                    y5            0.946 0.016   58.806  0.000    0.915    0.978
6                     sy =~                    y1            0.000 0.000       NA     NA    0.000    0.000
7                     sy =~                    y2            0.066 0.006   10.782  0.000    0.054    0.078
8                     sy =~                    y3            0.130 0.012   10.785  0.000    0.107    0.154
9                     sy =~                    y4            0.193 0.018   10.855  0.000    0.158    0.228
10                    sy =~                    y5            0.253 0.023   10.990  0.000    0.208    0.298
11                    iy ~~                    iy            0.818 0.033   25.116  0.000    0.754    0.882
12                    sy ~~                    sy            0.991 0.012   80.820  0.000    0.967    1.015
13                    iy ~~                    sy            0.075 0.070    1.074  0.283   -0.062    0.213
14                    iy ~1                                 -0.111 0.044   -2.517  0.012   -0.198   -0.025
15                    sy ~1                                  1.061 0.100   10.602  0.000    0.865    1.257
16                    y1 ~~                    y1  theta_y   0.017 0.002    9.681  0.000    0.013    0.020
17                    y2 ~~                    y2  theta_y   0.017 0.002    9.788  0.000    0.013    0.020
18                    y3 ~~                    y3  theta_y   0.016 0.002    9.826  0.000    0.013    0.019
19                    y4 ~~                    y4  theta_y   0.016 0.002    9.800  0.000    0.013    0.019
20                    y5 ~~                    y5  theta_y   0.015 0.002    9.717  0.000    0.012    0.018
21                    y1 ~1                                  0.000 0.000       NA     NA    0.000    0.000
22                    y2 ~1                                  0.000 0.000       NA     NA    0.000    0.000
23                    y3 ~1                                  0.000 0.000       NA     NA    0.000    0.000
24                    y4 ~1                                  0.000 0.000       NA     NA    0.000    0.000
25                    y5 ~1                                  0.000 0.000       NA     NA    0.000    0.000
26                    iy  ~               age_M00            0.365 0.043    8.464  0.000    0.281    0.450
27                    iy  ~                   sex            0.175 0.062    2.829  0.005    0.054    0.296
28                    iy  ~               edyears           -0.037 0.045   -0.814  0.415   -0.125    0.052
29                    iy  ~ vascular_risk_sumcorr            0.090 0.045    1.978  0.048    0.001    0.179
30                    iy  ~                  eTIV            0.256 0.057    4.531  0.000    0.145    0.367
31                    sy  ~               age_M00           -0.059 0.067   -0.884  0.377   -0.189    0.072
32                    sy  ~                   sex            0.039 0.087    0.452  0.651   -0.131    0.209
33                    sy  ~               edyears           -0.066 0.066   -0.999  0.318   -0.196    0.064
34                    sy  ~ vascular_risk_sumcorr            0.011 0.066    0.167  0.867   -0.119    0.141
35                    sy  ~                  eTIV            0.022 0.074    0.300  0.765   -0.124    0.168
36                    ix =~                    x1            0.933 0.008  113.548  0.000    0.917    0.949
37                    ix =~                    x2            0.926 0.013   73.080  0.000    0.901    0.950
38                    ix =~                    x3            0.909 0.018   50.874  0.000    0.874    0.945
39                    ix =~                    x4            0.886 0.022   39.735  0.000    0.842    0.930
40                    ix =~                    x5            0.857 0.026   32.538  0.000    0.806    0.909
41                    sx =~                    x1            0.000 0.000       NA     NA    0.000    0.000
42                    sx =~                    x2            0.099 0.019    5.204  0.000    0.062    0.136
43                    sx =~                    x3            0.194 0.037    5.190  0.000    0.121    0.267
44                    sx =~                    x4            0.284 0.054    5.264  0.000    0.178    0.389
45                    sx =~                    x5            0.366 0.068    5.418  0.000    0.234    0.498
46                    ix ~~                    ix            0.795 0.036   21.981  0.000    0.724    0.866
47                    sx ~~                    sx            0.958 0.035   27.607  0.000    0.890    1.026
48                    ix ~~                    sx           -0.011 0.115   -0.092  0.927   -0.236    0.215
49                    ix ~1                                  0.214 0.046    4.611  0.000    0.123    0.305
50                    sx ~1                                 -0.272 0.082   -3.298  0.001   -0.433   -0.110
51                    x1 ~~                    x1  theta_x   0.129 0.015    8.413  0.000    0.099    0.159
52                    x2 ~~                    x2  theta_x   0.127 0.015    8.596  0.000    0.098    0.156
53                    x3 ~~                    x3  theta_x   0.123 0.014    8.659  0.000    0.095    0.150
54                    x4 ~~                    x4  theta_x   0.116 0.013    8.630  0.000    0.090    0.143
55                    x5 ~~                    x5  theta_x   0.109 0.013    8.483  0.000    0.084    0.134
56                    x1 ~1                                  0.000 0.000       NA     NA    0.000    0.000
57                    x2 ~1                                  0.000 0.000       NA     NA    0.000    0.000
58                    x3 ~1                                  0.000 0.000       NA     NA    0.000    0.000
59                    x4 ~1                                  0.000 0.000       NA     NA    0.000    0.000
60                    x5 ~1                                  0.000 0.000       NA     NA    0.000    0.000
61                    y1 ~~                    x1 theta_xy  -0.022 0.031   -0.723  0.470   -0.083    0.038
62                    y2 ~~                    x2 theta_xy  -0.022 0.031   -0.723  0.470   -0.083    0.038
63                    y3 ~~                    x3 theta_xy  -0.022 0.031   -0.723  0.470   -0.083    0.038
64                    y4 ~~                    x4 theta_xy  -0.022 0.031   -0.723  0.470   -0.083    0.038
65                    y5 ~~                    x5 theta_xy  -0.022 0.031   -0.723  0.470   -0.083    0.038
66                    ix  ~               age_M00           -0.375 0.042   -8.844  0.000   -0.458   -0.292
67                    ix  ~                   sex           -0.015 0.059   -0.254  0.799   -0.130    0.100
68                    ix  ~               edyears           -0.038 0.050   -0.762  0.446   -0.135    0.060
69                    ix  ~ vascular_risk_sumcorr           -0.076 0.048   -1.589  0.112   -0.170    0.018
70                    ix  ~                  eTIV           -0.204 0.058   -3.543  0.000   -0.317   -0.091
71                    sx  ~               age_M00           -0.147 0.090   -1.641  0.101   -0.323    0.029
72                    sx  ~                   sex            0.103 0.107    0.964  0.335   -0.107    0.313
73                    sx  ~               edyears            0.069 0.082    0.843  0.399   -0.092    0.230
74                    sx  ~ vascular_risk_sumcorr            0.004 0.084    0.043  0.966   -0.161    0.168
75                    sx  ~                  eTIV            0.134 0.094    1.430  0.153   -0.050    0.319
76                    iy ~~                    ix           -0.160 0.049   -3.285  0.001   -0.256   -0.065
77                    iy ~~                    sx           -0.121 0.076   -1.595  0.111   -0.270    0.028
78                    sy ~~                    ix           -0.123 0.063   -1.964  0.049   -0.246    0.000
79                    sy ~~                    sx           -0.149 0.145   -1.026  0.305   -0.432    0.135
```

```
80               age_M00 ~1                                  0.000 0.000       NA     NA    0.000    0.000
81               edyears ~1                                  0.000 0.000       NA     NA    0.000    0.000
82                   sex ~1                                  0.000 0.000       NA     NA    0.000    0.000
83 vascular_risk_sumcorr ~1                                  0.000 0.000       NA     NA    0.000    0.000
84                  eTIV ~1                                  0.000 0.000       NA     NA    0.000    0.000
85               age_M00 ~~               age_M00            1.000 0.000       NA     NA    1.000    1.000
86                   sex ~~                   sex            1.000 0.000       NA     NA    1.000    1.000
87               edyears ~~               edyears            1.000 0.000       NA     NA    1.000    1.000
88 vascular_risk_sumcorr ~~ vascular_risk_sumcorr            1.000 0.000       NA     NA    1.000    1.000
89                  eTIV ~~                  eTIV            1.000 0.000       NA     NA    1.000    1.000
90               age_M00 ~~                   sex           -0.168 0.047   -3.598  0.000   -0.259   -0.076
91               age_M00 ~~               edyears           -0.128 0.050   -2.558  0.011   -0.226   -0.030
92               age_M00 ~~ vascular_risk_sumcorr            0.140 0.048    2.931  0.003    0.046    0.233
93               age_M00 ~~                  eTIV            0.072 0.047    1.539  0.124   -0.020    0.165
94                   sex ~~               edyears           -0.233 0.044   -5.346  0.000   -0.318   -0.147
95                   sex ~~ vascular_risk_sumcorr           -0.195 0.045   -4.305  0.000   -0.283   -0.106
96                   sex ~~                  eTIV           -0.670 0.022  -30.135  0.000   -0.713   -0.626
97               edyears ~~ vascular_risk_sumcorr           -0.154 0.044   -3.502  0.000   -0.240   -0.068
98               edyears ~~                  eTIV            0.242 0.043    5.627  0.000    0.158    0.326
99 vascular_risk_sumcorr ~~                  eTIV            0.111 0.050    2.220  0.026    0.013    0.209
```

### BLGCM – Global - Frontal WMH and Mean Cortical Thickness

### lavaan 0.6.17 ended normally after 131 iterations

```
  Estimator                                         ML
  Optimization method                           NLMINB
  Number of model parameters                        59
  Number of equality constraints                    12

  Number of observations                           416
  Number of missing patterns                        47

Model Test User Model:
                                              Standard      Scaled
  Test Statistic                               101.986      73.100
  Degrees of freedom                                88          88
  P-value (Chi-square)                           0.146       0.873
  Scaling correction factor                                  1.395
    Yuan-Bentler correction (Mplus variant)

Model Test Baseline Model:

  Test statistic                              5948.010    3937.335
  Degrees of freedom                               105         105
  P-value                                        0.000       0.000
  Scaling correction factor                                  1.511

User Model versus Baseline Model:

  Comparative Fit Index (CFI)                    0.998       1.000
  Tucker-Lewis Index (TLI)                       0.997       1.005

  Robust Comparative Fit Index (CFI)                         1.000
  Robust Tucker-Lewis Index (TLI)                            1.006

Loglikelihood and Information Criteria:

  Loglikelihood user model (H0)              -3826.524   -3826.524
  Scaling correction factor                                  1.060
      for the MLR correction
  Loglikelihood unrestricted model (H1)      -3775.531   -3775.531
  Scaling correction factor                                  1.373
      for the MLR correction

  Akaike (AIC)                                7747.049    7747.049
  Bayesian (BIC)                              7936.491    7936.491
  Sample-size adjusted Bayesian (SABIC)       7787.348    7787.348

Root Mean Square Error of Approximation:

  RMSEA                                          0.020       0.000
  90 Percent confidence interval - lower         0.000       0.000
  90 Percent confidence interval - upper         0.034       0.010
  P-value H_0: RMSEA <= 0.050                    1.000       1.000
  P-value H_0: RMSEA >= 0.080                    0.000       0.000

  Robust RMSEA                                               0.000
  90 Percent confidence interval - lower                     0.000
  90 Percent confidence interval - upper                     0.012
  P-value H_0: Robust RMSEA <= 0.050                         1.000
  P-value H_0: Robust RMSEA >= 0.080                         0.000

Standardized Root Mean Square Residual:

  SRMR                                           0.018       0.018

Parameter Estimates:

  Standard errors                             Sandwich
  Information bread                           Observed
  Observed information based on                Hessian

Latent Variables:
                   Estimate  Std.Err  z-value  P(>|z|)   Std.lv  Std.all
  iy =~
    y1                1.000                               0.980    0.982
    y2                1.000                               0.980    0.987
    y3                1.000                               0.980    0.988
    y4                1.000                               0.980    0.986
    y5                1.000                               0.980    0.980
  sy =~
    y1                0.000                               0.000    0.000
    y2                1.000                               0.060    0.061
    y3                2.000                               0.120    0.122
    y4                3.000                               0.181    0.182
    y5                4.000                               0.241    0.241
  ix =~
    x1                1.000                               0.511    0.930
    x2                1.000                               0.511    0.924
    x3                1.000                               0.511    0.909
    x4                1.000                               0.511    0.885
    x5                1.000                               0.511    0.853
  sx =~
    x1                0.000                               0.000    0.000
    x2                1.000                               0.060    0.108
    x3                2.000                               0.119    0.212
    x4                3.000                               0.179    0.310
    x5                4.000                               0.239    0.398

Regressions:
                   Estimate  Std.Err  z-value  P(>|z|)   Std.lv  Std.all
  iy ~
    age_M00           0.295    0.048    6.120    0.000    0.301    0.301
    sex               0.184    0.063    2.925    0.003    0.188    0.188
    edyears          -0.023    0.045   -0.503    0.615   -0.023   -0.023
    vsclr_rsk_smcr    0.075    0.048    1.581    0.114    0.077    0.077
    eTIV              0.253    0.060    4.193    0.000    0.258    0.258
  sy ~
    age_M00          -0.005    0.005   -1.011    0.312   -0.083   -0.083
    sex              -0.001    0.007   -0.108    0.914   -0.012   -0.012
    edyears          -0.005    0.005   -1.075    0.282   -0.087   -0.087
    vsclr_rsk_smcr    0.004    0.005    0.791    0.429    0.070    0.070
    eTIV             -0.003    0.006   -0.607    0.544   -0.058   -0.058
  ix ~
    age_M00          -0.181    0.024   -7.384    0.000   -0.353   -0.353
    sex              -0.009    0.031   -0.292    0.770   -0.018   -0.018
    edyears          -0.022    0.026   -0.835    0.404   -0.042   -0.042
    vsclr_rsk_smcr   -0.038    0.026   -1.480    0.139   -0.074   -0.074
    eTIV             -0.113    0.031   -3.664    0.000   -0.222   -0.222
  sx ~
    age_M00          -0.010    0.005   -1.922    0.055   -0.162   -0.162
    sex               0.009    0.006    1.447    0.148    0.143    0.143
    edyears           0.003    0.004    0.792    0.429    0.058    0.058
    vsclr_rsk_smcr   -0.001    0.005   -0.198    0.843   -0.016   -0.016
    eTIV              0.010    0.005    1.948    0.051    0.175    0.175

Covariances:
                           Estimate  Std.Err  z-value  P(>|z|)   Std.lv  Std.all
 .iy ~~
   .sy                       -0.006    0.005   -1.202    0.229   -0.102   -0.102
 .ix ~~
   .sx                       -0.001    0.003   -0.481    0.630   -0.056   -0.056
 .y1 ~~
   .x1      (tht_)           -0.002    0.001   -1.173    0.241   -0.002   -0.045
 .y2 ~~
   .x2      (tht_)           -0.002    0.001   -1.173    0.241   -0.002   -0.045
 .y3 ~~
   .x3      (tht_)           -0.002    0.001   -1.173    0.241   -0.002   -0.045
 .y4 ~~
   .x4      (tht_)           -0.002    0.001   -1.173    0.241   -0.002   -0.045
 .y5 ~~
   .x5      (tht_)           -0.002    0.001   -1.173    0.241   -0.002   -0.045
 .iy ~~
   .ix                       -0.059    0.020   -2.896    0.004   -0.141   -0.141
   .sx                       -0.003    0.004   -0.744    0.457   -0.062   -0.062
 .sy ~~
   .ix                        0.000    0.002    0.009    0.993    0.001    0.001
   .sx                       -0.001    0.000   -2.059    0.040   -0.269   -0.269
  age_M00 ~~
    sex                      -0.165    0.048   -3.439    0.001   -0.165   -0.165
    edyears                  -0.134    0.051   -2.609    0.009   -0.134   -0.134
    vsclr__                   0.135    0.048    2.791    0.005    0.135    0.135
    eTIV                      0.058    0.048    1.195    0.232    0.058    0.058
  sex ~~
    edyears                  -0.228    0.045   -5.111    0.000   -0.228   -0.228
    vsclr__                  -0.206    0.046   -4.491    0.000   -0.206   -0.206
    eTIV                     -0.672    0.023  -29.509    0.000   -0.672   -0.672
  edyears ~~
    vsclr__                  -0.148    0.045   -3.292    0.001   -0.148   -0.148
    eTIV                      0.248    0.044    5.690    0.000    0.248    0.248
  vascular_risk_sumcorr ~~
    eTIV                      0.119    0.051    2.330    0.020    0.119    0.119

Intercepts:
                   Estimate  Std.Err  z-value  P(>|z|)   Std.lv  Std.all
   .iy               -0.122    0.045   -2.679    0.007   -0.124   -0.124
   .sy                0.074    0.005   16.002    0.000    1.221    1.221
   .y1                0.000                               0.000    0.000
   .y2                0.000                               0.000    0.000
   .y3                0.000                               0.000    0.000
   .y4                0.000                               0.000    0.000
   .y5                0.000                               0.000    0.000
   .ix                0.102    0.024    4.249    0.000    0.200    0.200
   .sx               -0.014    0.005   -2.886    0.004   -0.230   -0.230
   .x1                0.000                               0.000    0.000
   .x2                0.000                               0.000    0.000
   .x3                0.000                               0.000    0.000
   .x4                0.000                               0.000    0.000
   .x5                0.000                               0.000    0.000
    age_M00           0.000                               0.000    0.000
    edyears           0.000                               0.000    0.000
    sex               0.000                               0.000    0.000
    vsclr_rsk_smcr    0.000                               0.000    0.000
    eTIV              0.000                               0.000    0.000

Variances:
                   Estimate  Std.Err  z-value  P(>|z|)   Std.lv  Std.all
   .iy                0.835    0.052   16.206    0.000    0.869    0.869
   .sy                0.004    0.001    4.056    0.000    0.978    0.978
   .y1     (tht_y)    0.035    0.004    9.643    0.000    0.035    0.035
   .y2     (tht_y)    0.035    0.004    9.643    0.000    0.035    0.036
   .y3     (tht_y)    0.035    0.004    9.643    0.000    0.035    0.036
   .y4     (tht_y)    0.035    0.004    9.643    0.000    0.035    0.036
   .y5     (tht_y)    0.035    0.004    9.643    0.000    0.035    0.035
   .ix                0.211    0.018   12.056    0.000    0.807    0.807
   .sx                0.003    0.001    2.701    0.007    0.943    0.943
   .x1     (tht_x)    0.041    0.005    8.924    0.000    0.041    0.136
   .x2     (tht_x)    0.041    0.005    8.924    0.000    0.041    0.134
   .x3     (tht_x)    0.041    0.005    8.924    0.000    0.041    0.130
   .x4     (tht_x)    0.041    0.005    8.924    0.000    0.041    0.123
   .x5     (tht_x)    0.041    0.005    8.924    0.000    0.041    0.114
    ag_M00            1.000                               1.000    1.000
    sex               1.000                               1.000    1.000
    edyers            1.000                               1.000    1.000
    vscl__            1.000                               1.000    1.000
    eTIV              1.000                               1.000    1.000

R-Square:
                   Estimate
    iy                0.131
    sy                0.022
    y1                0.965
    y2                0.964
    y3                0.964
    y4                0.964
    y5                0.965
    ix                0.193
    sx                0.057
    x1                0.864
    x2                0.866
    x3                0.870
    x4                0.877
    x5                0.886

                     lhs op                   rhs    label est.std    se       z pvalue ci.lower ci.upper
1                     iy =~                    y1            0.982 0.002 489.802  0.000    0.978    0.986
2                     iy =~                    y2            0.987 0.006 174.712  0.000    0.976    0.998
3                     iy =~                    y3            0.988 0.010  98.029  0.000    0.969    1.008
4                     iy =~                    y4            0.986 0.014  68.215  0.000    0.958    1.014
5                     iy =~                    y5            0.980 0.019  52.426  0.000    0.944    1.017
6                     sy =~                    y1            0.000 0.000      NA     NA    0.000    0.000
7                     sy =~                    y2            0.061 0.008   7.778  0.000    0.045    0.076
8                     sy =~                    y3            0.122 0.016   7.754  0.000    0.091    0.152
9                     sy =~                    y4            0.182 0.023   7.777  0.000    0.136    0.228
10                    sy =~                    y5            0.241 0.031   7.847  0.000    0.181    0.301
11                    iy ~~                    iy            0.869 0.031  28.242  0.000    0.809    0.930
12                    sy ~~                    sy            0.978 0.023  42.198  0.000    0.933    1.023
13                    iy ~~                    sy           -0.102 0.080  -1.276  0.202   -0.258    0.055
14                    iy ~1                                 -0.124 0.046  -2.672  0.008   -0.215   -0.033
15                    sy ~1                                  1.221 0.162   7.543  0.000    0.904    1.538
16                    y1 ~~                    y1  theta_y   0.035 0.004   8.969  0.000    0.028    0.043
17                    y2 ~~                    y2  theta_y   0.036 0.004   9.185  0.000    0.028    0.043
18                    y3 ~~                    y3  theta_y   0.036 0.004   9.368  0.000    0.028    0.043
19                    y4 ~~                    y4  theta_y   0.036 0.004   9.511  0.000    0.028    0.043
20                    y5 ~~                    y5  theta_y   0.035 0.004   9.602  0.000    0.028    0.042
21                    y1 ~1                                  0.000 0.000      NA     NA    0.000    0.000
22                    y2 ~1                                  0.000 0.000      NA     NA    0.000    0.000
23                    y3 ~1                                  0.000 0.000      NA     NA    0.000    0.000
24                    y4 ~1                                  0.000 0.000      NA     NA    0.000    0.000
25                    y5 ~1                                  0.000 0.000      NA     NA    0.000    0.000
26                    iy  ~               age_M00            0.301 0.047   6.356  0.000    0.208    0.394
27                    iy  ~                   sex            0.188 0.064   2.956  0.003    0.063    0.313
28                    iy  ~               edyears           -0.023 0.046  -0.502  0.615   -0.114    0.067
29                    iy  ~ vascular_risk_sumcorr            0.077 0.048   1.593  0.111   -0.018    0.171
30                    iy  ~                  eTIV            0.258 0.060   4.326  0.000    0.141    0.375
31                    sy  ~               age_M00           -0.083 0.081  -1.032  0.302   -0.242    0.075
32                    sy  ~                   sex           -0.012 0.108  -0.109  0.913   -0.223    0.200
33                    sy  ~               edyears           -0.087 0.079  -1.107  0.269   -0.242    0.067
34                    sy  ~ vascular_risk_sumcorr            0.070 0.087   0.810  0.418   -0.100    0.240
35                    sy  ~                  eTIV           -0.058 0.096  -0.606  0.545   -0.246    0.130
36                    ix =~                    x1            0.930 0.009 104.045  0.000    0.912    0.947
37                    ix =~                    x2            0.924 0.013  72.122  0.000    0.899    0.950
38                    ix =~                    x3            0.909 0.018  50.225  0.000    0.873    0.944
39                    ix =~                    x4            0.885 0.023  38.833  0.000    0.840    0.929
40                    ix =~                    x5            0.853 0.027  31.333  0.000    0.800    0.907
41                    sx =~                    x1            0.000 0.000      NA     NA    0.000    0.000
42                    sx =~                    x2            0.108 0.020   5.520  0.000    0.070    0.146
43                    sx =~                    x3            0.212 0.039   5.510  0.000    0.137    0.288
44                    sx =~                    x4            0.310 0.055   5.612  0.000    0.202    0.418
45                    sx =~                    x5            0.398 0.069   5.816  0.000    0.264    0.533
46                    ix ~~                    ix            0.807 0.037  21.789  0.000    0.734    0.879
47                    sx ~~                    sx            0.943 0.038  24.498  0.000    0.868    1.018
48                    ix ~~                    sx           -0.056 0.109  -0.516  0.606   -0.269    0.157
49                    ix ~1                                  0.200 0.048   4.174  0.000    0.106    0.293
50                    sx ~1                                 -0.230 0.086  -2.683  0.007   -0.398   -0.062
51                    x1 ~~                    x1  theta_x   0.136 0.017   8.167  0.000    0.103    0.168
52                    x2 ~~                    x2  theta_x   0.134 0.016   8.231  0.000    0.102    0.166
53                    x3 ~~                    x3  theta_x   0.130 0.016   8.308  0.000    0.099    0.160
54                    x4 ~~                    x4  theta_x   0.123 0.015   8.416  0.000    0.094    0.151
55                    x5 ~~                    x5  theta_x   0.114 0.013   8.484  0.000    0.088    0.141
56                    x1 ~1                                  0.000 0.000      NA     NA    0.000    0.000
57                    x2 ~1                                  0.000 0.000      NA     NA    0.000    0.000
58                    x3 ~1                                  0.000 0.000      NA     NA    0.000    0.000
59                    x4 ~1                                  0.000 0.000      NA     NA    0.000    0.000
60                    x5 ~1                                  0.000 0.000      NA     NA    0.000    0.000
61                    y1 ~~                    x1 theta_xy  -0.045 0.038  -1.171  0.242   -0.119    0.030
62                    y2 ~~                    x2 theta_xy  -0.045 0.038  -1.171  0.242   -0.119    0.030
63                    y3 ~~                    x3 theta_xy  -0.045 0.038  -1.171  0.242   -0.119    0.030
64                    y4 ~~                    x4 theta_xy  -0.045 0.038  -1.171  0.242   -0.119    0.030
65                    y5 ~~                    x5 theta_xy  -0.045 0.038  -1.171  0.242   -0.119    0.030
66                    ix  ~               age_M00           -0.353 0.044  -8.006  0.000   -0.440   -0.267
67                    ix  ~                   sex           -0.018 0.060  -0.292  0.770   -0.136    0.101
68                    ix  ~               edyears           -0.042 0.051  -0.836  0.403   -0.141    0.057
69                    ix  ~ vascular_risk_sumcorr           -0.074 0.050  -1.493  0.135   -0.171    0.023
70                    ix  ~                  eTIV           -0.222 0.059  -3.753  0.000   -0.338   -0.106
71                    sx  ~               age_M00           -0.162 0.085  -1.907  0.057   -0.329    0.005
72                    sx  ~                   sex            0.143 0.101   1.415  0.157   -0.055    0.341
73                    sx  ~               edyears            0.058 0.076   0.770  0.441   -0.090    0.207
74                    sx  ~ vascular_risk_sumcorr           -0.016 0.082  -0.200  0.842   -0.177    0.144
75                    sx  ~                  eTIV            0.175 0.088   1.993  0.046    0.003    0.348
76                    iy ~~                    ix           -0.141 0.048  -2.958  0.003   -0.235   -0.048
77                    iy ~~                    sx           -0.062 0.086  -0.723  0.470   -0.230    0.106
78                    sy ~~                    ix            0.001 0.080   0.009  0.993   -0.156    0.157
79                    sy ~~                    sx           -0.269 0.145  -1.861  0.063   -0.552    0.014
80               age_M00 ~1                                  0.000 0.000      NA     NA    0.000    0.000
81               edyears ~1                                  0.000 0.000      NA     NA    0.000    0.000
82                   sex ~1                                  0.000 0.000      NA     NA    0.000    0.000
83 vascular_risk_sumcorr ~1                                  0.000 0.000      NA     NA    0.000    0.000
84                  eTIV ~1                                  0.000 0.000      NA     NA    0.000    0.000
85               age_M00 ~~               age_M00            1.000 0.000      NA     NA    1.000    1.000
86                   sex ~~                   sex            1.000 0.000      NA     NA    1.000    1.000
87               edyears ~~               edyears            1.000 0.000      NA     NA    1.000    1.000
88 vascular_risk_sumcorr ~~ vascular_risk_sumcorr            1.000 0.000      NA     NA    1.000    1.000
89                  eTIV ~~                  eTIV            1.000 0.000      NA     NA    1.000    1.000
90               age_M00 ~~                   sex           -0.165 0.048  -3.439  0.001   -0.258   -0.071
91               age_M00 ~~               edyears           -0.134 0.051  -2.609  0.009   -0.235   -0.033
92               age_M00 ~~ vascular_risk_sumcorr            0.135 0.048   2.791  0.005    0.040    0.230
93               age_M00 ~~                  eTIV            0.058 0.048   1.195  0.232   -0.037    0.152
94                   sex ~~               edyears           -0.228 0.045  -5.111  0.000   -0.316   -0.141
95                   sex ~~ vascular_risk_sumcorr           -0.206 0.046  -4.491  0.000   -0.295   -0.116
96                   sex ~~                  eTIV           -0.672 0.023 -29.509  0.000   -0.717   -0.627
97               edyears ~~ vascular_risk_sumcorr           -0.148 0.045  -3.292  0.001   -0.236   -0.060
98               edyears ~~                  eTIV            0.248 0.044   5.690  0.000    0.162    0.333
99 vascular_risk_sumcorr ~~                  eTIV            0.119 0.051   2.330  0.020    0.019    0.21
```

### BLGCM – Global - Occipital WMH and Mean Cortical Thickness

```
lavaan 0.6.17 ended normally after 99 iterations

  Estimator                                         ML
  Optimization method                           NLMINB
  Number of model parameters                        59
  Number of equality constraints                    12

  Number of observations                           390
  Number of missing patterns                        58

Model Test User Model:
                                              Standard      Scaled
  Test Statistic                                94.447      74.736
  Degrees of freedom                                88          88
  P-value (Chi-square)                           0.300       0.842
  Scaling correction factor                                  1.264
    Yuan-Bentler correction (Mplus variant)

Model Test Baseline Model:

  Test statistic                              4558.203    3348.682
  Degrees of freedom                               105         105
  P-value                                        0.000       0.000
  Scaling correction factor                                  1.361

User Model versus Baseline Model:

  Comparative Fit Index (CFI)                    0.999       1.000
  Tucker-Lewis Index (TLI)                       0.998       1.005

  Robust Comparative Fit Index (CFI)                         1.000
  Robust Tucker-Lewis Index (TLI)                            1.004

Loglikelihood and Information Criteria:

  Loglikelihood user model (H0)              -4099.206   -4099.206
  Scaling correction factor                                  1.001
      for the MLR correction
  Loglikelihood unrestricted model (H1)      -4051.983   -4051.983
  Scaling correction factor                                  1.261
      for the MLR correction

  Akaike (AIC)                                8292.412    8292.412
  Bayesian (BIC)                              8478.821    8478.821
  Sample-size adjusted Bayesian (SABIC)       8329.693    8329.693

Root Mean Square Error of Approximation:

  RMSEA                                          0.014       0.000
  90 Percent confidence interval - lower         0.000       0.000
  90 Percent confidence interval - upper         0.032       0.014
  P-value H_0: RMSEA <= 0.050                    1.000       1.000
  P-value H_0: RMSEA >= 0.080                    0.000       0.000

  Robust RMSEA                                               0.000
  90 Percent confidence interval - lower                     0.000
  90 Percent confidence interval - upper                     0.027
  P-value H_0: Robust RMSEA <= 0.050                         1.000
  P-value H_0: Robust RMSEA >= 0.080                         0.000

Standardized Root Mean Square Residual:

  SRMR                                           0.017       0.017

Parameter Estimates:

  Standard errors                             Sandwich
  Information bread                           Observed
  Observed information based on                Hessian

Latent Variables:
                   Estimate  Std.Err  z-value  P(>|z|)   Std.lv  Std.all
  iy =~
    y1                1.000                               0.929    0.957
    y2                1.000                               0.929    0.964
    y3                1.000                               0.929    0.960
    y4                1.000                               0.929    0.945
    y5                1.000                               0.929    0.921
  sy =~
    y1                0.000                               0.000    0.000
    y2                1.000                               0.105    0.109
    y3                2.000                               0.211    0.218
    y4                3.000                               0.316    0.322
    y5                4.000                               0.422    0.418
  ix =~
    x1                1.000                               0.551    0.936
    x2                1.000                               0.551    0.932
    x3                1.000                               0.551    0.920
    x4                1.000                               0.551    0.901
    x5                1.000                               0.551    0.876
  sx =~
    x1                0.000                               0.000    0.000
    x2                1.000                               0.056    0.095
    x3                2.000                               0.112    0.187
    x4                3.000                               0.168    0.275
    x5                4.000                               0.224    0.356

Regressions:
                   Estimate  Std.Err  z-value  P(>|z|)   Std.lv  Std.all
  iy ~
    age_M00           0.297    0.043    6.993    0.000    0.320    0.320
    sex              -0.015    0.065   -0.233    0.816   -0.016   -0.016
    edyears          -0.047    0.049   -0.953    0.341   -0.050   -0.050
    vsclr_rsk_smcr    0.053    0.050    1.060    0.289    0.057    0.057
    eTIV              0.132    0.068    1.949    0.051    0.142    0.142
  sy ~
    age_M00          -0.000    0.008   -0.046    0.963   -0.003   -0.003
    sex               0.023    0.010    2.441    0.015    0.221    0.221
    edyears          -0.004    0.009   -0.483    0.629   -0.042   -0.042
    vsclr_rsk_smcr   -0.011    0.008   -1.425    0.154   -0.106   -0.106
    eTIV              0.038    0.010    3.710    0.000    0.356    0.356
  ix ~
    age_M00          -0.208    0.026   -7.935    0.000   -0.378   -0.378
    sex              -0.026    0.034   -0.776    0.437   -0.048   -0.048
    edyears          -0.022    0.029   -0.777    0.437   -0.040   -0.040
    vsclr_rsk_smcr   -0.031    0.028   -1.118    0.264   -0.056   -0.056
    eTIV             -0.123    0.034   -3.654    0.000   -0.223   -0.223
  sx ~
    age_M00          -0.006    0.005   -1.246    0.213   -0.111   -0.111
    sex               0.005    0.006    0.829    0.407    0.088    0.088
    edyears           0.004    0.005    0.815    0.415    0.069    0.069
    vsclr_rsk_smcr   -0.000    0.005   -0.008    0.993   -0.001   -0.001
    eTIV              0.007    0.005    1.317    0.188    0.129    0.129

Covariances:
                           Estimate  Std.Err  z-value  P(>|z|)   Std.lv  Std.all
 .iy ~~
   .sy                       -0.014    0.008   -1.839    0.066   -0.162   -0.162
 .ix ~~
   .sx                       -0.001    0.003   -0.283    0.777   -0.036   -0.036
 .y1 ~~
   .x1      (tht_)           -0.002    0.002   -1.065    0.287   -0.002   -0.040
 .y2 ~~
   .x2      (tht_)           -0.002    0.002   -1.065    0.287   -0.002   -0.040
 .y3 ~~
   .x3      (tht_)           -0.002    0.002   -1.065    0.287   -0.002   -0.040
 .y4 ~~
   .x4      (tht_)           -0.002    0.002   -1.065    0.287   -0.002   -0.040
 .y5 ~~
   .x5      (tht_)           -0.002    0.002   -1.065    0.287   -0.002   -0.040
 .iy ~~
   .ix                       -0.071    0.024   -2.985    0.003   -0.169   -0.169
   .sx                       -0.010    0.004   -2.221    0.026   -0.210   -0.210
 .sy ~~
   .ix                       -0.009    0.004   -2.330    0.020   -0.183   -0.183
   .sx                        0.001    0.001    0.815    0.415    0.093    0.093
  age_M00 ~~
    sex                      -0.186    0.049   -3.804    0.000   -0.186   -0.186
    edyears                  -0.097    0.053   -1.842    0.066   -0.097   -0.097
    vsclr__                   0.123    0.051    2.417    0.016    0.123    0.123
    eTIV                      0.097    0.049    1.966    0.049    0.097    0.097
  sex ~~
    edyears                  -0.219    0.047   -4.711    0.000   -0.219   -0.219
    vsclr__                  -0.186    0.048   -3.854    0.000   -0.186   -0.186
    eTIV                     -0.675    0.023  -28.872    0.000   -0.675   -0.675
  edyears ~~
    vsclr__                  -0.143    0.047   -3.054    0.002   -0.143   -0.143
    eTIV                      0.238    0.046    5.224    0.000    0.238    0.238
  vascular_risk_sumcorr ~~
    eTIV                      0.102    0.053    1.926    0.054    0.102    0.102

Intercepts:
                   Estimate  Std.Err  z-value  P(>|z|)   Std.lv  Std.all
   .iy               -0.102    0.045   -2.258    0.024   -0.109   -0.109
   .sy                0.058    0.008    7.471    0.000    0.549    0.549
   .y1                0.000                               0.000    0.000
   .y2                0.000                               0.000    0.000
   .y3                0.000                               0.000    0.000
   .y4                0.000                               0.000    0.000
   .y5                0.000                               0.000    0.000
   .ix                0.103    0.026    3.880    0.000    0.186    0.186
   .sx               -0.016    0.005   -3.258    0.001   -0.285   -0.285
   .x1                0.000                               0.000    0.000
   .x2                0.000                               0.000    0.000
   .x3                0.000                               0.000    0.000
   .x4                0.000                               0.000    0.000
   .x5                0.000                               0.000    0.000
    age_M00           0.000                               0.000    0.000
    edyears           0.000                               0.000    0.000
    sex               0.000                               0.000    0.000
    vsclr_rsk_smcr    0.000                               0.000    0.000
    eTIV              0.000                               0.000    0.000

Variances:
                   Estimate  Std.Err  z-value  P(>|z|)   Std.lv  Std.all
   .iy                0.735    0.053   13.925    0.000    0.851    0.851
   .sy                0.010    0.002    5.094    0.000    0.921    0.921
   .y1     (tht_y)    0.080    0.009    9.166    0.000    0.080    0.084
   .y2     (tht_y)    0.080    0.009    9.166    0.000    0.080    0.086
   .y3     (tht_y)    0.080    0.009    9.166    0.000    0.080    0.085
   .y4     (tht_y)    0.080    0.009    9.166    0.000    0.080    0.082
   .y5     (tht_y)    0.080    0.009    9.166    0.000    0.080    0.078
   .ix                0.243    0.021   11.663    0.000    0.798    0.798
   .sx                0.003    0.001    2.334    0.020    0.970    0.970
   .x1     (tht_x)    0.043    0.005    9.405    0.000    0.043    0.124
   .x2     (tht_x)    0.043    0.005    9.405    0.000    0.043    0.123
   .x3     (tht_x)    0.043    0.005    9.405    0.000    0.043    0.119
   .x4     (tht_x)    0.043    0.005    9.405    0.000    0.043    0.115
   .x5     (tht_x)    0.043    0.005    9.405    0.000    0.043    0.108
    ag_M00            1.000                               1.000    1.000
    sex               1.000                               1.000    1.000
    edyers            1.000                               1.000    1.000
    vscl__            1.000                               1.000    1.000
    eTIV              1.000                               1.000    1.000

R-Square:
                   Estimate
    iy                0.149
    sy                0.079
    y1                0.916
    y2                0.914
    y3                0.915
    y4                0.918
    y5                0.922
    ix                0.202
    sx                0.030
    x1                0.876
    x2                0.877
    x3                0.881
    x4                0.885
    x5                0.892

                     lhs op                   rhs    label est.std    se       z pvalue ci.lower ci.upper
1                     iy =~                    y1            0.957 0.005 174.269  0.000    0.946    0.968
2                     iy =~                    y2            0.964 0.011  91.422  0.000    0.944    0.985
3                     iy =~                    y3            0.960 0.017  57.263  0.000    0.927    0.993
4                     iy =~                    y4            0.945 0.022  42.399  0.000    0.901    0.989
5                     iy =~                    y5            0.921 0.027  34.192  0.000    0.868    0.973
6                     sy =~                    y1            0.000 0.000      NA     NA    0.000    0.000
7                     sy =~                    y2            0.109 0.011   9.545  0.000    0.087    0.132
8                     sy =~                    y3            0.218 0.023   9.451  0.000    0.173    0.263
9                     sy =~                    y4            0.322 0.034   9.538  0.000    0.256    0.388
10                    sy =~                    y5            0.418 0.043   9.796  0.000    0.334    0.502
11                    iy ~~                    iy            0.851 0.032  26.280  0.000    0.787    0.914
12                    sy ~~                    sy            0.921 0.040  23.169  0.000    0.843    0.999
13                    iy ~~                    sy           -0.162 0.078  -2.074  0.038   -0.316   -0.009
14                    iy ~1                                 -0.109 0.049  -2.236  0.025   -0.205   -0.014
15                    sy ~1                                  0.549 0.085   6.448  0.000    0.382    0.716
16                    y1 ~~                    y1  theta_y   0.084 0.011   8.031  0.000    0.064    0.105
17                    y2 ~~                    y2  theta_y   0.086 0.010   8.238  0.000    0.065    0.106
18                    y3 ~~                    y3  theta_y   0.085 0.010   8.301  0.000    0.065    0.105
19                    y4 ~~                    y4  theta_y   0.082 0.010   8.229  0.000    0.063    0.102
20                    y5 ~~                    y5  theta_y   0.078 0.010   8.043  0.000    0.059    0.097
21                    y1 ~1                                  0.000 0.000      NA     NA    0.000    0.000
22                    y2 ~1                                  0.000 0.000      NA     NA    0.000    0.000
23                    y3 ~1                                  0.000 0.000      NA     NA    0.000    0.000
24                    y4 ~1                                  0.000 0.000      NA     NA    0.000    0.000
25                    y5 ~1                                  0.000 0.000      NA     NA    0.000    0.000
26                    iy  ~               age_M00            0.320 0.043   7.484  0.000    0.236    0.404
27                    iy  ~                   sex           -0.016 0.070  -0.233  0.816   -0.153    0.121
28                    iy  ~               edyears           -0.050 0.053  -0.954  0.340   -0.154    0.053
29                    iy  ~ vascular_risk_sumcorr            0.057 0.054   1.067  0.286   -0.048    0.162
30                    iy  ~                  eTIV            0.142 0.071   1.983  0.047    0.002    0.282
31                    sy  ~               age_M00           -0.003 0.074  -0.047  0.963   -0.149    0.142
32                    sy  ~                   sex            0.221 0.087   2.523  0.012    0.049    0.392
33                    sy  ~               edyears           -0.042 0.086  -0.489  0.625   -0.210    0.126
34                    sy  ~ vascular_risk_sumcorr           -0.106 0.072  -1.473  0.141   -0.248    0.035
35                    sy  ~                  eTIV            0.356 0.088   4.064  0.000    0.184    0.528
36                    ix =~                    x1            0.936 0.008 118.517  0.000    0.921    0.952
37                    ix =~                    x2            0.932 0.012  74.656  0.000    0.908    0.957
38                    ix =~                    x3            0.920 0.018  51.261  0.000    0.885    0.956
39                    ix =~                    x4            0.901 0.023  40.051  0.000    0.857    0.945
40                    ix =~                    x5            0.876 0.027  32.955  0.000    0.824    0.928
41                    sx =~                    x1            0.000 0.000      NA     NA    0.000    0.000
42                    sx =~                    x2            0.095 0.020   4.789  0.000    0.056    0.134
43                    sx =~                    x3            0.187 0.039   4.761  0.000    0.110    0.264
44                    sx =~                    x4            0.275 0.057   4.810  0.000    0.163    0.387
45                    sx =~                    x5            0.356 0.072   4.930  0.000    0.215    0.498
46                    ix ~~                    ix            0.798 0.038  21.172  0.000    0.724    0.872
47                    sx ~~                    sx            0.970 0.030  32.745  0.000    0.912    1.028
48                    ix ~~                    sx           -0.036 0.120  -0.296  0.767   -0.271    0.200
49                    ix ~1                                  0.186 0.049   3.804  0.000    0.090    0.282
50                    sx ~1                                 -0.285 0.090  -3.158  0.002   -0.463   -0.108
51                    x1 ~~                    x1  theta_x   0.124 0.015   8.354  0.000    0.095    0.153
52                    x2 ~~                    x2  theta_x   0.123 0.014   8.487  0.000    0.094    0.151
53                    x3 ~~                    x3  theta_x   0.119 0.014   8.501  0.000    0.092    0.147
54                    x4 ~~                    x4  theta_x   0.115 0.014   8.442  0.000    0.088    0.141
55                    x5 ~~                    x5  theta_x   0.108 0.013   8.292  0.000    0.083    0.134
56                    x1 ~1                                  0.000 0.000      NA     NA    0.000    0.000
57                    x2 ~1                                  0.000 0.000      NA     NA    0.000    0.000
58                    x3 ~1                                  0.000 0.000      NA     NA    0.000    0.000
59                    x4 ~1                                  0.000 0.000      NA     NA    0.000    0.000
60                    x5 ~1                                  0.000 0.000      NA     NA    0.000    0.000
61                    y1 ~~                    x1 theta_xy  -0.040 0.038  -1.074  0.283   -0.114    0.033
62                    y2 ~~                    x2 theta_xy  -0.040 0.038  -1.074  0.283   -0.114    0.033
63                    y3 ~~                    x3 theta_xy  -0.040 0.038  -1.074  0.283   -0.114    0.033
64                    y4 ~~                    x4 theta_xy  -0.040 0.038  -1.074  0.283   -0.114    0.033
65                    y5 ~~                    x5 theta_xy  -0.040 0.038  -1.074  0.283   -0.114    0.033
66                    ix  ~               age_M00           -0.378 0.044  -8.590  0.000   -0.465   -0.292
67                    ix  ~                   sex           -0.048 0.061  -0.779  0.436   -0.168    0.072
68                    ix  ~               edyears           -0.040 0.052  -0.779  0.436   -0.142    0.061
69                    ix  ~ vascular_risk_sumcorr           -0.056 0.050  -1.125  0.260   -0.154    0.042
70                    ix  ~                  eTIV           -0.223 0.059  -3.766  0.000   -0.339   -0.107
71                    sx  ~               age_M00           -0.111 0.094  -1.176  0.240   -0.296    0.074
72                    sx  ~                   sex            0.088 0.109   0.805  0.421   -0.126    0.301
73                    sx  ~               edyears            0.069 0.089   0.778  0.437   -0.106    0.245
74                    sx  ~ vascular_risk_sumcorr           -0.001 0.091  -0.008  0.993   -0.179    0.177
75                    sx  ~                  eTIV            0.129 0.097   1.325  0.185   -0.062    0.320
76                    iy ~~                    ix           -0.169 0.055  -3.045  0.002   -0.277   -0.060
77                    iy ~~                    sx           -0.210 0.079  -2.654  0.008   -0.366   -0.055
78                    sy ~~                    ix           -0.183 0.076  -2.395  0.017   -0.333   -0.033
79                    sy ~~                    sx            0.093 0.112   0.833  0.405   -0.126    0.313
80               age_M00 ~1                                  0.000 0.000      NA     NA    0.000    0.000
81               edyears ~1                                  0.000 0.000      NA     NA    0.000    0.000
82                   sex ~1                                  0.000 0.000      NA     NA    0.000    0.000
83 vascular_risk_sumcorr ~1                                  0.000 0.000      NA     NA    0.000    0.000
84                  eTIV ~1                                  0.000 0.000      NA     NA    0.000    0.000
85               age_M00 ~~               age_M00            1.000 0.000      NA     NA    1.000    1.000
86                   sex ~~                   sex            1.000 0.000      NA     NA    1.000    1.000
87               edyears ~~               edyears            1.000 0.000      NA     NA    1.000    1.000
88 vascular_risk_sumcorr ~~ vascular_risk_sumcorr            1.000 0.000      NA     NA    1.000    1.000
89                  eTIV ~~                  eTIV            1.000 0.000      NA     NA    1.000    1.000
90               age_M00 ~~                   sex           -0.186 0.049  -3.804  0.000   -0.282   -0.090
91               age_M00 ~~               edyears           -0.097 0.053  -1.842  0.066   -0.200    0.006
92               age_M00 ~~ vascular_risk_sumcorr            0.123 0.051   2.417  0.016    0.023    0.223
93               age_M00 ~~                  eTIV            0.097 0.049   1.966  0.049    0.000    0.194
94                   sex ~~               edyears           -0.219 0.047  -4.711  0.000   -0.310   -0.128
95                   sex ~~ vascular_risk_sumcorr           -0.186 0.048  -3.854  0.000   -0.281   -0.092
96                   sex ~~                  eTIV           -0.675 0.023 -28.872  0.000   -0.721   -0.629
97               edyears ~~ vascular_risk_sumcorr           -0.143 0.047  -3.054  0.002   -0.235   -0.051
98               edyears ~~                  eTIV            0.238 0.046   5.224  0.000    0.149    0.328
99 vascular_risk_sumcorr ~~                  eTIV            0.102 0.053   1.926  0.054   -0.002    0.206
```

### BLGCM – Global - Parietal WMH and Mean Cortical Thickness

```
lavaan 0.6.17 ended normally after 135 iterations

  Estimator                                         ML
  Optimization method                           NLMINB
  Number of model parameters                        59
  Number of equality constraints                    12

  Number of observations                           387
  Number of missing patterns                        49

Model Test User Model:
                                              Standard      Scaled
  Test Statistic                               115.800      87.133
  Degrees of freedom                                88          88
  P-value (Chi-square)                           0.025       0.506
  Scaling correction factor                                  1.329
    Yuan-Bentler correction (Mplus variant)

Model Test Baseline Model:

  Test statistic                              5749.251    3950.841
  Degrees of freedom                               105         105
  P-value                                        0.000       0.000
  Scaling correction factor                                  1.455

User Model versus Baseline Model:

  Comparative Fit Index (CFI)                    0.995       1.000
  Tucker-Lewis Index (TLI)                       0.994       1.000

  Robust Comparative Fit Index (CFI)                         1.000
  Robust Tucker-Lewis Index (TLI)                            1.000

Loglikelihood and Information Criteria:

  Loglikelihood user model (H0)              -3485.560   -3485.560
  Scaling correction factor                                  1.062
      for the MLR correction
  Loglikelihood unrestricted model (H1)      -3427.660   -3427.660
  Scaling correction factor                                  1.330
      for the MLR correction

  Akaike (AIC)                                7065.120    7065.120
  Bayesian (BIC)                              7251.166    7251.166
  Sample-size adjusted Bayesian (SABIC)       7102.040    7102.040

Root Mean Square Error of Approximation:

  RMSEA                                          0.029       0.000
  90 Percent confidence interval - lower         0.011       0.000
  90 Percent confidence interval - upper         0.042       0.025
  P-value H_0: RMSEA <= 0.050                    0.997       1.000
  P-value H_0: RMSEA >= 0.080                    0.000       0.000

  Robust RMSEA                                               0.000
  90 Percent confidence interval - lower                     0.000
  90 Percent confidence interval - upper                     0.041
  P-value H_0: Robust RMSEA <= 0.050                         0.991
  P-value H_0: Robust RMSEA >= 0.080                         0.000

Standardized Root Mean Square Residual:

  SRMR                                           0.019       0.019

Parameter Estimates:

  Standard errors                             Sandwich
  Information bread                           Observed
  Observed information based on                Hessian

Latent Variables:
                   Estimate  Std.Err  z-value  P(>|z|)   Std.lv  Std.all
  iy =~
    y1                1.000                               1.023    0.986
    y2                1.000                               1.023    1.010
    y3                1.000                               1.023    1.029
    y4                1.000                               1.023    1.043
    y5                1.000                               1.023    1.051
  sy =~
    y1                0.000                               0.000    0.000
    y2                1.000                               0.077    0.076
    y3                2.000                               0.154    0.155
    y4                3.000                               0.231    0.236
    y5                4.000                               0.309    0.317
  ix =~
    x1                1.000                               0.534    0.938
    x2                1.000                               0.534    0.937
    x3                1.000                               0.534    0.927
    x4                1.000                               0.534    0.910
    x5                1.000                               0.534    0.886
  sx =~
    x1                0.000                               0.000    0.000
    x2                1.000                               0.055    0.096
    x3                2.000                               0.109    0.190
    x4                3.000                               0.164    0.279
    x5                4.000                               0.218    0.362

Regressions:
                   Estimate  Std.Err  z-value  P(>|z|)   Std.lv  Std.all
  iy ~
    age_M00           0.300    0.050    5.989    0.000    0.294    0.294
    sex               0.074    0.069    1.064    0.287    0.072    0.072
    edyears          -0.020    0.051   -0.391    0.696   -0.019   -0.019
    vsclr_rsk_smcr    0.078    0.054    1.448    0.148    0.076    0.076
    eTIV              0.187    0.068    2.758    0.006    0.183    0.183
  sy ~
    age_M00          -0.018    0.006   -3.158    0.002   -0.235   -0.235
    sex               0.003    0.007    0.416    0.677    0.035    0.035
    edyears          -0.013    0.005   -2.452    0.014   -0.168   -0.168
    vsclr_rsk_smcr   -0.001    0.005   -0.247    0.805   -0.017   -0.017
    eTIV             -0.003    0.007   -0.530    0.596   -0.045   -0.045
  ix ~
    age_M00          -0.194    0.026   -7.535    0.000   -0.363   -0.363
    sex              -0.007    0.033   -0.224    0.823   -0.014   -0.014
    edyears          -0.018    0.029   -0.627    0.531   -0.034   -0.034
    vsclr_rsk_smcr   -0.040    0.027   -1.485    0.137   -0.075   -0.075
    eTIV             -0.112    0.033   -3.418    0.001   -0.209   -0.209
  sx ~
    age_M00          -0.008    0.005   -1.565    0.118   -0.140   -0.140
    sex               0.002    0.006    0.326    0.744    0.034    0.034
    edyears           0.004    0.005    0.787    0.431    0.068    0.068
    vsclr_rsk_smcr    0.001    0.005    0.219    0.826    0.019    0.019
    eTIV              0.005    0.005    1.032    0.302    0.095    0.095

Covariances:
                           Estimate  Std.Err  z-value  P(>|z|)   Std.lv  Std.all
 .iy ~~
   .sy                       -0.022    0.006   -3.720    0.000   -0.308   -0.308
 .ix ~~
   .sx                       -0.002    0.003   -0.653    0.514   -0.079   -0.079
 .y1 ~~
   .x1      (tht_)           -0.000    0.001   -0.153    0.879   -0.000   -0.006
 .y2 ~~
   .x2      (tht_)           -0.000    0.001   -0.153    0.879   -0.000   -0.006
 .y3 ~~
   .x3      (tht_)           -0.000    0.001   -0.153    0.879   -0.000   -0.006
 .y4 ~~
   .x4      (tht_)           -0.000    0.001   -0.153    0.879   -0.000   -0.006
 .y5 ~~
   .x5      (tht_)           -0.000    0.001   -0.153    0.879   -0.000   -0.006
 .iy ~~
   .ix                       -0.056    0.025   -2.251    0.024   -0.123   -0.123
   .sx                       -0.004    0.004   -0.929    0.353   -0.075   -0.075
 .sy ~~
   .ix                       -0.005    0.002   -1.972    0.049   -0.136   -0.136
   .sx                       -0.001    0.001   -1.254    0.210   -0.183   -0.183
  age_M00 ~~
    sex                      -0.162    0.050   -3.269    0.001   -0.162   -0.162
    edyears                  -0.129    0.053   -2.433    0.015   -0.129   -0.129
    vsclr__                   0.108    0.050    2.150    0.032    0.108    0.108
    eTIV                      0.066    0.050    1.327    0.185    0.066    0.066
  sex ~~
    edyears                  -0.243    0.046   -5.333    0.000   -0.243   -0.243
    vsclr__                  -0.204    0.048   -4.280    0.000   -0.204   -0.204
    eTIV                     -0.661    0.024  -27.656    0.000   -0.661   -0.661
  edyears ~~
    vsclr__                  -0.141    0.047   -3.010    0.003   -0.141   -0.141
    eTIV                      0.254    0.046    5.575    0.000    0.254    0.254
  vascular_risk_sumcorr ~~
    eTIV                      0.122    0.053    2.273    0.023    0.122    0.122

Intercepts:
                   Estimate  Std.Err  z-value  P(>|z|)   Std.lv  Std.all
   .iy               -0.153    0.049   -3.107    0.002   -0.150   -0.150
   .sy                0.085    0.005   16.374    0.000    1.095    1.095
   .y1                0.000                               0.000    0.000
   .y2                0.000                               0.000    0.000
   .y3                0.000                               0.000    0.000
   .y4                0.000                               0.000    0.000
   .y5                0.000                               0.000    0.000
   .ix                0.093    0.026    3.636    0.000    0.175    0.175
   .sx               -0.015    0.005   -3.197    0.001   -0.274   -0.274
   .x1                0.000                               0.000    0.000
   .x2                0.000                               0.000    0.000
   .x3                0.000                               0.000    0.000
   .x4                0.000                               0.000    0.000
   .x5                0.000                               0.000    0.000
    age_M00           0.000                               0.000    0.000
    edyears           0.000                               0.000    0.000
    sex               0.000                               0.000    0.000
    vsclr_rsk_smcr    0.000                               0.000    0.000
    eTIV              0.000                               0.000    0.000

Variances:
                   Estimate  Std.Err  z-value  P(>|z|)   Std.lv  Std.all
   .iy                0.921    0.059   15.712    0.000    0.880    0.880
   .sy                0.005    0.001    4.340    0.000    0.910    0.910
   .y1     (tht_y)    0.029    0.004    8.146    0.000    0.029    0.027
   .y2     (tht_y)    0.029    0.004    8.146    0.000    0.029    0.028
   .y3     (tht_y)    0.029    0.004    8.146    0.000    0.029    0.029
   .y4     (tht_y)    0.029    0.004    8.146    0.000    0.029    0.030
   .y5     (tht_y)    0.029    0.004    8.146    0.000    0.029    0.031
   .ix                0.229    0.019   11.804    0.000    0.804    0.804
   .sx                0.003    0.001    2.376    0.018    0.966    0.966
   .x1     (tht_x)    0.039    0.004    9.235    0.000    0.039    0.120
   .x2     (tht_x)    0.039    0.004    9.235    0.000    0.039    0.120
   .x3     (tht_x)    0.039    0.004    9.235    0.000    0.039    0.117
   .x4     (tht_x)    0.039    0.004    9.235    0.000    0.039    0.113
   .x5     (tht_x)    0.039    0.004    9.235    0.000    0.039    0.107
    ag_M00            1.000                               1.000    1.000
    sex               1.000                               1.000    1.000
    edyers            1.000                               1.000    1.000
    vscl__            1.000                               1.000    1.000
    eTIV              1.000                               1.000    1.000

R-Square:
                   Estimate
    iy                0.120
    sy                0.090
    y1                0.973
    y2                0.972
    y3                0.971
    y4                0.970
    y5                0.969
    ix                0.196
    sx                0.034
    x1                0.880
    x2                0.880
    x3                0.883
    x4                0.887
    x5                0.893

                     lhs op                   rhs    label est.std    se       z pvalue ci.lower ci.upper
1                     iy =~                    y1            0.986 0.002 588.243  0.000    0.983    0.990
2                     iy =~                    y2            1.010 0.006 159.191  0.000    0.998    1.022
3                     iy =~                    y3            1.029 0.012  84.513  0.000    1.005    1.053
4                     iy =~                    y4            1.043 0.018  58.194  0.000    1.008    1.078
5                     iy =~                    y5            1.051 0.023  44.843  0.000    1.005    1.097
6                     sy =~                    y1            0.000 0.000      NA     NA    0.000    0.000
7                     sy =~                    y2            0.076 0.009   8.526  0.000    0.059    0.094
8                     sy =~                    y3            0.155 0.019   8.355  0.000    0.119    0.192
9                     sy =~                    y4            0.236 0.029   8.263  0.000    0.180    0.292
10                    sy =~                    y5            0.317 0.038   8.254  0.000    0.242    0.392
11                    iy ~~                    iy            0.880 0.032  27.566  0.000    0.817    0.942
12                    sy ~~                    sy            0.910 0.040  22.841  0.000    0.832    0.988
13                    iy ~~                    sy           -0.308 0.066  -4.691  0.000   -0.436   -0.179
14                    iy ~1                                 -0.150 0.049  -3.082  0.002   -0.245   -0.054
15                    sy ~1                                  1.095 0.112   9.735  0.000    0.875    1.316
16                    y1 ~~                    y1  theta_y   0.027 0.003   8.168  0.000    0.021    0.034
17                    y2 ~~                    y2  theta_y   0.028 0.003   8.190  0.000    0.022    0.035
18                    y3 ~~                    y3  theta_y   0.029 0.004   8.113  0.000    0.022    0.037
19                    y4 ~~                    y4  theta_y   0.030 0.004   7.950  0.000    0.023    0.038
20                    y5 ~~                    y5  theta_y   0.031 0.004   7.720  0.000    0.023    0.038
21                    y1 ~1                                  0.000 0.000      NA     NA    0.000    0.000
22                    y2 ~1                                  0.000 0.000      NA     NA    0.000    0.000
23                    y3 ~1                                  0.000 0.000      NA     NA    0.000    0.000
24                    y4 ~1                                  0.000 0.000      NA     NA    0.000    0.000
25                    y5 ~1                                  0.000 0.000      NA     NA    0.000    0.000
26                    iy  ~               age_M00            0.294 0.047   6.218  0.000    0.201    0.386
27                    iy  ~                   sex            0.072 0.068   1.064  0.287   -0.061    0.205
28                    iy  ~               edyears           -0.019 0.050  -0.391  0.696   -0.117    0.078
29                    iy  ~ vascular_risk_sumcorr            0.076 0.052   1.456  0.145   -0.026    0.178
30                    iy  ~                  eTIV            0.183 0.065   2.808  0.005    0.055    0.311
31                    sy  ~               age_M00           -0.235 0.068  -3.446  0.001   -0.368   -0.101
32                    sy  ~                   sex            0.035 0.085   0.415  0.678   -0.131    0.202
33                    sy  ~               edyears           -0.168 0.066  -2.546  0.011   -0.297   -0.039
34                    sy  ~ vascular_risk_sumcorr           -0.017 0.070  -0.248  0.804   -0.154    0.120
35                    sy  ~                  eTIV           -0.045 0.084  -0.538  0.591   -0.209    0.119
36                    ix =~                    x1            0.938 0.008 120.897  0.000    0.923    0.953
37                    ix =~                    x2            0.937 0.012  77.223  0.000    0.913    0.961
38                    ix =~                    x3            0.927 0.018  52.571  0.000    0.893    0.962
39                    ix =~                    x4            0.910 0.023  40.302  0.000    0.866    0.954
40                    ix =~                    x5            0.886 0.027  32.405  0.000    0.833    0.940
41                    sx =~                    x1            0.000 0.000      NA     NA    0.000    0.000
42                    sx =~                    x2            0.096 0.020   4.872  0.000    0.057    0.134
43                    sx =~                    x3            0.190 0.039   4.858  0.000    0.113    0.266
44                    sx =~                    x4            0.279 0.057   4.924  0.000    0.168    0.390
45                    sx =~                    x5            0.362 0.072   5.065  0.000    0.222    0.502
46                    ix ~~                    ix            0.804 0.038  21.304  0.000    0.730    0.878
47                    sx ~~                    sx            0.966 0.031  30.767  0.000    0.904    1.028
48                    ix ~~                    sx           -0.079 0.110  -0.719  0.472   -0.294    0.136
49                    ix ~1                                  0.175 0.049   3.586  0.000    0.079    0.271
50                    sx ~1                                 -0.274 0.088  -3.099  0.002   -0.446   -0.101
51                    x1 ~~                    x1  theta_x   0.120 0.015   8.254  0.000    0.092    0.149
52                    x2 ~~                    x2  theta_x   0.120 0.014   8.373  0.000    0.092    0.148
53                    x3 ~~                    x3  theta_x   0.117 0.014   8.386  0.000    0.090    0.145
54                    x4 ~~                    x4  theta_x   0.113 0.014   8.319  0.000    0.086    0.140
55                    x5 ~~                    x5  theta_x   0.107 0.013   8.144  0.000    0.081    0.133
56                    x1 ~1                                  0.000 0.000      NA     NA    0.000    0.000
57                    x2 ~1                                  0.000 0.000      NA     NA    0.000    0.000
58                    x3 ~1                                  0.000 0.000      NA     NA    0.000    0.000
59                    x4 ~1                                  0.000 0.000      NA     NA    0.000    0.000
60                    x5 ~1                                  0.000 0.000      NA     NA    0.000    0.000
61                    y1 ~~                    x1 theta_xy  -0.006 0.041  -0.153  0.879   -0.086    0.074
62                    y2 ~~                    x2 theta_xy  -0.006 0.041  -0.153  0.879   -0.086    0.074
63                    y3 ~~                    x3 theta_xy  -0.006 0.041  -0.153  0.879   -0.086    0.074
64                    y4 ~~                    x4 theta_xy  -0.006 0.041  -0.153  0.879   -0.086    0.074
65                    y5 ~~                    x5 theta_xy  -0.006 0.041  -0.153  0.879   -0.086    0.074
66                    ix  ~               age_M00           -0.363 0.045  -8.152  0.000   -0.451   -0.276
67                    ix  ~                   sex           -0.014 0.062  -0.224  0.823   -0.135    0.107
68                    ix  ~               edyears           -0.034 0.054  -0.628  0.530   -0.138    0.071
69                    ix  ~ vascular_risk_sumcorr           -0.075 0.050  -1.498  0.134   -0.174    0.023
70                    ix  ~                  eTIV           -0.209 0.060  -3.504  0.000   -0.326   -0.092
71                    sx  ~               age_M00           -0.140 0.094  -1.488  0.137   -0.324    0.044
72                    sx  ~                   sex            0.034 0.106   0.322  0.747   -0.173    0.241
73                    sx  ~               edyears            0.068 0.089   0.756  0.450   -0.108    0.243
74                    sx  ~ vascular_risk_sumcorr            0.019 0.088   0.218  0.828   -0.154    0.192
75                    sx  ~                  eTIV            0.095 0.092   1.031  0.303   -0.086    0.276
76                    iy ~~                    ix           -0.123 0.054  -2.268  0.023   -0.229   -0.017
77                    iy ~~                    sx           -0.075 0.085  -0.888  0.375   -0.242    0.091
78                    sy ~~                    ix           -0.136 0.068  -2.005  0.045   -0.269   -0.003
79                    sy ~~                    sx           -0.183 0.154  -1.190  0.234   -0.484    0.118
80               age_M00 ~1                                  0.000 0.000      NA     NA    0.000    0.000
81               edyears ~1                                  0.000 0.000      NA     NA    0.000    0.000
82                   sex ~1                                  0.000 0.000      NA     NA    0.000    0.000
83 vascular_risk_sumcorr ~1                                  0.000 0.000      NA     NA    0.000    0.000
84                  eTIV ~1                                  0.000 0.000      NA     NA    0.000    0.000
85               age_M00 ~~               age_M00            1.000 0.000      NA     NA    1.000    1.000
86                   sex ~~                   sex            1.000 0.000      NA     NA    1.000    1.000
87               edyears ~~               edyears            1.000 0.000      NA     NA    1.000    1.000
88 vascular_risk_sumcorr ~~ vascular_risk_sumcorr            1.000 0.000      NA     NA    1.000    1.000
89                  eTIV ~~                  eTIV            1.000 0.000      NA     NA    1.000    1.000
90               age_M00 ~~                   sex           -0.162 0.050  -3.269  0.001   -0.260   -0.065
91               age_M00 ~~               edyears           -0.129 0.053  -2.433  0.015   -0.234   -0.025
92               age_M00 ~~ vascular_risk_sumcorr            0.108 0.050   2.150  0.032    0.010    0.206
93               age_M00 ~~                  eTIV            0.066 0.050   1.327  0.185   -0.032    0.164
94                   sex ~~               edyears           -0.243 0.046  -5.333  0.000   -0.332   -0.154
95                   sex ~~ vascular_risk_sumcorr           -0.204 0.048  -4.280  0.000   -0.297   -0.111
96                   sex ~~                  eTIV           -0.661 0.024 -27.656  0.000   -0.708   -0.614
97               edyears ~~ vascular_risk_sumcorr           -0.141 0.047  -3.010  0.003   -0.233   -0.049
98               edyears ~~                  eTIV            0.254 0.046   5.575  0.000    0.165    0.343
99 vascular_risk_sumcorr ~~                  eTIV            0.122 0.053   2.273  0.023    0.017    0.226
```

### BLGCM – Global - Temporal WMH and Mean Cortical Thickness

```
lavaan 0.6.17 ended normally after 111 iterations

  Estimator                                         ML
  Optimization method                           NLMINB
  Number of model parameters                        59
  Number of equality constraints                    12

  Number of observations                           318
  Number of missing patterns                        56

Model Test User Model:
                                              Standard      Scaled
  Test Statistic                               113.604      87.455
  Degrees of freedom                                88          88
  P-value (Chi-square)                           0.034       0.496
  Scaling correction factor                                  1.299
    Yuan-Bentler correction (Mplus variant)

Model Test Baseline Model:

  Test statistic                              3567.100    2524.451
  Degrees of freedom                               105         105
  P-value                                        0.000       0.000
  Scaling correction factor                                  1.413

User Model versus Baseline Model:

  Comparative Fit Index (CFI)                    0.993       1.000
  Tucker-Lewis Index (TLI)                       0.991       1.000

  Robust Comparative Fit Index (CFI)                         0.995
  Robust Tucker-Lewis Index (TLI)                            0.994

Loglikelihood and Information Criteria:

  Loglikelihood user model (H0)              -3310.467   -3310.467
  Scaling correction factor                                  1.047
      for the MLR correction
  Loglikelihood unrestricted model (H1)      -3253.665   -3253.665
  Scaling correction factor                                  1.304
      for the MLR correction

  Akaike (AIC)                                6714.934    6714.934
  Bayesian (BIC)                              6891.751    6891.751
  Sample-size adjusted Bayesian (SABIC)       6742.677    6742.677

Root Mean Square Error of Approximation:

  RMSEA                                          0.030       0.000
  90 Percent confidence interval - lower         0.009       0.000
  90 Percent confidence interval - upper         0.045       0.028
  P-value H_0: RMSEA <= 0.050                    0.988       1.000
  P-value H_0: RMSEA >= 0.080                    0.000       0.000

  Robust RMSEA                                               0.029
  90 Percent confidence interval - lower                     0.000
  90 Percent confidence interval - upper                     0.056
  P-value H_0: Robust RMSEA <= 0.050                         0.886
  P-value H_0: Robust RMSEA >= 0.080                         0.000

Standardized Root Mean Square Residual:

  SRMR                                           0.023       0.023

Parameter Estimates:

  Standard errors                             Sandwich
  Information bread                           Observed
  Observed information based on                Hessian

Latent Variables:
                   Estimate  Std.Err  z-value  P(>|z|)   Std.lv  Std.all
  iy =~
    y1                1.000                               0.969    0.950
    y2                1.000                               0.969    0.967
    y3                1.000                               0.969    0.978
    y4                1.000                               0.969    0.985
    y5                1.000                               0.969    0.985
  sy =~
    y1                0.000                               0.000    0.000
    y2                1.000                               0.075    0.075
    y3                2.000                               0.150    0.152
    y4                3.000                               0.225    0.229
    y5                4.000                               0.300    0.305
  ix =~
    x1                1.000                               0.499    0.933
    x2                1.000                               0.499    0.928
    x3                1.000                               0.499    0.917
    x4                1.000                               0.499    0.900
    x5                1.000                               0.499    0.878
  sx =~
    x1                0.000                               0.000    0.000
    x2                1.000                               0.046    0.086
    x3                2.000                               0.092    0.169
    x4                3.000                               0.138    0.249
    x5                4.000                               0.184    0.324

Regressions:
                   Estimate  Std.Err  z-value  P(>|z|)   Std.lv  Std.all
  iy ~
    age_M00           0.189    0.054    3.489    0.000    0.195    0.195
    sex               0.125    0.077    1.636    0.102    0.129    0.129
    edyears          -0.096    0.056   -1.704    0.088   -0.099   -0.099
    vsclr_rsk_smcr   -0.016    0.060   -0.269    0.788   -0.017   -0.017
    eTIV              0.228    0.075    3.063    0.002    0.235    0.235
  sy ~
    age_M00          -0.015    0.007   -2.000    0.045   -0.194   -0.194
    sex              -0.016    0.010   -1.565    0.118   -0.208   -0.208
    edyears          -0.001    0.008   -0.097    0.923   -0.011   -0.011
    vsclr_rsk_smcr    0.007    0.011    0.645    0.519    0.095    0.095
    eTIV             -0.009    0.009   -0.954    0.340   -0.114   -0.114
  ix ~
    age_M00          -0.184    0.027   -6.773    0.000   -0.369   -0.369
    sex              -0.011    0.034   -0.314    0.754   -0.021   -0.021
    edyears          -0.044    0.028   -1.564    0.118   -0.089   -0.089
    vsclr_rsk_smcr   -0.053    0.029   -1.836    0.066   -0.106   -0.106
    eTIV             -0.097    0.033   -2.961    0.003   -0.195   -0.195
  sx ~
    age_M00          -0.009    0.005   -1.896    0.058   -0.196   -0.196
    sex               0.002    0.005    0.316    0.752    0.037    0.037
    edyears           0.003    0.005    0.676    0.499    0.069    0.069
    vsclr_rsk_smcr    0.003    0.005    0.566    0.571    0.057    0.057
    eTIV              0.004    0.005    0.875    0.382    0.097    0.097

Covariances:
                           Estimate  Std.Err  z-value  P(>|z|)   Std.lv  Std.all
 .iy ~~
   .sy                       -0.017    0.011   -1.520    0.128   -0.253   -0.253
 .ix ~~
   .sx                       -0.001    0.003   -0.217    0.828   -0.036   -0.036
 .y1 ~~
   .x1      (tht_)            0.004    0.003    1.591    0.112    0.004    0.066
 .y2 ~~
   .x2      (tht_)            0.004    0.003    1.591    0.112    0.004    0.066
 .y3 ~~
   .x3      (tht_)            0.004    0.003    1.591    0.112    0.004    0.066
 .y4 ~~
   .x4      (tht_)            0.004    0.003    1.591    0.112    0.004    0.066
 .y5 ~~
   .x5      (tht_)            0.004    0.003    1.591    0.112    0.004    0.066
 .iy ~~
   .ix                       -0.023    0.027   -0.827    0.408   -0.054   -0.054
   .sx                       -0.003    0.006   -0.467    0.641   -0.064   -0.064
 .sy ~~
   .ix                       -0.006    0.004   -1.479    0.139   -0.185   -0.185
   .sx                       -0.001    0.001   -1.524    0.128   -0.369   -0.369
  age_M00 ~~
    sex                      -0.187    0.054   -3.446    0.001   -0.187   -0.187
    edyears                  -0.128    0.058   -2.188    0.029   -0.128   -0.128
    vsclr__                   0.093    0.057    1.643    0.100    0.093    0.093
    eTIV                      0.054    0.054    0.994    0.320    0.054    0.054
  sex ~~
    edyears                  -0.244    0.050   -4.850    0.000   -0.244   -0.244
    vsclr__                  -0.198    0.052   -3.778    0.000   -0.198   -0.198
    eTIV                     -0.665    0.027  -24.937    0.000   -0.665   -0.665
  edyears ~~
    vsclr__                  -0.133    0.052   -2.543    0.011   -0.133   -0.133
    eTIV                      0.235    0.049    4.768    0.000    0.235    0.235
  vascular_risk_sumcorr ~~
    eTIV                      0.132    0.061    2.154    0.031    0.132    0.132

Intercepts:
                   Estimate  Std.Err  z-value  P(>|z|)   Std.lv  Std.all
   .iy               -0.166    0.055   -3.047    0.002   -0.171   -0.171
   .sy                0.072    0.008    8.512    0.000    0.955    0.955
   .y1                0.000                               0.000    0.000
   .y2                0.000                               0.000    0.000
   .y3                0.000                               0.000    0.000
   .y4                0.000                               0.000    0.000
   .y5                0.000                               0.000    0.000
   .ix                0.104    0.027    3.911    0.000    0.208    0.208
   .sx               -0.013    0.005   -2.779    0.005   -0.289   -0.289
   .x1                0.000                               0.000    0.000
   .x2                0.000                               0.000    0.000
   .x3                0.000                               0.000    0.000
   .x4                0.000                               0.000    0.000
   .x5                0.000                               0.000    0.000
    age_M00           0.000                               0.000    0.000
    edyears           0.000                               0.000    0.000
    sex               0.000                               0.000    0.000
    vsclr_rsk_smcr    0.000                               0.000    0.000
    eTIV              0.000                               0.000    0.000

Variances:
                   Estimate  Std.Err  z-value  P(>|z|)   Std.lv  Std.all
   .iy                0.870    0.071   12.240    0.000    0.926    0.926
   .sy                0.005    0.003    1.563    0.118    0.941    0.941
   .y1     (tht_y)    0.101    0.011    9.196    0.000    0.101    0.097
   .y2     (tht_y)    0.101    0.011    9.196    0.000    0.101    0.100
   .y3     (tht_y)    0.101    0.011    9.196    0.000    0.101    0.102
   .y4     (tht_y)    0.101    0.011    9.196    0.000    0.101    0.104
   .y5     (tht_y)    0.101    0.011    9.196    0.000    0.101    0.104
   .ix                0.199    0.019   10.453    0.000    0.799    0.799
   .sx                0.002    0.001    1.578    0.115    0.944    0.944
   .x1     (tht_x)    0.037    0.004    8.335    0.000    0.037    0.130
   .x2     (tht_x)    0.037    0.004    8.335    0.000    0.037    0.129
   .x3     (tht_x)    0.037    0.004    8.335    0.000    0.037    0.126
   .x4     (tht_x)    0.037    0.004    8.335    0.000    0.037    0.121
   .x5     (tht_x)    0.037    0.004    8.335    0.000    0.037    0.115
    ag_M00            1.000                               1.000    1.000
    sex               1.000                               1.000    1.000
    edyers            1.000                               1.000    1.000
    vscl__            1.000                               1.000    1.000
    eTIV              1.000                               1.000    1.000

R-Square:
                   Estimate
    iy                0.074
    sy                0.059
    y1                0.903
    y2                0.900
    y3                0.898
    y4                0.896
    y5                0.896
    ix                0.201
    sx                0.056
    x1                0.870
    x2                0.871
    x3                0.874
    x4                0.879
    x5                0.885

                     lhs op                   rhs    label est.std    se       z pvalue ci.lower ci.upper
1                     iy =~                    y1            0.950 0.006 149.251  0.000    0.938    0.963
2                     iy =~                    y2            0.967 0.014  70.554  0.000    0.940    0.994
3                     iy =~                    y3            0.978 0.022  45.048  0.000    0.936    1.021
4                     iy =~                    y4            0.985 0.029  33.678  0.000    0.927    1.042
5                     iy =~                    y5            0.985 0.037  26.925  0.000    0.913    1.057
6                     sy =~                    y1            0.000 0.000      NA     NA    0.000    0.000
7                     sy =~                    y2            0.075 0.023   3.328  0.001    0.031    0.119
8                     sy =~                    y3            0.152 0.046   3.301  0.001    0.062    0.242
9                     sy =~                    y4            0.229 0.069   3.309  0.001    0.093    0.364
10                    sy =~                    y5            0.305 0.091   3.353  0.001    0.127    0.484
11                    iy ~~                    iy            0.926 0.031  29.999  0.000    0.865    0.986
12                    sy ~~                    sy            0.941 0.066  14.263  0.000    0.811    1.070
13                    iy ~~                    sy           -0.253 0.123  -2.050  0.040   -0.495   -0.011
14                    iy ~1                                 -0.171 0.057  -3.000  0.003   -0.283   -0.059
15                    sy ~1                                  0.955 0.284   3.366  0.001    0.399    1.510
16                    y1 ~~                    y1  theta_y   0.097 0.012   7.984  0.000    0.073    0.120
17                    y2 ~~                    y2  theta_y   0.100 0.012   8.492  0.000    0.077    0.123
18                    y3 ~~                    y3  theta_y   0.102 0.012   8.690  0.000    0.079    0.126
19                    y4 ~~                    y4  theta_y   0.104 0.012   8.521  0.000    0.080    0.128
20                    y5 ~~                    y5  theta_y   0.104 0.013   8.014  0.000    0.078    0.129
21                    y1 ~1                                  0.000 0.000      NA     NA    0.000    0.000
22                    y2 ~1                                  0.000 0.000      NA     NA    0.000    0.000
23                    y3 ~1                                  0.000 0.000      NA     NA    0.000    0.000
24                    y4 ~1                                  0.000 0.000      NA     NA    0.000    0.000
25                    y5 ~1                                  0.000 0.000      NA     NA    0.000    0.000
26                    iy  ~               age_M00            0.195 0.055   3.552  0.000    0.087    0.302
27                    iy  ~                   sex            0.129 0.078   1.656  0.098   -0.024    0.282
28                    iy  ~               edyears           -0.099 0.058  -1.712  0.087   -0.213    0.014
29                    iy  ~ vascular_risk_sumcorr           -0.017 0.062  -0.269  0.788   -0.137    0.104
30                    iy  ~                  eTIV            0.235 0.075   3.154  0.002    0.089    0.382
31                    sy  ~               age_M00           -0.194 0.120  -1.620  0.105   -0.428    0.041
32                    sy  ~                   sex           -0.208 0.144  -1.448  0.148   -0.489    0.073
33                    sy  ~               edyears           -0.011 0.111  -0.096  0.923   -0.228    0.207
34                    sy  ~ vascular_risk_sumcorr            0.095 0.148   0.638  0.523   -0.196    0.385
35                    sy  ~                  eTIV           -0.114 0.126  -0.906  0.365   -0.362    0.133
36                    ix =~                    x1            0.933 0.009  99.727  0.000    0.914    0.951
37                    ix =~                    x2            0.928 0.015  63.241  0.000    0.899    0.957
38                    ix =~                    x3            0.917 0.021  44.731  0.000    0.877    0.957
39                    ix =~                    x4            0.900 0.025  35.592  0.000    0.851    0.950
40                    ix =~                    x5            0.878 0.030  29.481  0.000    0.820    0.937
41                    sx =~                    x1            0.000 0.000      NA     NA    0.000    0.000
42                    sx =~                    x2            0.086 0.026   3.349  0.001    0.036    0.136
43                    sx =~                    x3            0.169 0.051   3.334  0.001    0.070    0.269
44                    sx =~                    x4            0.249 0.074   3.364  0.001    0.104    0.395
45                    sx =~                    x5            0.324 0.094   3.435  0.001    0.139    0.509
46                    ix ~~                    ix            0.799 0.043  18.672  0.000    0.715    0.883
47                    sx ~~                    sx            0.944 0.057  16.444  0.000    0.832    1.057
48                    ix ~~                    sx           -0.036 0.155  -0.228  0.819   -0.340    0.269
49                    ix ~1                                  0.208 0.054   3.823  0.000    0.101    0.315
50                    sx ~1                                 -0.289 0.121  -2.392  0.017   -0.526   -0.052
51                    x1 ~~                    x1  theta_x   0.130 0.017   7.453  0.000    0.096    0.164
52                    x2 ~~                    x2  theta_x   0.129 0.017   7.641  0.000    0.096    0.162
53                    x3 ~~                    x3  theta_x   0.126 0.016   7.668  0.000    0.094    0.158
54                    x4 ~~                    x4  theta_x   0.121 0.016   7.575  0.000    0.090    0.152
55                    x5 ~~                    x5  theta_x   0.115 0.016   7.352  0.000    0.085    0.146
56                    x1 ~1                                  0.000 0.000      NA     NA    0.000    0.000
57                    x2 ~1                                  0.000 0.000      NA     NA    0.000    0.000
58                    x3 ~1                                  0.000 0.000      NA     NA    0.000    0.000
59                    x4 ~1                                  0.000 0.000      NA     NA    0.000    0.000
60                    x5 ~1                                  0.000 0.000      NA     NA    0.000    0.000
61                    y1 ~~                    x1 theta_xy   0.066 0.041   1.631  0.103   -0.013    0.146
62                    y2 ~~                    x2 theta_xy   0.066 0.041   1.631  0.103   -0.013    0.146
63                    y3 ~~                    x3 theta_xy   0.066 0.041   1.631  0.103   -0.013    0.146
64                    y4 ~~                    x4 theta_xy   0.066 0.041   1.631  0.103   -0.013    0.146
65                    y5 ~~                    x5 theta_xy   0.066 0.041   1.631  0.103   -0.013    0.146
66                    ix  ~               age_M00           -0.369 0.050  -7.322  0.000   -0.467   -0.270
67                    ix  ~                   sex           -0.021 0.068  -0.314  0.754   -0.155    0.112
68                    ix  ~               edyears           -0.089 0.056  -1.570  0.116   -0.199    0.022
69                    ix  ~ vascular_risk_sumcorr           -0.106 0.057  -1.859  0.063   -0.218    0.006
70                    ix  ~                  eTIV           -0.195 0.065  -3.019  0.003   -0.322   -0.068
71                    sx  ~               age_M00           -0.196 0.117  -1.669  0.095   -0.425    0.034
72                    sx  ~                   sex            0.037 0.122   0.307  0.759   -0.201    0.276
73                    sx  ~               edyears            0.069 0.111   0.625  0.532   -0.148    0.286
74                    sx  ~ vascular_risk_sumcorr            0.057 0.103   0.551  0.582   -0.145    0.258
75                    sx  ~                  eTIV            0.097 0.113   0.854  0.393   -0.125    0.318
76                    iy ~~                    ix           -0.054 0.066  -0.825  0.409   -0.183    0.075
77                    iy ~~                    sx           -0.064 0.145  -0.445  0.656   -0.348    0.219
78                    sy ~~                    ix           -0.185 0.117  -1.578  0.114   -0.414    0.045
79                    sy ~~                    sx           -0.369 0.300  -1.228  0.219   -0.957    0.220
80               age_M00 ~1                                  0.000 0.000      NA     NA    0.000    0.000
81               edyears ~1                                  0.000 0.000      NA     NA    0.000    0.000
82                   sex ~1                                  0.000 0.000      NA     NA    0.000    0.000
83 vascular_risk_sumcorr ~1                                  0.000 0.000      NA     NA    0.000    0.000
84                  eTIV ~1                                  0.000 0.000      NA     NA    0.000    0.000
85               age_M00 ~~               age_M00            1.000 0.000      NA     NA    1.000    1.000
86                   sex ~~                   sex            1.000 0.000      NA     NA    1.000    1.000
87               edyears ~~               edyears            1.000 0.000      NA     NA    1.000    1.000
88 vascular_risk_sumcorr ~~ vascular_risk_sumcorr            1.000 0.000      NA     NA    1.000    1.000
89                  eTIV ~~                  eTIV            1.000 0.000      NA     NA    1.000    1.000
90               age_M00 ~~                   sex           -0.187 0.054  -3.446  0.001   -0.293   -0.080
91               age_M00 ~~               edyears           -0.128 0.058  -2.188  0.029   -0.242   -0.013
92               age_M00 ~~ vascular_risk_sumcorr            0.093 0.057   1.643  0.100   -0.018    0.205
93               age_M00 ~~                  eTIV            0.054 0.054   0.994  0.320   -0.052    0.160
94                   sex ~~               edyears           -0.244 0.050  -4.850  0.000   -0.343   -0.146
95                   sex ~~ vascular_risk_sumcorr           -0.198 0.052  -3.778  0.000   -0.301   -0.095
96                   sex ~~                  eTIV           -0.665 0.027 -24.937  0.000   -0.717   -0.612
97               edyears ~~ vascular_risk_sumcorr           -0.133 0.052  -2.543  0.011   -0.236   -0.031
98               edyears ~~                  eTIV            0.235 0.049   4.768  0.000    0.139    0.332
99 vascular_risk_sumcorr ~~                  eTIV            0.132 0.061   2.154  0.031    0.012    0.251
```
